# Reproducible Biochemical Modes Within a Structured Neurochemical Continuum in Autism Spectrum Disorder Revealed by NeuroCLAD

**DOI:** 10.64898/2026.05.07.26352658

**Authors:** Alok Sharma, Vinnit George, Hemangi Sane, Nandini Gokulchandran, Pooja Kulkarni, Siddhi Talgaonkar, Prerna Badhe

## Abstract

**Background:** Autism Spectrum Disorder (ASD) is biologically heterogeneous, yet most neurochemical studies rely on single-analyte comparisons that cannot capture coordinated variation across neurotransmitter systems. Whether peripheral neurochemical variation in ASD is best represented by discrete biochemical subtypes or continuous multivariate organisation remains unresolved.

**Methods:** We developed and applied NeuroCLAD, a structured multivariate analytical framework, to peripheral blood neurotransmitter profiles from 261 children with ASD (mean age 6.98 ± 3.13 years; 78.5% male). The pipeline combined age- and sex-adjusted multivariate profiling with principal component analysis, consensus k-means clustering with null-model validation, BIC-guided Gaussian mixture modelling, enrichment analysis, and clinical symptom mapping. Cross-compartment consistency was explored using urine neurotransmitter profiles from the same cohort.

**Results:** Twelve reproducible biochemical clusters were identified, each characterised by distinct multi-analyte neurochemical fingerprints involving trace amines, monoamines, catecholamine metabolism, histamine, and amino-acid neurotransmitter systems. Stability was confirmed across 200 bootstrap iterations and exceeded that expected under matched isotropic and covariance-preserving Gaussian null models. Bayesian Information Criterion guided Gaussian mixture model enumeration identified an optimal two-component solution comprising one dominant mixture component (n=222) and one smaller component characterized by elevated phenethylamine (PEA) and tyrosine (n=39). Behavioral mapping revealed graded symptom tendencies, particularly for aggressiveness, self-injurious behaviour, and picky eating. Formal symptom-cluster associations were non-significant after multiple-testing correction, consistent with graded rather than categorical relationships between peripheral neurochemistry and behavioural phenotype.

**Limitations:** Peripheral blood provides an indirect, analyte-dependent representation of neurochemical biology. The cross-sectional design, absence of a neurotypical comparison group, modest cluster sizes, and lack of external validation limit clinical interpretation.

**Conclusions:** Peripheral neurochemistry in ASD is organised into reproducible biochemical modes within a continuous multivariate landscape rather than discrete biochemical subtypes. This dimensional framework may better represent biological heterogeneity and provides a basis for future pathway-informed stratification and treatment-response studies.

## Background

Autism Spectrum Disorder (ASD) is a neurodevelopmental condition characterised by early-onset deficits in social communication and restricted or repetitive behaviours [1, 2]. Reported ASD prevalence has increased over recent decades, although estimates vary across populations, surveillance systems, diagnostic practices, and access to clinical services. ASD is widely recognised as a spectrum due to its marked clinical and biological heterogeneity, with substantial variation in symptom severity, developmental trajectory, and associated features [3, 4].

Current understanding suggests that ASD arises from a complex interplay of genetic and environmental factors, with contributions ranging from common polygenic variation to rare de novo mutations [5, 6, 7]. Despite this diversity, converging evidence points toward disruption of shared biological processes, including synaptic dysfunction, excitation–inhibition imbalance, altered neurochemical signalling, neuroinflammation, oxidative stress, and mitochondrial abnormalities [8, 9, 10]. Neuroimaging and neuropathological studies further highlight variability in brain volume, connectivity, and metabolic function across individuals with ASD, often yielding inconsistent or region-specific findings [11, 12]. For example, baseline positron emission tomography studies have demonstrated region-specific alterations in brain metabolism and atypical developmental trajectories across ASD populations [13]. Similarly, structural imaging studies have reported specific findings in white matter organisation, especially the corpus callosum, reflecting underlying biological heterogeneity [14]. Large retrospective clinical studies have shown variable outcomes of the same intervention which highlights the need to study subgroup populations which may have different internal biochemical patterns [15]. Collectively, these observations emphasize the need to identify recurring biological patterns within this variability.

Neurotransmitters and related metabolites provide an important biochemical window into the biological heterogeneity of ASD [16, 17]. Monoamines such as dopamine, serotonin, and norepinephrine regulate motivation, reward processing, mood, and attention, while glutamate and GABA govern excitatory-inhibitory balance, cortical connectivity, and sensory integration [8, 18]. Histamine acts as both a neurotransmitter and an immune modulator, contributing to arousal, sleep-wake regulation, and inflammatory signalling [19, 20]. Although circulating neurotransmitters and related metabolites do not provide a direct measure of central neurotransmission, peripheral neurochemical profiles reflect systemic biological processes, including autonomic regulation, metabolism, immune activity, and gut-brain interactions that are increasingly recognised as relevant to ASD. Furthermore, the relationship between peripheral and central neurochemistry is analyte-dependent, with some molecules showing closer physiological coupling to central pathways than others [21, 22]. Accordingly, the present study interprets peripheral neurochemical profiles as markers of systemic neurochemical organisation rather than direct surrogates of brain neurotransmission. Alterations across these systems are highly relevant to ASD phenotypes, as they collectively influence domains such as social interaction, behavioural regulation, and sensory processing [2, 16].

However, findings across neurotransmitter studies in ASD have been highly variable and often contradictory. While hyperserotonemia has been widely reported, profiles with normal or reduced serotonergic turnover are also frequently observed [17, 23]. Catecholaminergic alterations, including elevated norepinephrine and epinephrine, have been described in specific subsets, suggesting dysregulated autonomic and stress responses [24]. Dopamine metabolites such as homovanillic acid (HVA) and DOPAC exhibit variable turnover patterns, while glutamate-GABA signatures show inconsistent but biologically meaningful trends linked to excitatory-inhibitory imbalance [16, 25]. Histaminergic pathways further contribute to this complexity, particularly in individuals with sensory and immune-linked phenotypes [19, 20]. Importantly, these systems are rarely studied in conjunction, and most findings arise from single-analyte analyses, limiting the ability to interpret coordinated biochemical variation. As a result, the literature remains fragmented, with inconsistent findings that likely reflect underlying structured heterogeneity rather than true contradiction.

Methodologically, prior work is constrained by small sample sizes, single-analyte approaches, and reliance on case-control comparisons, which do not capture intra-ASD variability [3, 17]. Recent multivariate approaches have begun to address this gap. Notably, Ferraro et al. [26] demonstrated that amino acid patterns in ASD can be meaningfully explored using dimensionality reduction and clustering strategies, suggesting that coordinated biochemical variation may reveal latent structure not evident in univariate analyses [26]. Building on this premise, and motivated by the availability of a large, multi-analyte neurotransmitter dataset, we sought to systematically explore whether similar multivariate structure exists within ASD neurochemical profiles.

We developed NeuroCLAD (NeuroGen Consensus Layered Analytics for Discovery), a structured, genomics-inspired analytical framework designed for multivariate subtyping in high-variance biological datasets, and applied it here to blood neurotransmitter profiles in ASD [27, 28]. The pipeline adapts high-dimensional analytical strategies originally developed in cancer genomics and multi-omics clustering, and brings them into the domain of biochemical profiling in neurodevelopmental disorders [29]. While the individual analytical methods are well-established, their layered integration into a single coherent workflow, and their application to neurochemical heterogeneity in ASD, is to our knowledge novel. The workflow combines covariate residualisation, dimensionality reduction, consensus stability assessment, probabilistic membership modelling, and pathway-level enrichment.

Briefly, the pipeline begins with z-score normalisation and spline-based covariate residualisation to remove non-linear age effects and adjust for sex [30]. Principal component analysis is then used to capture the major axes of variance and denoise the data [31], and k-means clustering is applied in the reduced space to resolve recurrent patterns of biochemical organisation [32]. Cluster robustness is evaluated through consensus clustering with repeated subsampling [27], and Gaussian mixture modelling is layered on top to estimate probabilistic membership and assignment uncertainty [33, 34]. Pathway-level enrichment, alluvial mapping across resolutions, and clinical correlation then provide biological and behavioural annotation. This layered approach is designed to produce reproducible, interpretable, and biologically meaningful neurochemical patterns. Full methodological details are provided in the Methods section.

In this study, we applied NeuroCLAD to a large cohort of children with ASD (n=261) to identify reproducible multivariate patterns in blood neurotransmitter profiles. The central motivation was to address long-standing questions in the field, including whether ASD reflects discrete subtypes or a gradient-like continuum, and why traditional case-control or single-analyte approaches have produced inconsistent findings [3, 4]. Our goal was to determine whether coordinated, pathway-level variation, rather than isolated metabolite differences, could reveal reproducible biochemical organization within ASD and clarify its underlying structure [9]. Applying NeuroCLAD to this cohort, we identified novel, biologically coherent biochemical fingerprints embedded within a continuous neurochemical landscape. These findings provide a foundation for data-driven exploration of ASD heterogeneity and support a shift toward understanding variation as a structured continuum rather than discrete categorical subtypes [4, 35].

## Methods

### Study Design

This study analysed peripheral blood neurotransmitter profiles from 261 individuals diagnosed with Autism Spectrum Disorder [1, 2]. The study was carried out at the NeuroGen Brain and Spine Institute, Navi Mumbai, India. Male and female patients between the age group of 2 to 18 years with a confirmed diagnosis of Autism based on the DSM-5 criteria were included. All patients were assessed by standardized clinical assessments including scales like Childhood Autism Rating Scale (CARS) and the Gilliam Autism Rating Scale (GARS) [36, 37]. These assessments were used to guide clinical characterisation; specific symptom presence (binary variables) and verbal/non-verbal status were derived from these evaluations for downstream descriptive analyses.

Sample sizes vary across samples based on the availability of behavioural and clinical data for the specific analysis. Patients with any co-morbidities, acute medical illness, recent infections, or missing core clinical data were excluded. Individuals with relatively higher functional ability, particularly those with preserved adaptive functioning and advanced language skills, were not included, so that the cohort remained focused on moderate-to-severe ASD presentations. Clinical variables were used exclusively for post-hoc analyses and were not incorporated into clustering or dimensionality reduction.

The aim of this study is to characterise the structure of variation within the ASD population rather than to establish deviation from neurotypical norms. All clustering and effect-size analyses operate on within-cohort z-scores, and cluster-level descriptors such as “elevated dopamine” refer to relative position within the ASD distribution rather than to absolute deviation from population reference values. Integration of clustering output with laboratory reference ranges was not pursued, because such ranges are themselves derived from population calibration studies of variable composition, and the values used in our multivariate analyses are post-residualisation rather than raw concentrations. Univariate reference-range comparisons are reported separately for descriptive context only.

Blood samples were collected prospectively under a standardised institutional protocol. The study was approved by the Central Drugs Standard Control Organization (CDSCO)-registered Institutional Ethics Committee (IEC) of NeuroGen Brain and Spine Institute, Navi Mumbai (Approval No. NGBSI/IEC/AT-INV-01/2025/ISSUE-01/REVISION-01). Written informed consent was obtained from the parents or legal guardians of all participants. All data were anonymised prior to analysis.

### Blood Neurotransmitter Panel

Peripheral neurotransmitter assays were performed by a third-party, clinically accredited laboratory using liquid chromatography–tandem mass spectrometry (LC–MS/MS). Plasma samples underwent protein precipitation followed by LC–MS/MS analysis using a multiple reaction monitoring (MRM) method in positive electrospray ionisation mode. Quantification was based on analyte-to-internal standard peak area ratios against calibration curves. The resulting assay values were provided as clinical reports and used as the basis for all downstream analyses. The analytes included in the analyses were

1. Catecholaminergic axis: Dopamine, Epinephrine, Norepinephrine, 3,4-Dihydroxyphenylacetic acid (DOPAC), 3-methoxytyramine (3-MT), normetanephrine, metanephrine
2. Trace amines and aromatic precursors: Phenethylamine (PEA), tyramine, tyrosine
3. Serotonergic axis: Serotonin (5-HT)
4. Excitatory-inhibitory amino acids: Glutamate, Gamma-aminobutyric acid (GABA), glycine
5. Histaminergic/other modulators: Histamine, Taurine

Raw concentrations were reported in their native units (pg/mol, ***μ***mol/L, ***μ***g/ml, pmol/L, nmol/L, ng/ml, pmoles/ml), depending on the analytes. Reference intervals were not rescaled or altered. Z-score normalisation was subsequently performed on the raw concentrations for multivariate analyses [29].

### Univariate analysis relative to laboratory reference ranges

Prior to multivariate analysis, raw blood neurotransmitter concentrations were examined relative to established laboratory reference intervals. For each analyte, values were classified as low, within range, or high based on the corresponding laboratory reference limits. The proportion of individuals falling into each category was calculated to provide an overview of cohort-level biochemical distributions. These analyses were performed on unadjusted raw concentrations and were intended to characterise baseline variability rather than contribute to subtype identification. These univariate summaries were used solely for descriptive context and were not incorporated into the multivariate modelling pipeline.

### Data Preparation (Pre-Processing and Quality Control)

All analyses were performed in R (version 4.5.2). Core packages included tidyverse (*dplyr*, *tidyr*, *ggplot2*), *cluster*, *mclust*, *pheatmap*, and *splines*, with *uwot* used for supplementary dimensionality visualisation.

### Data Handling

The pipeline was designed with the provision to consider analytes with >15% missingness to be excluded, samples missing >25% of analytes to be removed entirely, and if remaining missing values (<5%) were present, they were to be imputed using k-nearest neighbor (k=5) imputation applied on standardized data to preserve covariance structure [38]. Missingness was assessed prior to all downstream analysis (Supplementary Figure S1).

### Data Normalisation

As the neurotransmitters were measured in different units and concentration ranges, an initial z-score transformation was performed for diagnostic inspection and visualisation of the raw data. For all downstream multivariate analyses, spline residualisation was applied to the raw analyte concentrations to adjust for age and sex. The resulting residuals were then standardised to z-scores before principal component analysis, ensuring that each analyte contributed equally regardless of its original scale or residual variance and preventing clustering from being disproportionately influenced by differences in measurement scale [29].

### Covariate Adjustment via Natural Cubic Spline Regression

Age is a key confounder that exhibits non-linear effects on several neurotransmitters (e.g., serotonin peaking in childhood, catecholamine shifts during adolescence) [17, 24]. Similarly, sex differences also are known to influence monoamines [17]. To remove these demographic confounders while preserving biologically meaningful variance, each analyte was adjusted using a linear regression model incorporating a natural cubic spline term for age and a fixed-effect term for sex [30]. For each analyte, the model y ∼ ns(Age, df = 3) + Sex was fitted. Residuals from these models were extracted, standardised to z-scores, and used in all downstream multivariate analyses [30]. This approach provides a consistent and conservative correction across analytes and ensures that age- and sex-related variance does not dominate the principal component structure or clustering output [30, 31].

### Multivariate Analysis Pipeline (NeuroCLAD)

#### Overview

NeuroCLAD is a structured multivariate workflow designed to identify underlying stable biochemical patterns or data-driven subtypes from a high-variance dataset. After preprocessing (normalisation and covariate residualisation), the pipeline proceeds through a sequence of dimensionality-reduction, clustering, and stability-assessment stages [29]. First, principal component analysis (PCA) reduces noise and captures major variance components [31]. K-means clustering is then applied in the PCA space to generate initial subtype solutions [32], which are evaluated across multiple values of k using within-cluster dispersion and silhouette coefficients [40]. Low-dimensional visualisations of the PCA space are provided in supplementary analyses. Cluster robustness is subsequently quantified using consensus clustering with repeated subsampling [27]. Gaussian mixture modelling is used to assess probabilistic cluster membership and quantify overlap between clusters, with entropy used to characterise uncertainty in assignment. Finally, cluster profiles are annotated using enrichment analyses and clinical correlation.

#### Principal Component Analysis (PCA)

PCA is a method that turns many correlated variables into a smaller set of uncorrelated “summary variables” or “components” that capture the major patterns in the data [31].

Z-scored neurotransmitter matrix obtained after covariate-adjustment was used for multivariate analyses downstream. PCA was applied using the standard singular value decomposition (SVD) implementation (R prcomp), without additional centering or scaling, as all variables were standardized during pre-processing. PCA decomposes the covariance structure into orthogonal components ordered by the intrinsic variance [31].

For each component, the proportion of variance explained and the cumulative variance were calculated. A scree plot with cumulative variance was plotted and examined to determine how many components captured the majority of the signal without the noise [31]. The smallest number of principal components (PCs) that together explained approximately 70% of the total variance (as is usually the common practice) was retained for further downstream clustering analysis. The first 10 principal components (PCs) were retained which produced a reduced-dimension representation of the neurotransmitter space without the noise and mitigated collinearity among the analytes [31].

Age and sex columns in the dataset were carried forward as metadata and used only for visual inspection for the PCA scatter plot. The plot is coloured by sex and sized by age. This was done in order to confirm that spline residualisation had been effective in minimizing demographic gradients in the multivariate structure [30].

#### K-Means Clustering

Unsupervised clustering was performed to identify reproducible biochemical structure within the cohort, with an emphasis on stability rather than enforcing strictly discrete categories. K-means clustering partitions the dataset into k groups by iteratively assigning individuals to the nearest cluster centroid, maximising within-cluster similarity and between-cluster separation [32].

Clustering was conducted on the retained principal component (PC) space using Euclidean distance and multiple random starts to minimise sensitivity to initialisation [32]. To explore the appropriate number of clusters, k was varied systematically from 2 to 20. For each k, within-cluster sum of squares (WCSS) and average silhouette width were computed to assess cluster compactness and separation [40]. Silhouette widths were computed using the ‘*cluster*’ package in R.

Based on these diagnostics and the distribution of cluster sizes, a set of candidate resolutions (k = 8, 10, 12, 15, 20) was selected for further stability assessment using consensus clustering.

For interpretability, representative centroid profiles (mean analyte z-scores) were examined at k=3, 12, and 15, corresponding to coarse, intermediate, and higher-resolution partitions of the data (Supplementary Figures S5A and B). Among the evaluated solutions, k=12 was selected as the primary grouping, with k=15 considered as a higher-resolution refinement (Figure 5).

For each candidate k, cluster sizes and centroid profiles were examined, and heatmaps of these centroids were used to assess whether clusters represented biologically coherent biochemical patterns rather than arbitrary partitions.

#### Consensus Clustering or Bootstrapping (Cluster Stability Assessment)

Consensus clustering was applied as a robustness assessment to evaluate the stability of cluster assignments under repeated subsampling and data perturbation. Consensus clustering evaluates how reproducible a cluster solution is by repeatedly re-clustering random subsets of the data and quantifying how consistently individuals group together [27].

Consensus clustering was used to assess the stability of k-means solutions and prevent over-interpretation of unstable partitions [28]. For each candidate k (8, 10, 12, 15, 20), 200 bootstrap iterations were performed. In each iteration, approximately 80% of individuals and 80% of retained PCs were randomly subsampled without replacement, and k-means was re-run on this subset [27]. A consensus matrix was generated by computing, for each pair of individuals, the proportion of iterations in which they were assigned to the same cluster. The goal was to evaluate how consistently the same individuals co-clustered across repeated perturbations of the data. This provides a direct measure of cluster robustness [27].

Cluster-wise stability was quantified as the mean within-cluster consensus, representing how consistently individuals in that group clustered together across resamples [27]. Higher values reflect more reproducible and internally coherent clusters.

To summarise overall stability for each k, we calculated the Proportion of Ambiguous Clustering (PAC), defined as the proportion of consensus values that fall within an intermediate range (0.1–0.9) [28]. Lower PAC values indicate clearer separation between pairs that consistently cluster together and those that consistently do not, and therefore reflect more stable and interpretable solutions [28].

#### Consensus Cluster Null-Model Validation

To assess whether the observed cluster structure exceeded that expected from data lacking reproducible multivariate organisation, the consensus clustering procedure was repeated on two matched Gaussian null models constructed in the retained principal component space (261 individuals × 10 components) [40]. The first was an isotropic Gaussian null, in which each principal component was sampled independently from a standard normal distribution, representing the absence of multivariate structure. The second was a covariance-preserving Gaussian null generated from a multivariate normal distribution parameterised by the empirical mean vector and covariance matrix of the observed principal component scores, thereby preserving the overall covariance structure while eliminating reproducible cluster organisation. Each null dataset (n = 200 per model) was analysed using the identical consensus clustering procedure applied to the observed data (k=12; 200 bootstrap resamples of 80% of individuals and 80% of principal components; 50 random starts per k-means run; PAC thresholds 0.1–0.9). The proportion of ambiguous clustering (PAC) and mean within-cluster consensus were recorded for each null dataset and compared with the observed values obtained from the primary consensus clustering analysis. Empirical one-sided p-values were calculated as the proportion of null datasets exhibiting stronger apparent structure than the observed data (lower PAC or higher mean consensus, respectively). Where no null dataset exceeded the observed value, empirical significance is reported as p < 0.005, reflecting the resolution limit of 200 null simulations.

#### Bayesian Information Criterion (BIC)-Guided Mixture Enumeration

To independently assess the latent mixture complexity of the neurochemical data, Gaussian mixture models were fitted in the retained principal component space using the ‘*mclust’* package [33, 34]. Models comprising G=1-20 mixture components were evaluated under the equal-volume, equal-shape, equal-orientation (EEE) and variable-volume, variable-shape, variable-orientation (VVV) covariance parameterisations. The optimal model was selected using the Bayesian Information Criterion (BIC), where higher values indicate improved penalised model fit under the ‘*mclust’* convention. For the optimal solution, posterior membership probabilities and maximum a posteriori component assignments were extracted for each individual, and model-derived components were cross-tabulated against the primary k=12 partition to examine correspondence between the two analytical approaches. Component membership was subsequently compared across age (two-sample t-test) and sex (Fisher’s exact test).

#### Gaussian Mixture Modelling (Probabilistic Cluster Assignment)

To characterise probabilistic membership within the consensus-derived biochemical clusters, Gaussian Mixture Modelling (GMM) was applied in the same principal component (PC) space used for clustering. Unlike k-means clustering, which assigns each individual to a single cluster, GMM estimates the probability of membership across clusters for each individual, allowing assignment confidence and transitional biochemical profiles to be examined [33].

GMM was implemented using the ‘mclust’ package in R [34]. For consistency with the NeuroCLAD framework, probabilistic membership was estimated for the consensus clustering resolutions evaluated within the pipeline using the EEE and VVV covariance parameterisations.

Posterior probabilities of cluster membership were obtained for each individual. The cluster with the highest posterior probability was defined as the maximum posterior assignment, serving as a soft cluster label. The maximum posterior probability was used as a measure of assignment confidence, with higher values indicating stronger affiliation with a single biochemical subtype. The distribution of these values across individuals was examined to assess the degree of cluster separation.

#### Membership Entropy (Assignment Uncertainty)

Uncertainty in cluster membership was summarised by computing membership entropy based on the posterior probability distribution for each individual. Entropy was calculated as the Shannon entropy of the posterior probability vector across clusters [41].

Lower entropy values indicate confident assignment to a single cluster, whereas higher values reflect individuals positioned between cluster distributions. These measures were used to characterise assignment uncertainty and identify individuals occupying transitional positions within the continuous neurochemical landscape.

#### Analyte-Level Enrichment And Pathway Annotation (Cohen’s d)

To biologically characterise the emerging biochemical clusters, we performed analyte-level differential profiling for each cluster against the remainder of the cohort. For every analyte, a two-sample t-test was conducted comparing standardized neurotransmitter levels inside the cluster versus all other individuals combined. From each comparison we extracted: (i) the p-value, (ii) the direction and magnitude of change using Cohen’s d (standardised mean difference), and (iii) the mean analyte levels inside and outside the cluster [42].

Multiple comparisons across analytes were controlled using the Benjamini–Hochberg false discovery rate (FDR), with FDR-adjusted p < 0.05 considered statistically significant [43]. The sign of Cohen’s d was used to classify each significant analyte as relatively “high” or “low” within that cluster [42].

To support biological interpretation, analytes were grouped into functionally meaningful categories (e.g., dopaminergic, noradrenergic, serotonergic, GABAergic, glutamatergic, histaminergic, trace amines, catecholamine precursors/metabolites). For each cluster, we summarised: (i) the number of FDR-significant analytes, (ii) the strongest effect sizes, and (iii) a concise “fingerprint” describing key high/low analytes together with their pathway assignments.

These results were collated into cluster-level fingerprint tables and visualised through heatmaps of Cohen’s d values across analytes and clusters. This enrichment framework enabled each subtype to be annotated in terms of dominant neurotransmitter patterns (e.g., catecholamine-high, serotonin-low, or glutamate-high/GABA-low axes), providing a biologically interpretable description of the cluster structure. Heatmaps were generated using the *pheatmap* package in R.

#### Mapping Clusters To Qualitative Symptoms

To connect the biochemical clusters with behavioural presentation, cluster assignments were merged with an independently collected qualitative symptom dataset. This dataset was constructed from therapist-recorded clinical files. The dataset contained, for each patient, binary values (0/1) of specific behavioural and sensory features recorded by therapists (e.g., hyperactivity, aggression, social interaction, eye contact, stereotypical behaviour, self-injury, picky eating, sensory issues), as well as a binary verbal/non-verbal status. Five individuals were excluded from verbal–non-verbal classification due to incomplete or inconsistent behavioural records and were therefore not included in analyses involving this classification. These individuals were retained for all biochemical, PCA, clustering, and enrichment analyses. Accordingly, verbal–non-verbal status was available for 256 of the 261 participants.

Data preprocessing involved harmonising patient identifiers across files, removing empty or non-informative fields, and recoding qualitative responses into numeric form. Entries such as “A/NA”, “Yes/No”, or “Present/Absent” were converted to 1/0 values. Verbal status was derived from how the GARS was administered. The GARS includes additional verbal subscales that are applied only to children with sufficient expressive language; children evaluated with these subscales were classified as verbal, and those evaluated using only the core subscales (excluding the verbal-specific subscales) were classified as non-verbal [37]. Five children fell below the lower age cutoff for GARS classification and were therefore not assigned a verbal/non-verbal status. This classification was used for descriptive purposes only and was not treated as a validated measure of language ability.

For each cluster, the proportion of individuals exhibiting each behavioural feature was calculated using participants with available data for that symptom. These incidence values were assembled into cluster-by-symptom tables and visualised as annotated heatmaps to provide descriptive behavioural fingerprints. To formally assess whether symptom prevalence differed across clusters, Pearson’s chi-square tests were performed for each binary symptom. Where more than 20% of expected cell counts were below five, p-values were estimated using Monte Carlo simulation (10,000 replicates) [44]. Resulting p-values were adjusted for multiple testing using the Benjamini-Hochberg (BH) false discovery rate procedure [43].

#### Cross-Solution Correspondence And Alluvial Mapping (k=12 vs k=15)

To understand how the primary 12-cluster solution related to the higher-resolution 15-cluster partition, we constructed two types of flow diagrams connecting the k=12 and k=15 clusters [27].

1. **Enrichment-based overlap** For each k, we expanded the cluster fingerprint tables into long format, listing all FDR-significant analytes per cluster. We then identified analytes that were significantly associated with clusters in both solutions and counted, for each pair of (k=12 cluster, k=15 cluster), how many enriched analytes they shared. These counts were visualised as a two-column alluvial diagram, with the left side representing k=12 clusters, the right side representing k=15 clusters, and ribbon width proportional to the number of shared enriched analytes. This map highlighted how biochemical themes from the 12-cluster solution were split or refined in the 15-cluster space.
2. **Sample-based overlap** Cluster assignment tables for k=12 and k=15 were merged by individual. For each pair of clusters across the two solutions, we counted the number of shared individuals and constructed a second ribbon diagram with ribbon width proportional to patient overlap. This allowed us to visualise how individual patients migrated between clusters when moving from 12 to 15 groups and to identify k=15 clusters that represented subdivisions of specific k=12 groups.

Together, these alluvial mappings provided an interpretable bridge between resolutions, supporting the choice of k=12 as a compact and stable summary while showing that the k=15 solution largely represented coherent refinements rather than a completely different partitioning [28].

#### Supportive Cross-Compartment Urine Analysis

The NeuroCLAD framework was independently applied to urine neurotransmitter profiles from the same cohort (n = 261) to examine whether broadly comparable biochemical organisation could be identified in a second biological compartment. Preprocessing, z-score normalisation, and covariate adjustment for age and sex using natural cubic spline regression were performed identically to the blood analysis [30]. Clustering was conducted on residualised, z-scored urine analytes using PCA followed by k-means and consensus clustering across k= 8–20 [25, 29, 30]. Cluster stability was evaluated using the proportion of ambiguous clustering (PAC) and average within-cluster consensus [27, 28].

Downstream characterisation focused on effect-size–based enrichment (Cohen’s d) for analytes shared between blood and urine, enabling direct cross-fluid comparison of pathway-level signatures rather than exact cluster replication [42]. Creatinine and derived ratio variables were excluded from clustering to avoid confounding by urine concentration effects.

The analytical pipeline (NeuroCLAD) is available in a public repository for reproducibility (see Data Availability statement).

## Results

### Participant Characteristics

A total of 261 children with autism spectrum disorder were included in the analysis. The mean age was 6.98 ± 3.13 years (median 6 years, interquartile range 4 years; range 2–18 years). The cohort comprised 205 males (78.5%) and 56 females (21.5%) [7, 35]. Verbal status, derived from GARS, was available for 256 participants, of whom 171 (66.8%) were classified as verbal and 85 (33.2%) as non-verbal as five individuals fell below the lower age cutoff for GARS verbal/non-verbal classification. Baseline demographic characteristics of the cohort are summarised in Table 1.

**Table 1:** Demographic characteristics of the study cohort. *Summary of the study cohort characteristics.* The table summarises the baseline demographic profile of 261 individuals with autism spectrum disorder included in the analysis. Age is reported in years and presented as mean ± standard deviation, median, interquartile range, and range.

| Variable | Summary |
| --- | --- |
| No. of Patients | 261 |
| Mean Age (years) | 6.98 ± 3.13 |
| Median Age (years) | 6 |
| Age IQR (years) | 4 |
| Age Range (years) | 2-18 |
| Male, n | 205 |
| Female, n | 56 |
| Male (%) | 78.54 |
| Female (%) | 21.46 |
| Verbal, n (%) | 171 (66.8) |
| Non-verbal, n (%) | 85 (33.2) |

### Univariate Distribution of Blood Neurotransmitter Levels

The distribution of raw blood neurotransmitter concentrations was examined relative to established laboratory reference intervals. Substantial heterogeneity was observed across multiple analytes (Table 2).

**Table 2.**
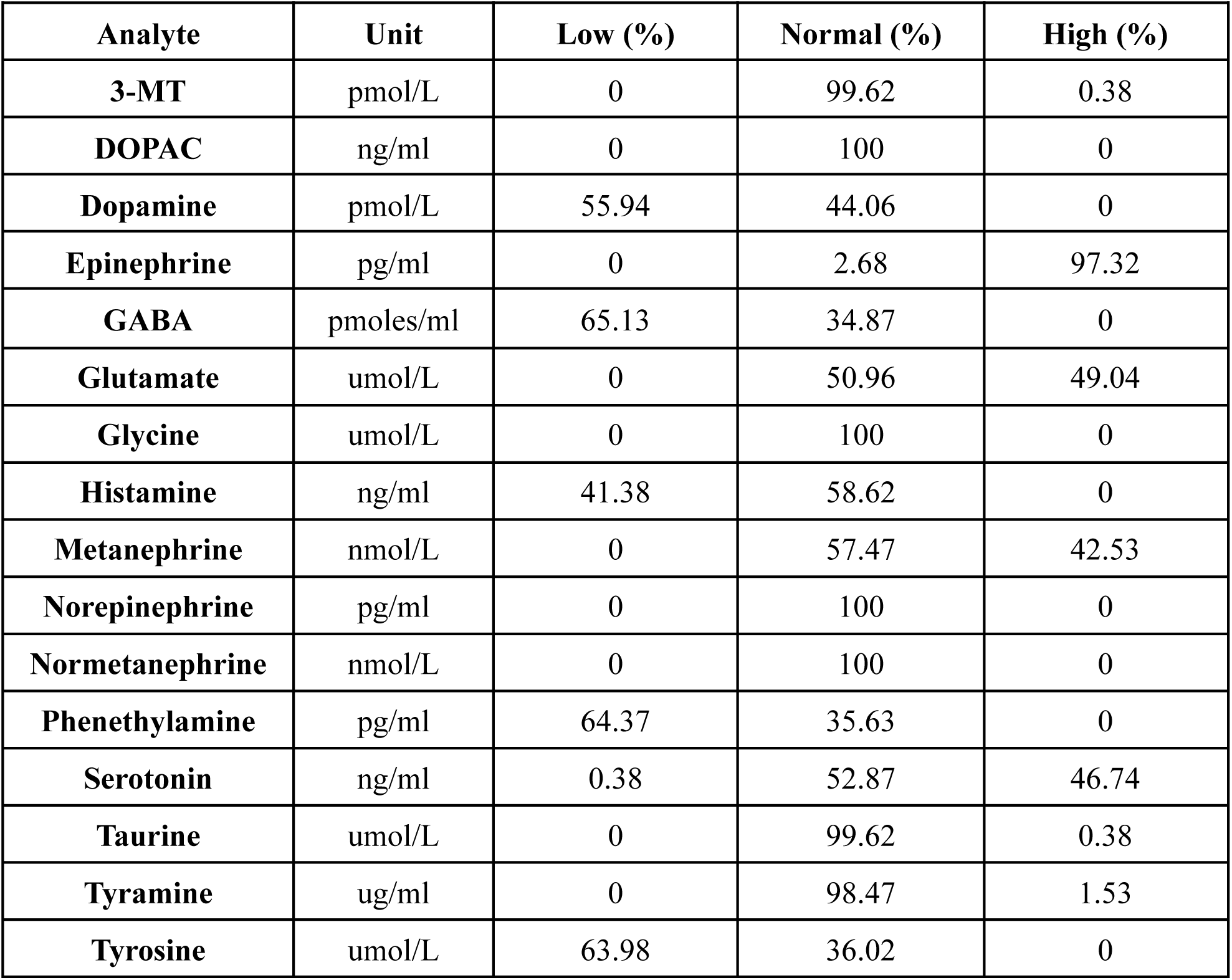
Univariate distribution of blood neurotransmitter levels relative to laboratory reference ranges. Percentage distribution of blood neurotransmitter levels relative to laboratory reference ranges. Raw blood neurotransmitter concentrations were classified as low, within range (normal), or high based on established laboratory reference intervals for each analyte. Values are expressed as the percentage of individuals in each category and were calculated using unadjusted concentrations prior to any preprocessing, covariate adjustment, or multivariate analysis.

| Analyte | Unit | Low (%) | Normal (%) | High (%) |
| --- | --- | --- | --- | --- |
| **3-MT** | pmol/L | 0 | 99.62 | 0.38 |
| **DOPAC** | ng/ml | 0 | 100 | 0 |
| **Dopamine** | pmol/L | 55.94 | 44.06 | 0 |
| **Epinephrine** | pg/ml | 0 | 2.68 | 97.32 |
| **GABA** | pmoles/ml | 65.13 | 34.87 | 0 |
| **Glutamate** | umol/L | 0 | 50.96 | 49.04 |
| **Glycine** | umol/L | 0 | 100 | 0 |
| **Histamine** | ng/ml | 41.38 | 58.62 | 0 |
| **Metanephrine** | nmol/L | 0 | 57.47 | 42.53 |
| **Norepinephrine** | pg/ml | 0 | 100 | 0 |
| **Normetanephrine** | nmol/L | 0 | 100 | 0 |
| **Phenethylamine** | pg/ml | 64.37 | 35.63 | 0 |
| **Serotonin** | ng/ml | 0.38 | 52.87 | 46.74 |
| **Taurine** | umol/L | 0 | 99.62 | 0.38 |
| **Tyramine** | ug/ml | 0 | 98.47 | 1.53 |
| **Tyrosine** | umol/L | 63.98 | 36.02 | 0 |

Several neurotransmitters exhibited marked deviations from reference ranges in large proportions of the cohort. Notably, reduced concentrations were prevalent for GABA (65.1%), phenethylamine (64.4%), tyrosine (64.0%), and dopamine (55.9%). In contrast, elevated concentrations were prominent for epinephrine (97.3%), serotonin (46.7%), metanephrine (42.5%), and glutamate (49.0%).

Other analytes, including glycine, norepinephrine, normetanephrine, and DOPAC, largely remained within reference limits in the majority of participants. Collectively, these findings highlight pronounced inter-individual biochemical variability that cannot be adequately captured by single-analyte interpretation [17, 45]. We therefore hypothesised that this variability reflects underlying multivariate structure within the blood neurotransmitter data, motivating the application of unsupervised clustering approaches to identify latent biochemical subgroups within the cohort [3].

### Age-Related Non-Linear Effects In Blood Neurotransmitters

Prior to multivariate analysis, we examined whether neurotransmitter concentrations varied systematically with age and sex in the raw dataset. Several analytes showed visible age-related structure, with the clearest example observed in GABA. As seen in the raw scatterplot, GABA concentrations were tightly grouped in younger children (approximately 3-6 years of age) and became progressively dispersed across later childhood and adolescence. This developmental curvature indicated that age contributed substantial, non-linear variance that, if left uncorrected, could dominate the downstream PCA and clustering structure [17, 24].

Following adjustment of each analyte using a natural cubic spline term for age and a fixed-effect term for sex, the residualized GABA values no longer displayed an age-related trend (figure 1). Inter-individual variability was preserved, but the developmental curvature was effectively removed, confirming successful alleviation of demographic confounding [30]. Comparable attenuation of age-related patterns was observed across multiple neurotransmitters (Supplementary Figure S2).

**Figure 1.**
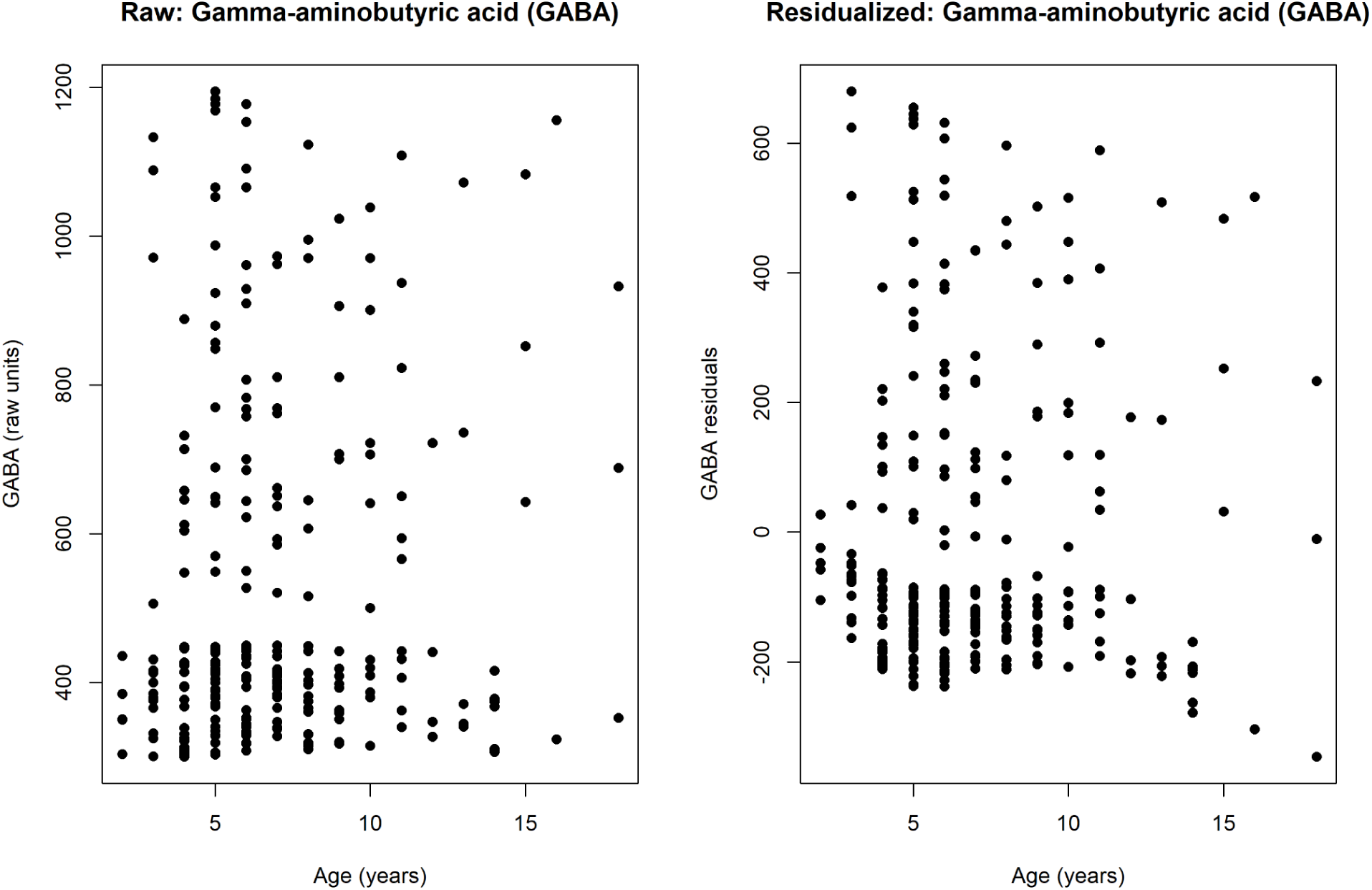
Age-related structure in GABA concentrations before and after spline residualisation. Raw values (left) show strong non-linear age curvature and dispersion across childhood and adolescence. Natural cubic spline residualisation (right) removes this developmental trend, yielding age-adjusted residuals suitable for unbiased multivariate analysis.

All subsequent PCA and clustering analyses were therefore performed on the age and sex-adjusted residual matrix to ensure demographic effects did not drive the underlying multivariate structure.

### Principal Component Analysis (PCA)

Following age and sex variable adjustment, PCA was performed on the standardized residual matrix to characterize the multivariate structure intrinsic to the dataset [31]. The scree plot (Figure 2) demonstrated a gradual decline in variance explained across components. There was no abrupt ‘elbow’ which would be consistent with a distributed, multi-analyte signal rather than one dominated by a small subset of metabolites [29]. The first 10 principal components (PCs) together accounted for approximately 70% of the total variance. These 10 PCs were retained for subsequent clustering analyses.

**Figure 2.**
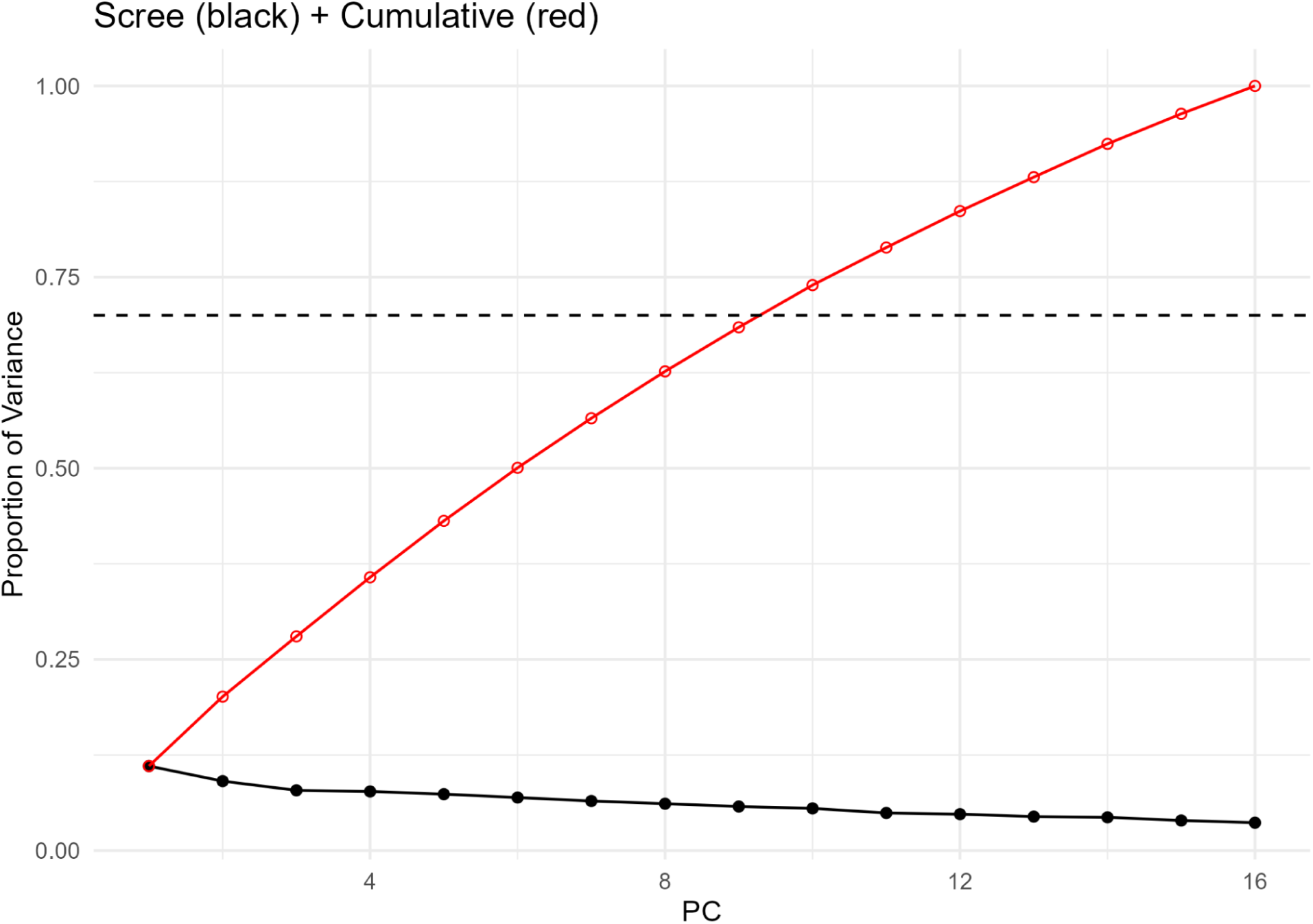
Scree plot (black) and cumulative variance (red) explained for the age- and sex-adjusted neurotransmitter dataset. The first few principal components capture only a modest proportion of variance, with the cumulative curve reaching approximately 70% by PC10, indicating a diffuse, continuum-like structure typical of heterogeneous biochemical datasets.

The PCA scores were visualized on a scatter plot with points coloured by sex and sized by age (Figure 3). It prompted the verification of removal of the demographic confounders during the spline regression step. No visible gradients or segregation by age or sex were observed along PC1 or PC2, confirming that residual demographic effects did not drive the multivariate patterns in the clusters. The sample distribution appeared isotropic and well mixed across both axes. This provided no visual evidence that residual age or sex gradients dominated the leading principal components [30].

**Figure 3.**
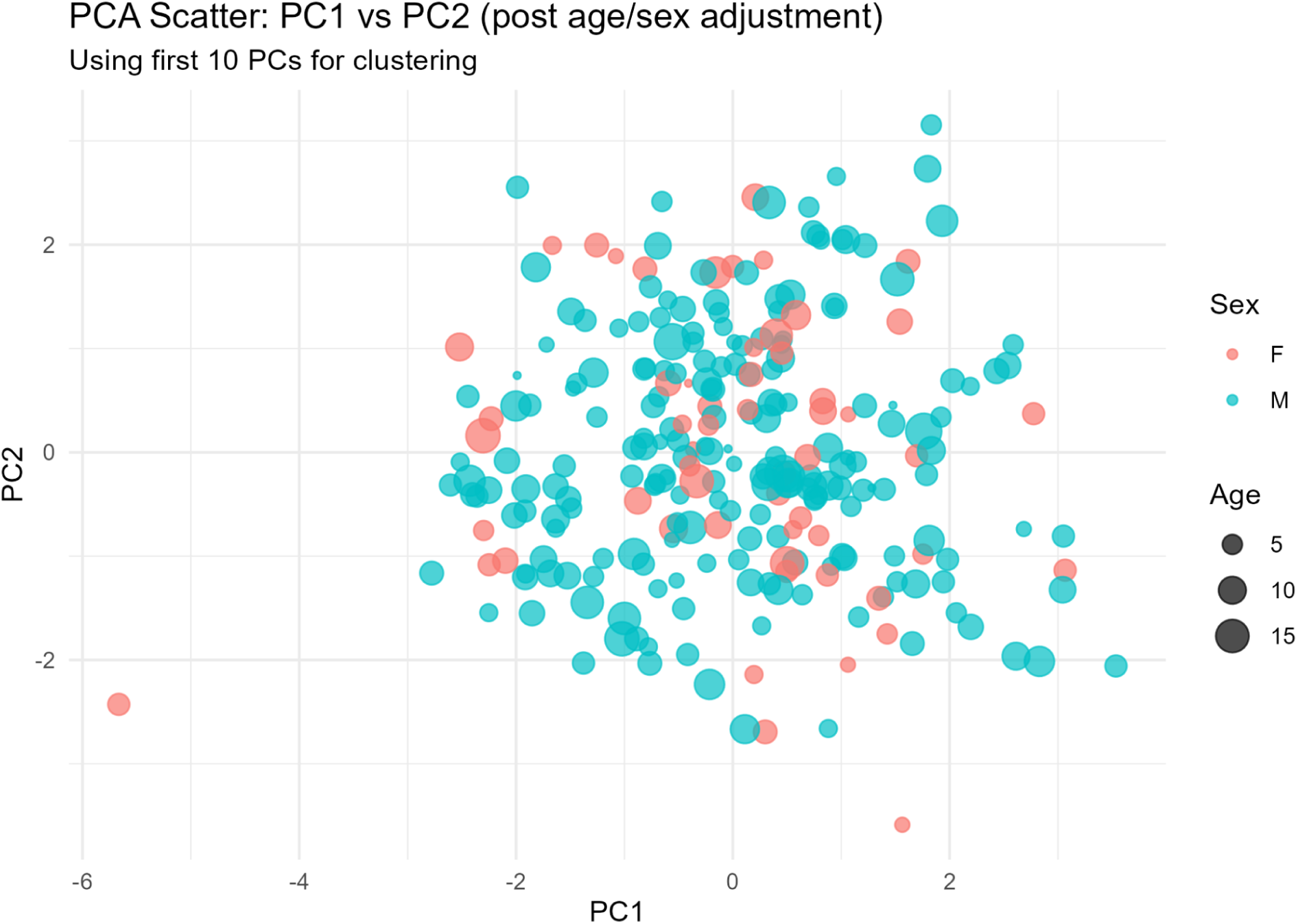
PCA scatterplot of age and sex-adjusted neurotransmitter data. PC1 and PC2 show no strong separation by sex or age, indicating that major demographic effects were effectively removed prior to downstream clustering.

These PCs formed the input of all downstream unsupervised analyses including k-means and Gaussian mixture modelling (GMM), consensus stability analysis and substructure enrichment.

### K-means Clustering

K-means clustering was applied to the retained PC space to explore potential biochemical subgroups within the cohort [32]. Across k = 2–20, the elbow curve demonstrated a smooth decline in ‘within-cluster sum of squares’ without a sharp inflection point (Figure 4A). This pattern provided no evidence of a sharply preferred discrete partition and was compatible with a broadly continuous underlying structure [4]. The silhouette curve increased gradually with k (Figure 4B), indicating improving separation at higher cluster resolutions [39].

**Figure 4.**
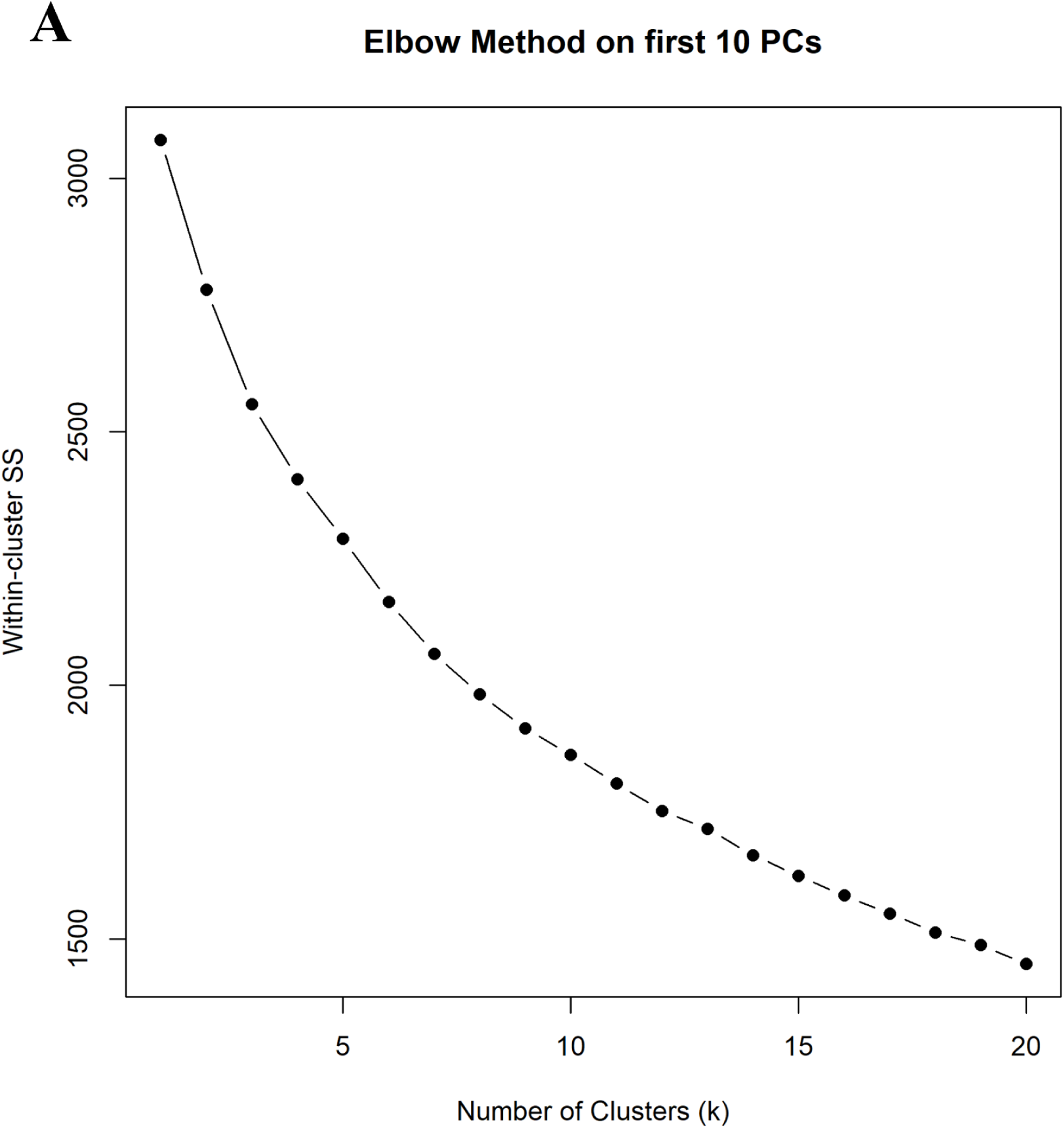
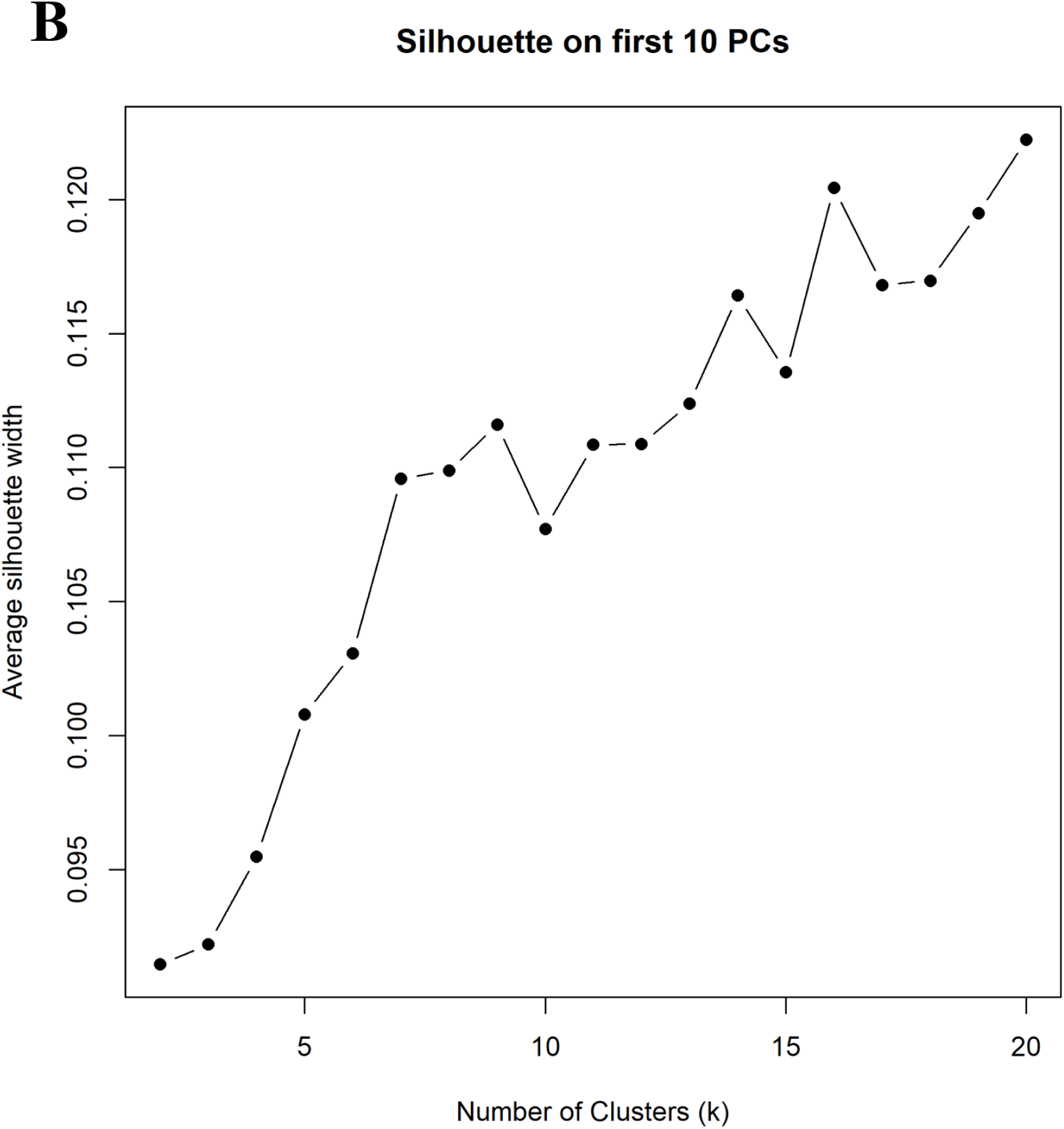
Elbow (A) and silhouette (B) analyses for k-means clustering on the first 10 principal components. (A) The elbow plot shows a progressive reduction in ‘within-cluster sum of squares’ with increasing k, with the rate of improvement flattening beyond k = 10–12. (B) Silhouette analysis demonstrates modest but increasing average silhouette widths across k, with relatively improved structure emerging around k=12–15. Together, these metrics indicate no single optimal k but support exploring moderately higher cluster solutions, consistent with the heterogeneous biochemical landscape of ASD.

Guided by these diagnostics, we selected several candidate solutions for closer inspection (k = 3, 12, and 15), reflecting low to high-resolution representations of the underlying structure. Examination of centroid heatmaps (Figure 5 for k12; and Supplementary Figure S5A for k=3 and S5B for k=15) showed that these solutions produced distinct biochemical signatures rather than noise-driven or arbitrary partitions.

**Figure 5.**
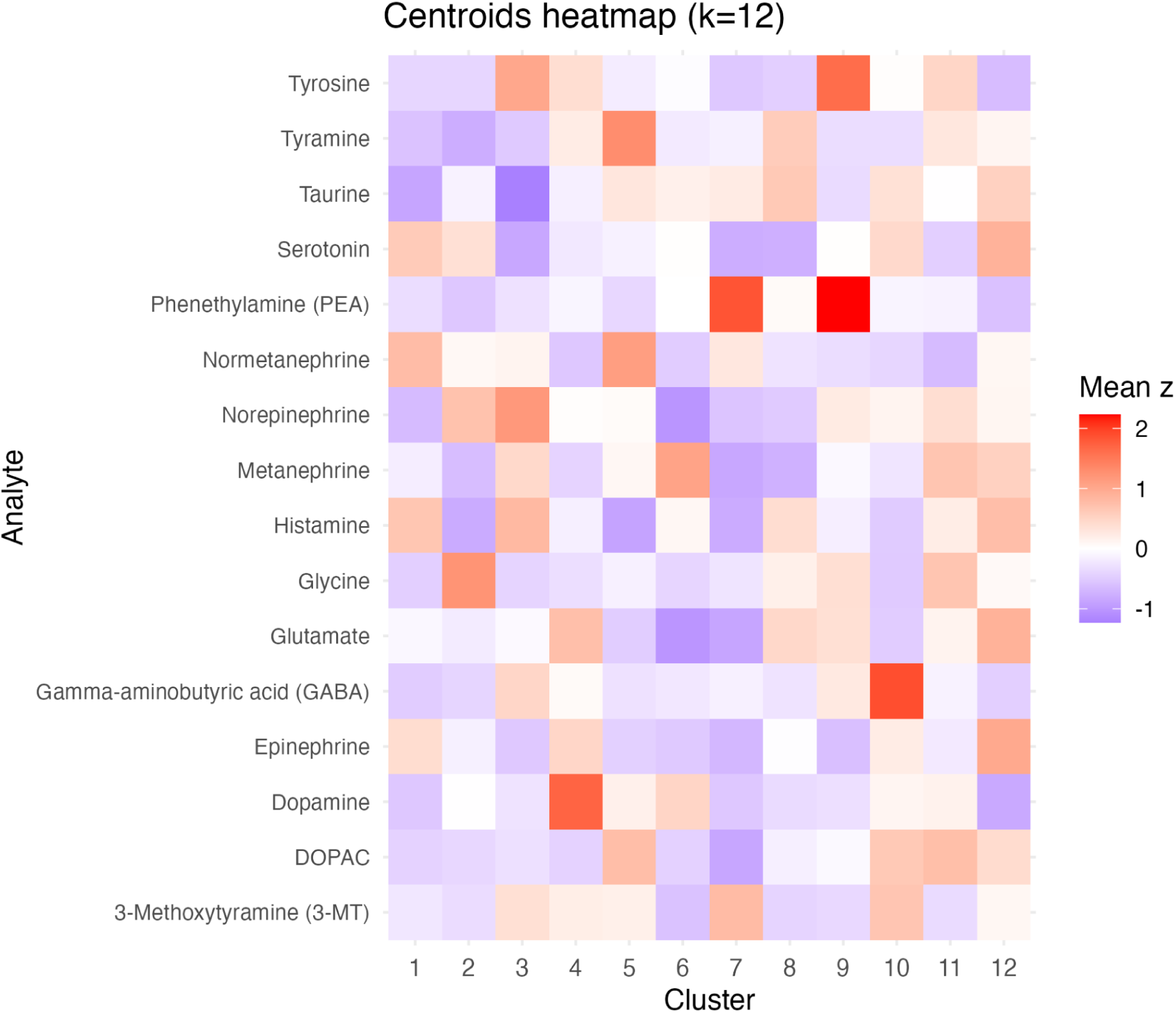
Cluster centroids heatmap for k=12. Heatmap of mean z-scored neurotransmitter levels across the 12 biochemical clusters, illustrating distinct monoaminergic and amino-acid related signatures.

The k=12 solution provided the clearest balance between resolution and interpretability. Several clusters displayed distinct monoaminergic and amino acid-related biochemical signatures, including [8, 16]:

- PEA-dominant profiles with elevated phenethylamine accompanied by moderate and low serotonin and tyrosine
- Catecholamine-elevated profiles characterised by higher dopamine, norepinephrine, and DOPAC
- Histamine-inclined profiles with accompanying reductions in GABA/glutamate
- Amino-acid neurotransmitter–shifted profiles involving glycine, glutamate, and related metabolites

These coherent axes of variation persisted at k=15 but were subdivided into smaller, more granular patterns, whereas k=3 compressed multiple distinct signatures into broader, less informative groupings.

The centroid structures identified at k=12 guided subsequent stability analyses and formed the basis for the biological interpretation of cluster-specific neurotransmitter patterns. Full centroid matrices are provided in (Supplementary Table S3)

### Consensus Clustering and Cluster Stability Assessment

The robustness of the candidate cluster solutions was evaluated using bootstrap consensus clustering across five values of k (8, 10, 12, 15, 20). For each k, 200 bootstrap iterations were generated, each consisting of 80% of individuals and 80% of retained PCs sampled without replacement. Consensus matrices were computed as the proportion of times each pair of individuals co-clustered across iterations, providing a direct estimate of cluster reproducibility under perturbation [27].

Across the evaluated solutions, cluster stability showed a characteristic pattern. The Proportion of Ambiguous Clustering (PAC) plot (Fig 6B) captures the fraction of pairwise consensus scores falling within the ambiguous range of 0.1–0.9, decreasing monotonically with increasing k (Table 3). This progression indicates a clear separation between consistently co-clustering and non-co-clustering pairs at higher values of k. However, if PAC alone would be used as a selection metric, it would favor very high-k solutions. Therefore, we also evaluated mean within-cluster consensus as a measure of average stability. This is designed to capture the internal cohesion and reproducibility of each cluster across bootstrap iterations [27]. As expected, average stability decreased as k increased (Table 3). This reflects the expected loss of reproducibility when partitions become increasingly fine-grained. Notably, while the k=12 solution showed a modest reduction in stability relative to k=8 and 10, it retained substantially higher cluster cohesion than higher k-models (Supplementary figure S7A).

**Figure 6.**
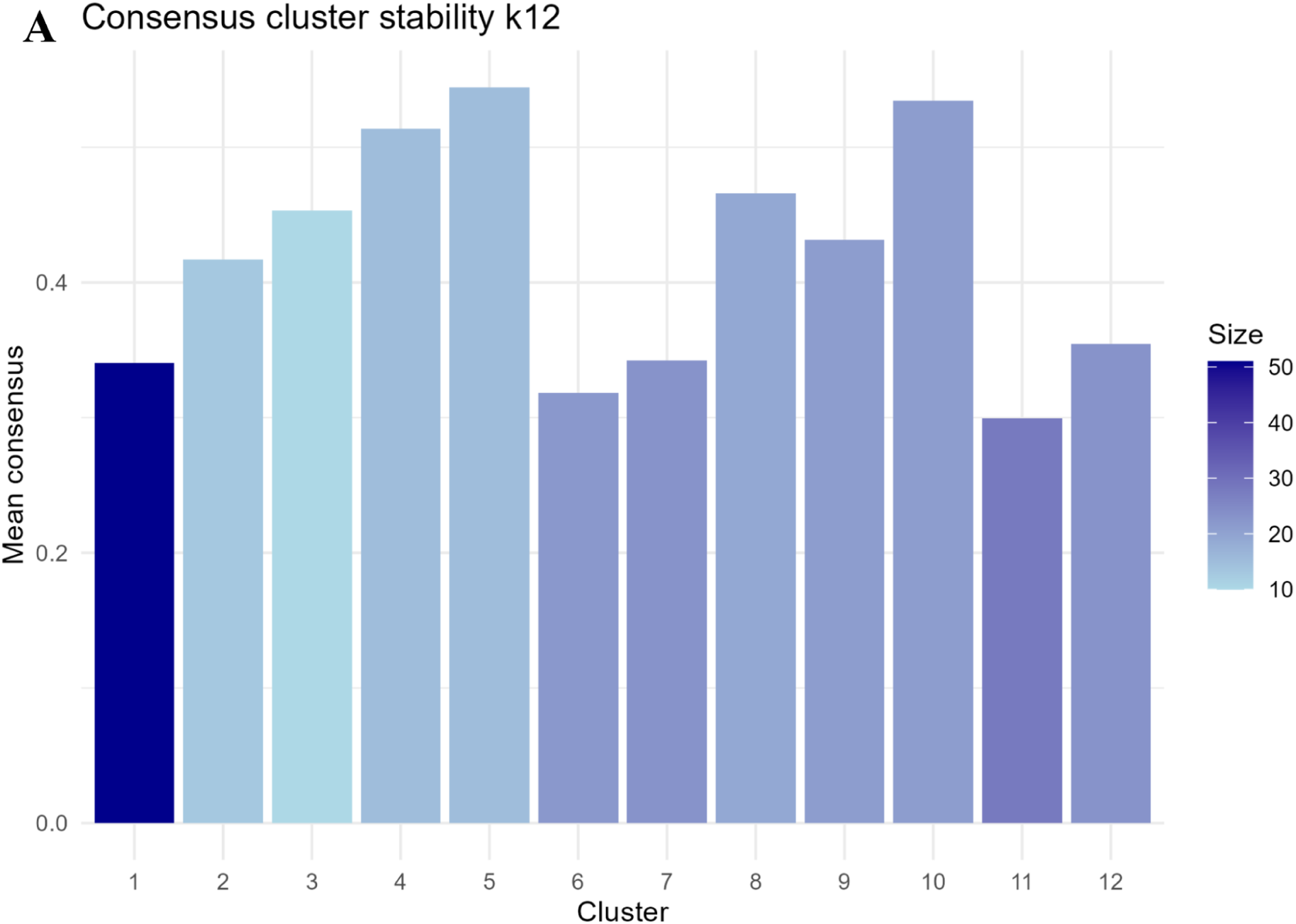
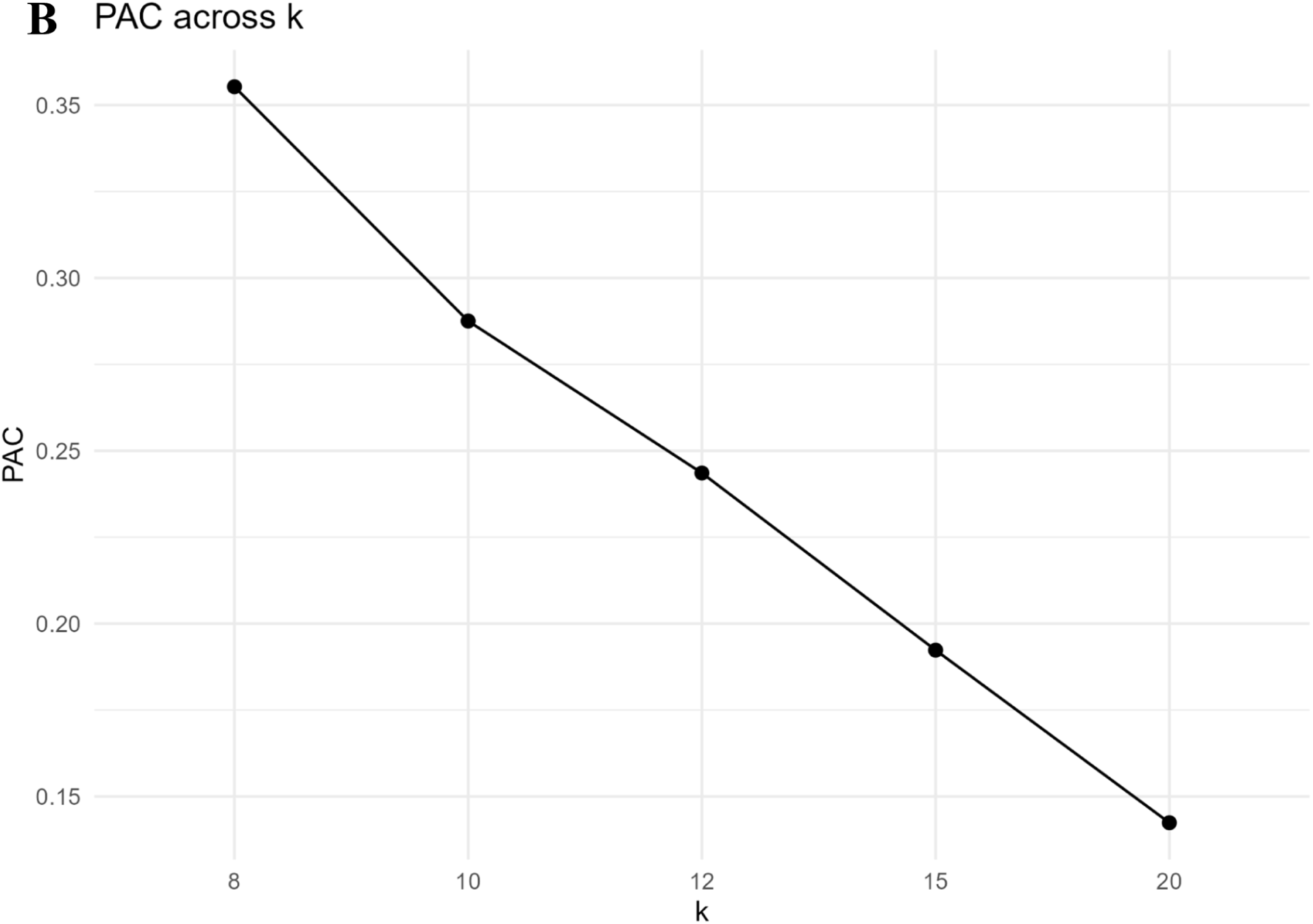
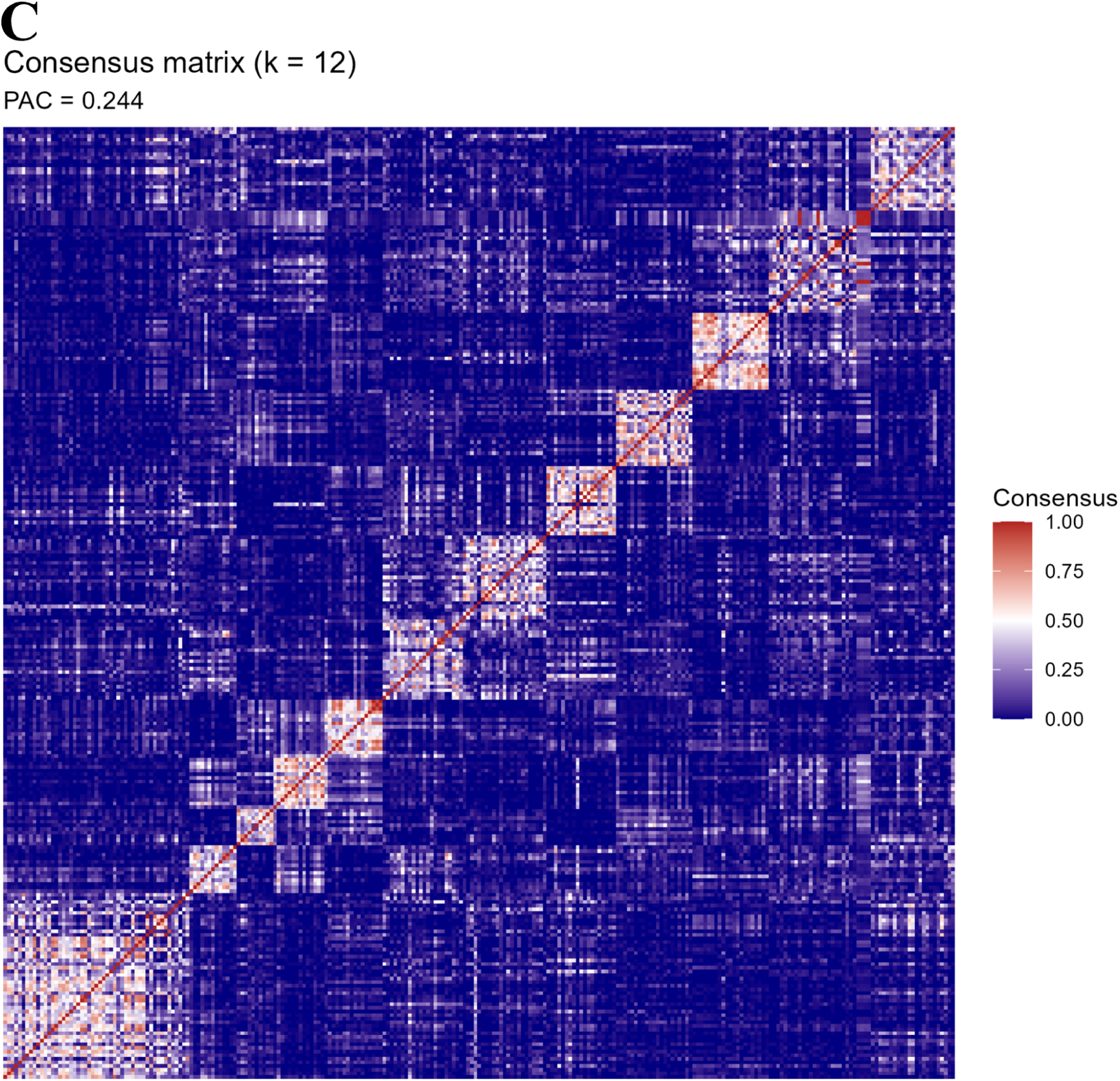
Consensus clustering diagnostics across candidate k values. **(A)** Cluster-wise mean consensus values for the *k=12* solution, with bar colour indicating cluster size. Several clusters exhibit moderate to high internal stability, whereas others display weaker cohesion, an expected pattern in heterogeneous biochemical datasets. **(B)** Proportion of Ambiguous Clustering (PAC) across k = 8–20, showing a steady decline as k increases. Lower PAC values reflect sharper consensus boundaries at higher k, though at the cost of increasing fragmentation. **(C)** Consensus matrix for *k=12*, illustrating the frequency with which pairs of individuals co-clustered across bootstrap iterations. Block-like structures indicate reasonably well-defined clusters, supporting k=12 as a balanced solution between resolution and stability.

**Table 3.** Consensus clustering stability metrics across k = 8–20. *Stability indices from consensus clustering across candidate k values.* Proportion of Ambiguous Clustering (PAC) decreases steadily from k = 8 to k = 20, reflecting sharper consensus boundaries at higher cluster numbers. Average within-cluster stability shows a mild downward trend, consistent with increasing fragmentation of clusters as k increases.

| **k** | **Proportion of**<br>**Ambiguous**<br>**Clustering (PAC)** | **Average**<br>**within-cluster**<br>**stability** |
| --- | --- | --- |
| 8 | 0.355 | 0.490 |
| 10 | 0.288 | 0.431 |
| 12 | 0.244 | 0.418 |
| 15 | 0.192 | 0.396 |
| 20 | 0.142 | 0.394 |

The k=12 consensus matrix (Figure 6C) displayed a set of clearly recognisable diagonal blocks corresponding to the 12 clusters, although a few clusters showed moderate boundary diffusion, reflecting underlying heterogeneity. Despite this, the overall structure remained interpretable and coherent, especially when contrasted with the higher-k solutions, which appeared visually sharper but achieved this clarity through over-fragmentation into smaller, less stable clusters [27]. The modest cross-cluster blending observed for a subset of clusters at k=12 corresponded to groups with slightly lower mean within-cluster consensus values in the bar plot (Figure 6A), consistent with the behaviour expected in moderately heterogeneous clusters. Additional diagnostics, including cluster size versus stability trends (Supplementary Figure 7B) further supported the conclusion that very high-k partitions yielded more unstable over-fragmented clusters without providing additional interpretable structure.

Together, the low PAC, preserved average stability, and coherent cluster boundaries, identified k=12 as a reproducible and interpretable operational partition for downstream biochemical characterisation. This configuration was therefore selected for deriving biochemical fingerprints and downstream clinical associations.

It is worth noting that the absolute magnitude of these stability metrics is modest relative to those reported in domains such as cancer subtyping. We interpret this as biologically informative rather than methodologically problematic, and return to this point in the Discussion.

### Null-Model Validation of Cluster Structure

To determine whether the observed consensus partition exceeded structure expected by chance, the k=12 solution was compared with matched isotropic and covariance-preserving Gaussian null models. The observed partition yielded a proportion of ambiguous clustering (PAC) of 0.244 and a mean within-cluster consensus of 0.418.

Against the isotropic Gaussian null, both metrics indicated stronger structure than expected by chance (median null PAC = 0.261, empirical p < 0.005; median null consensus = 0.367, p = 0.005). Against the covariance-preserving Gaussian null, PAC likewise remained significantly lower than expected (median = 0.259, p < 0.005), whereas mean within-cluster consensus remained significantly, though more modestly, elevated (median = 0.377, p = 0.015). Complete null distributions for both clustering metrics are shown in (Supplementary Figures S8A and B and summarised in Supplementary Table S8).

### Bayesian Information Criterion (BIC)-guided Mixture Enumeration

To independently examine the latent mixture complexity of the neurochemical data, Gaussian mixture models were fitted across G = 1–20 components in the retained principal component space using the *mclust* framework. Among the fitted solutions retained in the BIC scan, the highest BIC was obtained for the two-component EEE model. Bayesian Information Criterion (BIC) identified a two-component solution (G = 2) as the optimal model (BIC = −8086.5), with progressively lower BIC values observed for models comprising larger numbers of components (Supplementary Table S9; Supplementary Figure S9).

The optimal solution consisted of one large component containing 222 individuals and a smaller component containing 39 individuals. Cross-tabulation with the primary k=12 partition demonstrated that the smaller component corresponded predominantly to consensus Clusters 7, 8 and 9, encompassing all individuals assigned to Cluster 9 (14/14), most of Cluster 7 (12/15), and a subset of Cluster 8 (7/22), together with a small number of individuals distributed across the remaining clusters (Supplementary Table S10). These consensus clusters were characterised by the strongest phenylethylamine-related enrichment observed in the dataset, with Cluster 9 additionally exhibiting marked tyrosine elevation.

No significant differences were observed between the two mixture components with respect to age (6.74 ± 2.96 vs. 7.02 ± 3.17 years; Welch’s t-test, p = 0.600) or sex distribution (79.5% vs. 78.4% male; Fisher’s exact test, p = 1.00), indicating that the identified mixture components were not explained by demographic composition.

Model-based mixture enumeration therefore favoured a parsimonious two-component representation of the overall neurochemical density, whereas consensus clustering resolved a reproducible twelve-cluster operational partition of the same dataset. These analyses characterise complementary aspects of the underlying neurochemical landscape and are considered together in the Discussion.

### Gaussian Mixture Modelling (Probabilistic Cluster Membership)

Gaussian Mixture Modelling (GMM) was applied to evaluate the degree of probabilistic separation between the clusters using the k=12 solution in the same principal component (PC) space. GMM provides posterior probabilities of cluster membership for each individual, allowing assessment of both assignment confidence and overlap between clusters.

The distribution of maximum posterior probabilities (Figure 7) was strongly skewed toward higher values, with a substantial proportion of individuals exhibiting posterior probabilities close to 1. This indicates that many individuals are assigned to clusters with high confidence, reflecting regions of high assignment confidence within the operational k=12 partition [33, 34]. At the same time, a smaller subset of individuals showed lower maximum posterior values, indicating partial membership across multiple clusters and suggesting the presence of transitional profiles between neighbouring biochemical modes.

**Figure 7.**
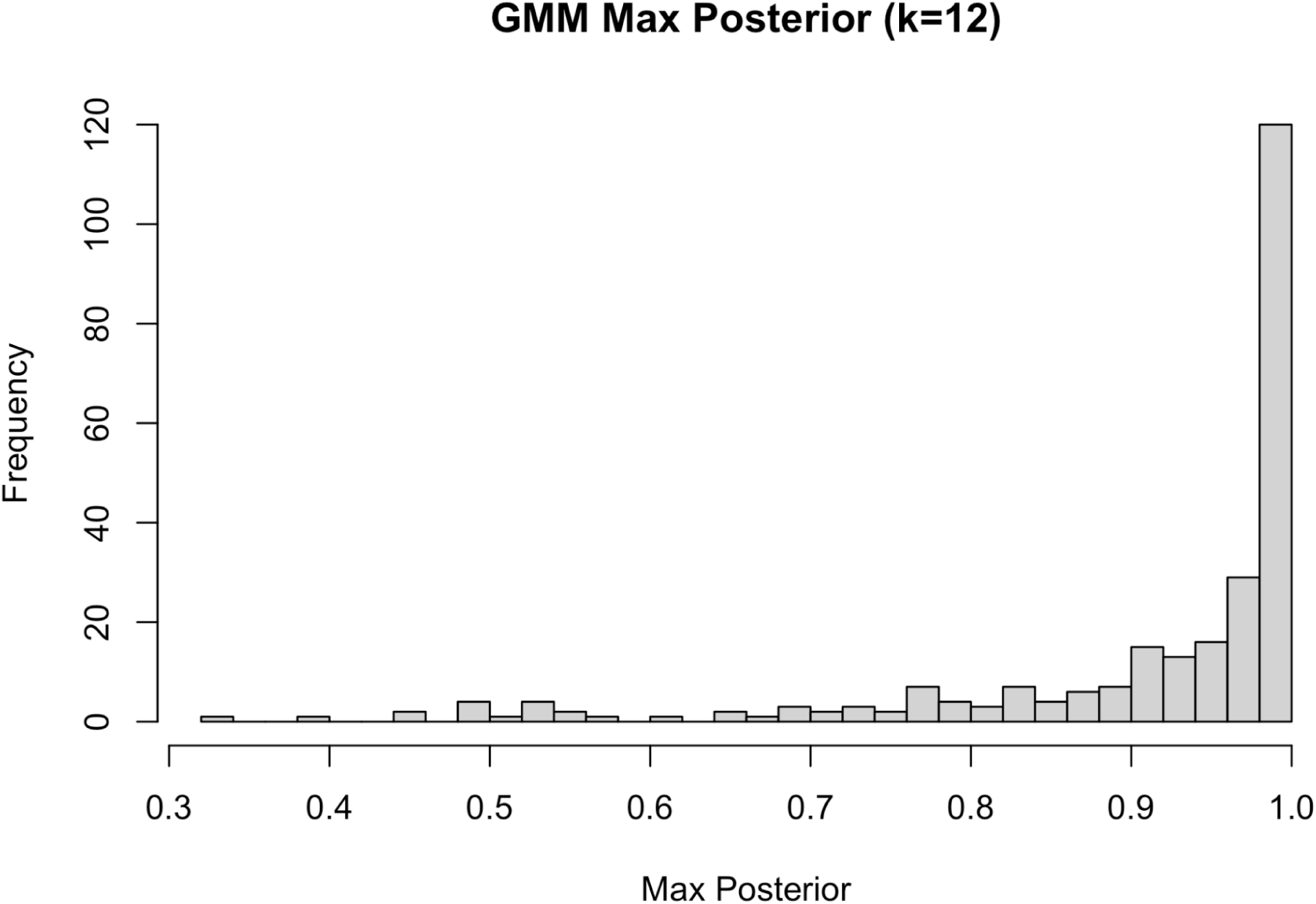
Gaussian mixture modelling distribution for k=12 solution. Distribution of maximum posterior probabilities from Gaussian mixture modelling (k=12). The histogram shows that most individuals exhibit high posterior probabilities (approaching 1), indicating strong cluster assignment, while a minority display lower values consistent with overlapping or intermediate profiles.

Taken together, the fixed k=12 Gaussian mixture model demonstrates that many individuals occupy reproducible biochemical regions with high assignment confidence, while others display more gradual transitions between neighbouring clusters. These findings complement the consensus clustering results by characterising local assignment uncertainty within the operational k=12 partition, rather than by determining the number of latent biochemical components, which was evaluated separately using BIC-guided model enumeration. [33].

### Membership Entropy (Assignment Uncertainty)

Assignment uncertainty in clusters was studied using entropy computed from the posterior probability distributions for each individual. Lower entropy values indicate concentrated membership in a single cluster, whereas higher values reflect more distributed membership across clusters [41].

In the k=12 solution, entropy values were strongly skewed toward lower values (figure 8), consistent with the high-confidence assignments observed in the maximum posterior distribution. This indicates that a large proportion of individuals occupy well-defined regions within the biochemical space. However, a tail of higher entropy values was observed, corresponding to individuals with more diffuse membership across clusters, indicative of intermediate or boundary profiles.

**Figure 8.**
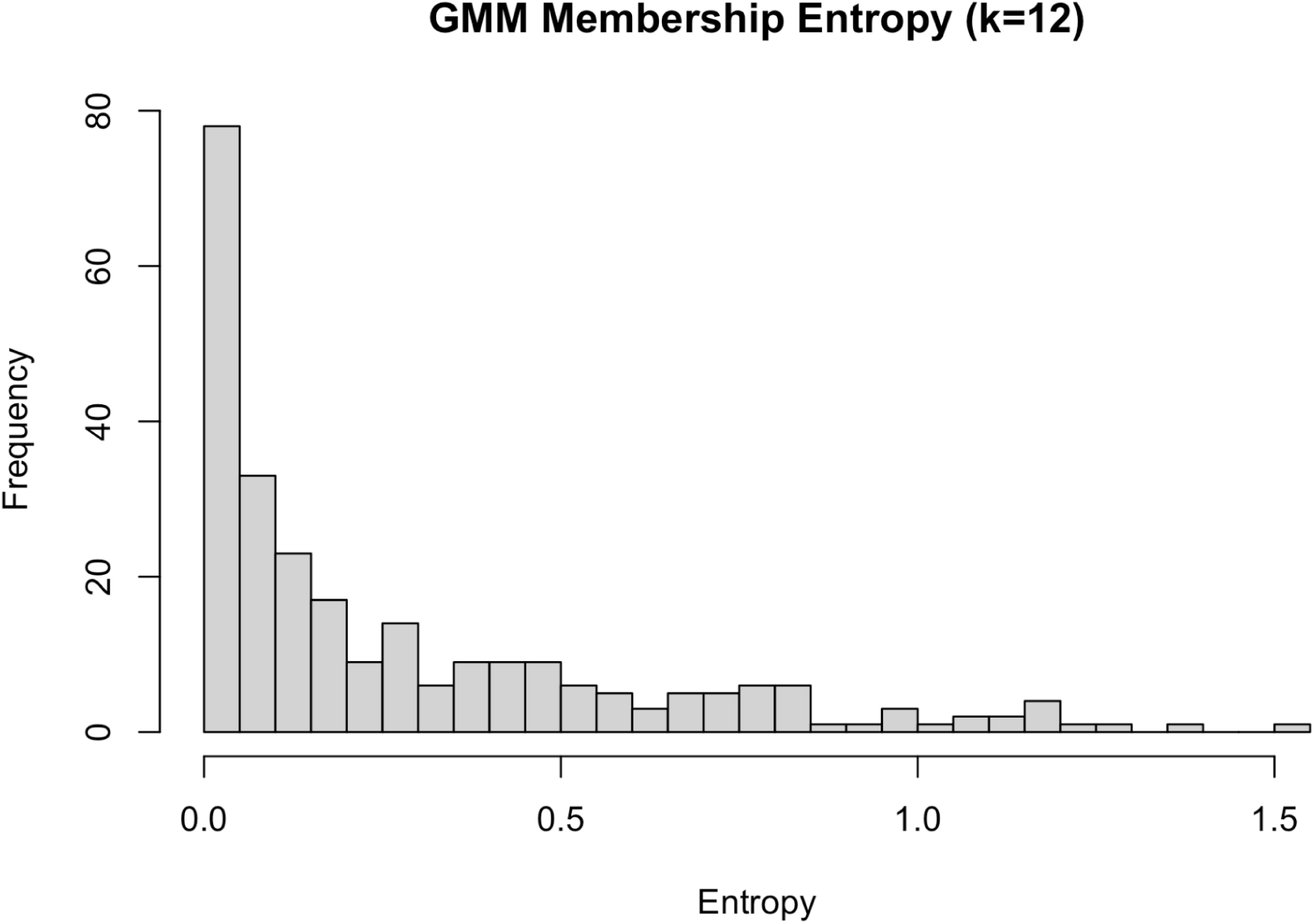
Membership entropy distribution calculated from posterior probability distributions of the k =12 solution. Distribution of membership entropy derived from Gaussian mixture modelling (k=12). Lower entropy values dominate, indicating confident cluster assignments, while a subset of individuals with higher entropy reflects transitional positioning between clusters.

Together, the maximum posterior and entropy analyses indicate that the biochemical clusters identified by k-means correspond to probabilistically well-supported structures with strong internal coherence, while still retaining a degree of overlap consistent with a continuous multivariate landscape.

The corresponding entropy distribution for the k=15 solution showed a similar pattern (Supplementary Figure 10B), indicating that increased resolution produces finer partitions without substantially altering the underlying probabilistic structure.

### Quantitative Enrichment (Cohen’s d) and Interpretive Biological Fingerprints

We performed analyte-level enrichment analyses to quantify the biochemical distinctiveness of each cluster in the k=12 solution. We used Cohen’s d effect sizes and FDR-adjusted t-tests comparing each cluster against the remainder of the cohort. The effect-size matrix (Figure 9) showed that several analytes shifted together within each cluster, indicating that the clusters reflected real underlying biochemical patterns rather than noise from the k-means algorithm [42, 43].

**Figure 9.**
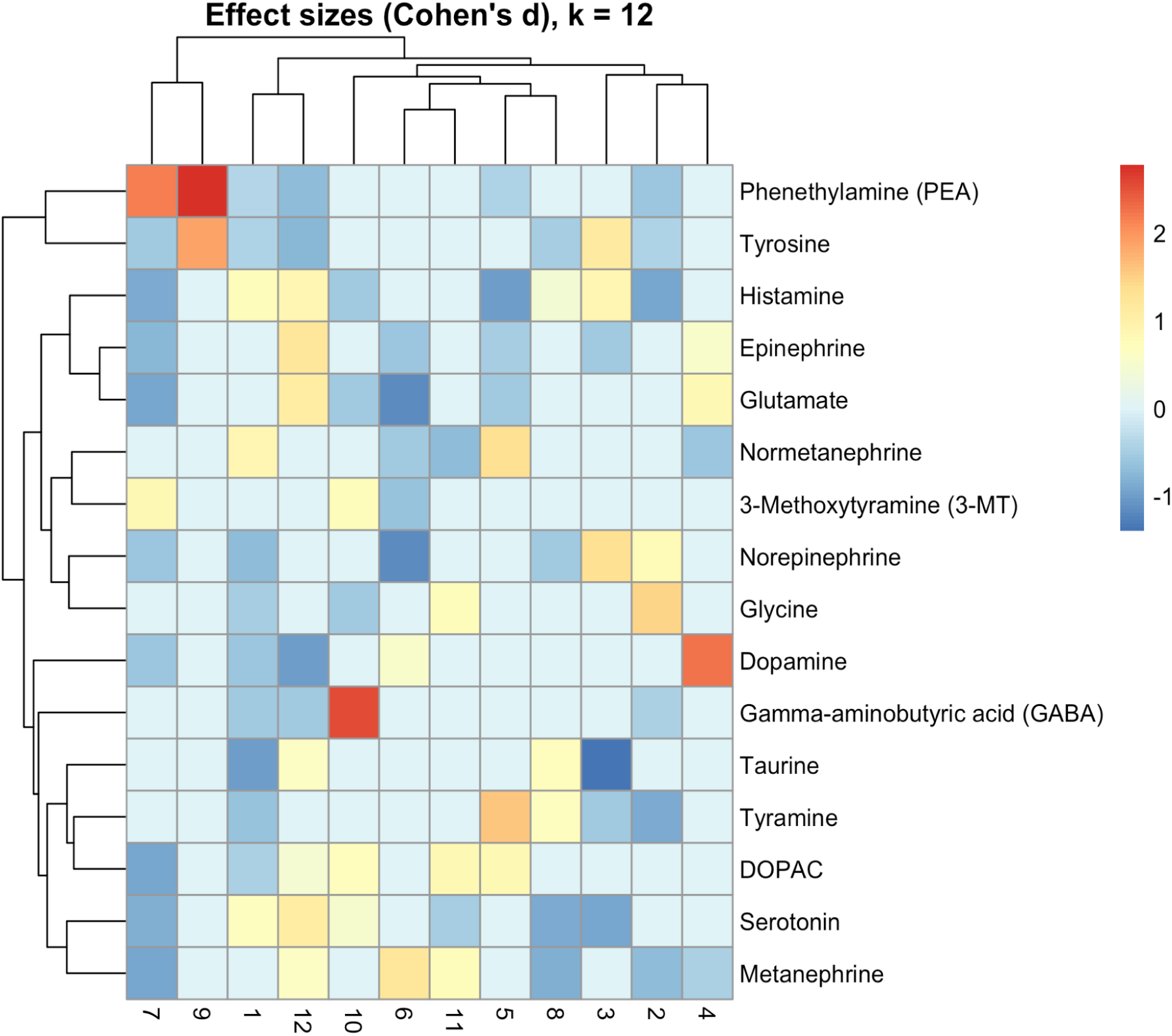
Cohen’s d effect sizes for neurotransmitter profiles across the 12 data-driven clusters. The heatmap displays standardized mean differences (Cohen’s d) comparing each cluster with the remainder of the cohort for each analyte. Warmer colours indicate elevations and cooler colours reductions within each cluster. Several clusters show distinct biochemical signatures, including strong phenethylamine elevations, dopamine shifts, and amino-acid related deviations, supporting the presence of interpretable multivariate patterns underlying the k=12 clustering solution.

Across the 12 clusters, the number of FDR-significant analytes ranged from 2 to 12, demonstrating heterogeneous but reproducible biochemical deviation patterns. Effect sizes varied in magnitude from small (|d| ≈ 0.3) to very large (up to d > 2.0 for trace amines such as phenethylamine). Both positive and negative shifts were observed across monoamines, catecholamine metabolites, inhibitory/excitatory amino acids, and histaminergic modulators, indicating that the clusters captured multi-dimensional variations rather than single-analyte extremes (Table 4).

**Table 4.** List of analytes with significant cluster-wise differences (padj ≤ 0.05) and corresponding Cohen’s d effect sizes across the 12-cluster solution. *Cluster-wise neurochemical features identified by effect-size–based enrichment analysis in the 12-cluster solution.* For each cluster, analytes differing significantly between each cluster and the remainder of the cohort after false discovery rate (FDR) correction (padj ≤ 0.05) are listed along with the direction and magnitude of Cohen’s *d*. Effect sizes were computed on age- and sex-adjusted, z-scored concentrations. Positive and negative values indicate relative elevation or reduction within the cluster, respectively. Only analytes meeting the FDR threshold are shown.

| Cluster | Cluster size (n) | No. of FDR-Sig<br>nificant<br>Analytes | FDR Significant Analytes (Cohen's $d$ with Direction) |
| --- | --- | --- | --- |
| 1 | 23 | 12 | Phenethylamine [PEA] (−0.36); Tyrosine (−0.43); Tyramine (−0.64); Dopamine (−0.59); DOPAC (−0.47); Norepinephrine (−0.73); Normetanephine (+0.87); Serotonin (+0.68); GABA (−0.54); Glycine (−0.50); Histamine (+0.75); Taurine (−0.98) |
| 2 | 23 | 8 | PEA (−0.59); Tyramine (−0.89); Tyrosine (−0.43); Metanephine (−0.70); Norepinephrine (+0.80); Glycine (+1.45); Histamine (−0.91); GABA (−0.44) |
| 3 | 18 | 7 | Serotonin (−0.93); Epinephrine (−0.57); Histamine (+0.89); Norepinephrine (+1.33); Tyrosine (+1.14); Tyramine (−0.56); Taurine (−1.40) |
| 4 | 25 | 5 | Dopamine (+2.26); Epinephrine (+0.54); Glutamate (+0.85); Normetanephine (−0.60); Metanephine (−0.46) |
| 5 | 24 | 7 | Normetanephine (+1.32); Tyramine (+1.57); DOPAC (+0.86); PEA (−0.42); Histamine (−1.0); Epinephrine (−0.50); Glutamate (−0.53) |
| 6 | 22 | 7 | Metanephine (+1.22); Dopamine (+0.54); Glutamate (−1.17); Norepinephrine (−1.17); Epinephrine (−0.57); Normetanephine (−0.54); 3-Methoxytyramine [3-MT](−0.64) |
| 7 | 15 | 11 | PEA (+2.19); 3-MT (+0.85); Tyrosine (−0.56); DOPAC (−0.92); Metanephine (−0.91); Glutamate (−0.94); Histamine (−0.87); Dopamine (−0.57); Epinephrine (−0.74); Serotonin (−0.85); Norepinephrine (−0.60) |
| 8 | 22 | 7 | Tyramine (+0.65); Taurine (+0.70); Histamine (+0.44); Serotonin (−0.86); Metanephine (−0.84); Tyrosine (−0.50); Norepinephrine (−0.55) |
| 9 | 14 | 2 | PEA (+2.78); Tyrosine (+1.85) |
| 10 | 23 | 7 | GABA (+2.56); DOPAC (+0.71); 3-MT (+0.75); Serotonin (+0.49); Glutamate (−0.54); Glycine (−0.56); Histamine (−0.55) |
| 11 | 20 | 5 | DOPAC (+0.83); Glycine (+0.74); Metanephine (+0.75); Normetanephine (−0.71); Serotonin (−0.50) |
| 12 | 32 | 11 | PEA (−0.69); Tyrosine (−0.74); Dopamine (−0.98); DOPAC (+0.47); Epinephrine (+1.22); Metanephine (+0.62); Serotonin (+1.07); Glutamate (+1.08); GABA (−0.53); Histamine (+0.89); Taurine (+0.63) |

Hierarchical clustering of the Cohen’s d matrix produced coherent blocks of analytes that tended to co-vary across clusters. These blocks corresponded to biologically meaningful pathways

- a trace-amine block (PEA, tyramine),
- a catecholamine metabolism block (DOPAC, 3-MT, metanephrine, normetanephrine),
- a serotonergic block (serotonin),
- an excitatory–inhibitory amino acid block (glutamate, GABA, glycine, taurine).

This organisation indicates that clusters were defined by coordinated multi-analyte profiles, not random variation.

Together, the statistical enrichment analysis shows that each cluster is characterised by a consistent and reproducible pattern of analyte shifts, providing a quantitative foundation for the biological interpretations presented below.

These quantitative patterns provided the basis for deriving pathway-level biochemical fingerprints for each subtype. Although Gaussian mixture modelling indicated that the biochemical space contains stable cluster cores embedded within an overall continuous structure, each of the 12 clusters displayed a coherent and biologically interpretable multi-analyte fingerprint. These fingerprints consisted of shifts across trace amines, monoamines, catecholamine metabolites, excitatory–inhibitory amino acids, and histamine-related markers.

The effect-size patterns across the 12 clusters revealed simultaneous deviations across multiple neurotransmitter pathways rather than isolated single-analyte shifts. Trace-amine alterations were among the strongest signals, with markedly elevated phenethylamine defining Cluster 7 and positive tyramine characterising Cluster 5. Several clusters exhibited distinctive catecholamine-turnover profiles: Clusters 5, 10, and 11 showed increased DOPAC and 3-MT, whereas Cluster 6 showed reduced metanephrine, normetanephrine, and 3-MT, indicating reduced adrenergic profile. Excitatory–inhibitory balance also varied, with Cluster 10 showing combined elevations in GABA and serotonin, while Clusters 1, 2, and 12 showed reduced GABA. Histamine displayed both elevations (Clusters 1, 3, 12) and reductions (Clusters 5, 7, 10), often co-occurring with serotonergic or catecholaminergic shifts. Serotonin showed bidirectional changes, with notable elevations in Clusters 10 and 12 and reductions in Clusters 7 and 8.

A structured summary of the subtype fingerprints is presented in Table 5, including the dominant analyte elevations/depletions, pathway-level tendencies, and their corresponding effect sizes. Many clusters displayed combinations that partially resemble previously reported ASD biochemical patterns (e.g., serotonergic imbalance, sympathetic overactivation, histamine-linked irritability), but the majority represented novel integrated multivariate biochemical patterns detectable only through multivariate profiling rather than single-analyte comparisons.

**Table 5.**
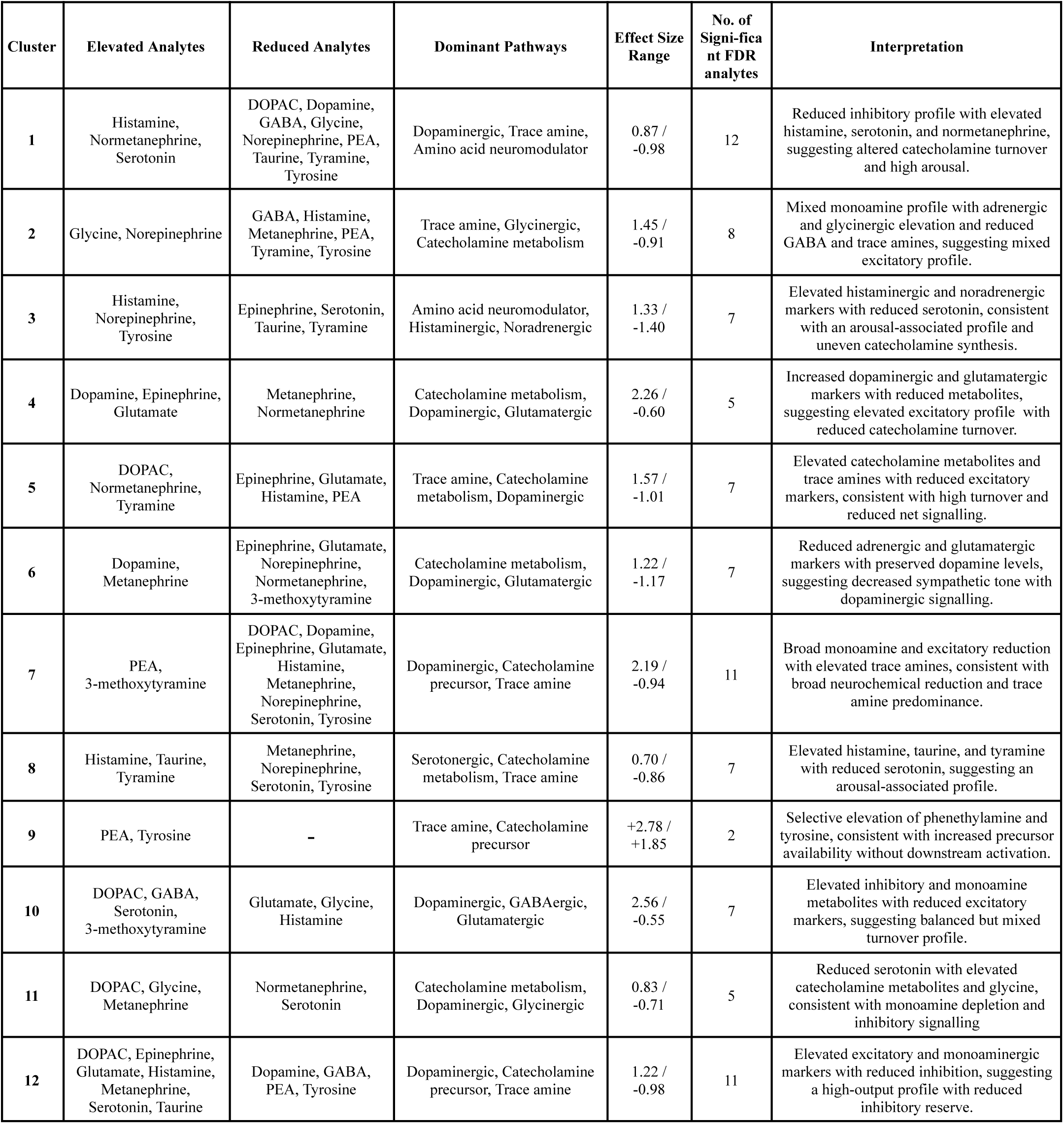
Biochemical fingerprints of the 12-cluster solution (k=12) *Summary of cluster-specific biochemical fingerprints identified in the 12-cluster (k=12) solution.* For each cluster, analytes showing significant cluster-versus-remainder differences after false discovery rate correction (padj ≤ 0.05) were grouped into elevated and reduced sets based on the direction of Cohen’s *d*. Dominant pathways represent pathway-level interpretations derived from the collective pattern of significant analytes rather than single-marker effects. Effect size ranges indicate the maximum positive and negative Cohen’s *d* observed within each cluster. Interpretations reflect integrated neurochemical profiles and are intended as descriptive summaries of cluster-level patterns.

These results show that, despite being within a continuous multivariate manifold, ASD blood neurotransmitter biology contains reproducible blocks of variation, biochemical “directions” or “tendencies” that define distinguishable ‘biochemical modes’ in terms of pathway activation, metabolic load, and excitatory-inhibitory tone. These fingerprint profiles form the biochemical substrate for subsequent clinical correlation analyses linking these clusters to behavioural and developmental phenotypes. Crucially, the biochemical fingerprints identified here are defined not by single analyte deviations, but by reproducible combinations of opposing and coordinated shifts across multiple neurotransmitter systems, a pattern not observed in prior single-analyte or case–control studies in ASD.

To evaluate whether higher-resolution subtyping produced more refined or more fragmented patterns, we repeated the enrichment analysis for the k=15 solution. Increasing k beyond 12 generated smaller and less stable clusters, and the effect-size patterns became more diffuse and fragmented across a larger number of groups. Although several k=15 subclusters preserved components of the k=12 biochemical fingerprints, these signals were split and redistributed across multiple smaller clusters, without revealing any new or more interpretable axes of variation. This behaviour was consistent with the decreasing stability observed in the consensus clustering results. Accordingly, a full biochemical interpretation of the k=15 solution was not pursued, and these partitions were treated as a sensitivity analysis illustrating that the coherent profiles seen at k=12 become increasingly over-partitioned at higher resolutions, without meaningful gains in biological clarity.

### Clinical Correlations: Mapping Biochemical Clusters to Qualitative Symptoms

To examine whether the biochemical clusters corresponded to differences in behavioural presentation, we merged cluster assignments with therapist-reported qualitative symptom data, which included binary indicators of behavioural and sensory features and verbal/non-verbal communication status. After harmonizing patient identifiers and encoding qualitative responses into numeric form, symptom incidence was calculated for each cluster with at least one FDR-significant analyte in the enrichment analysis. The resulting incidence map (figure 10) provides descriptive context for the biochemical patterns. Formal statistical evaluation of symptom–cluster associations is presented separately below and in (Supplementary Table S7). Across all clusters, core ASD behaviours were highly prevalent: stereotypical behaviour (92–100%), hyperactivity (87–100%), sensory issues (88–100%), and attention difficulties (78–97%) [35]. These uniformly high rates indicate that no biochemical cluster was defined by the absence of core ASD traits; rather, clusters varied primarily in the degree of symptom expression, consistent with the continuous biochemical organisation supported by the clustering analyses.

**Figure 10.**
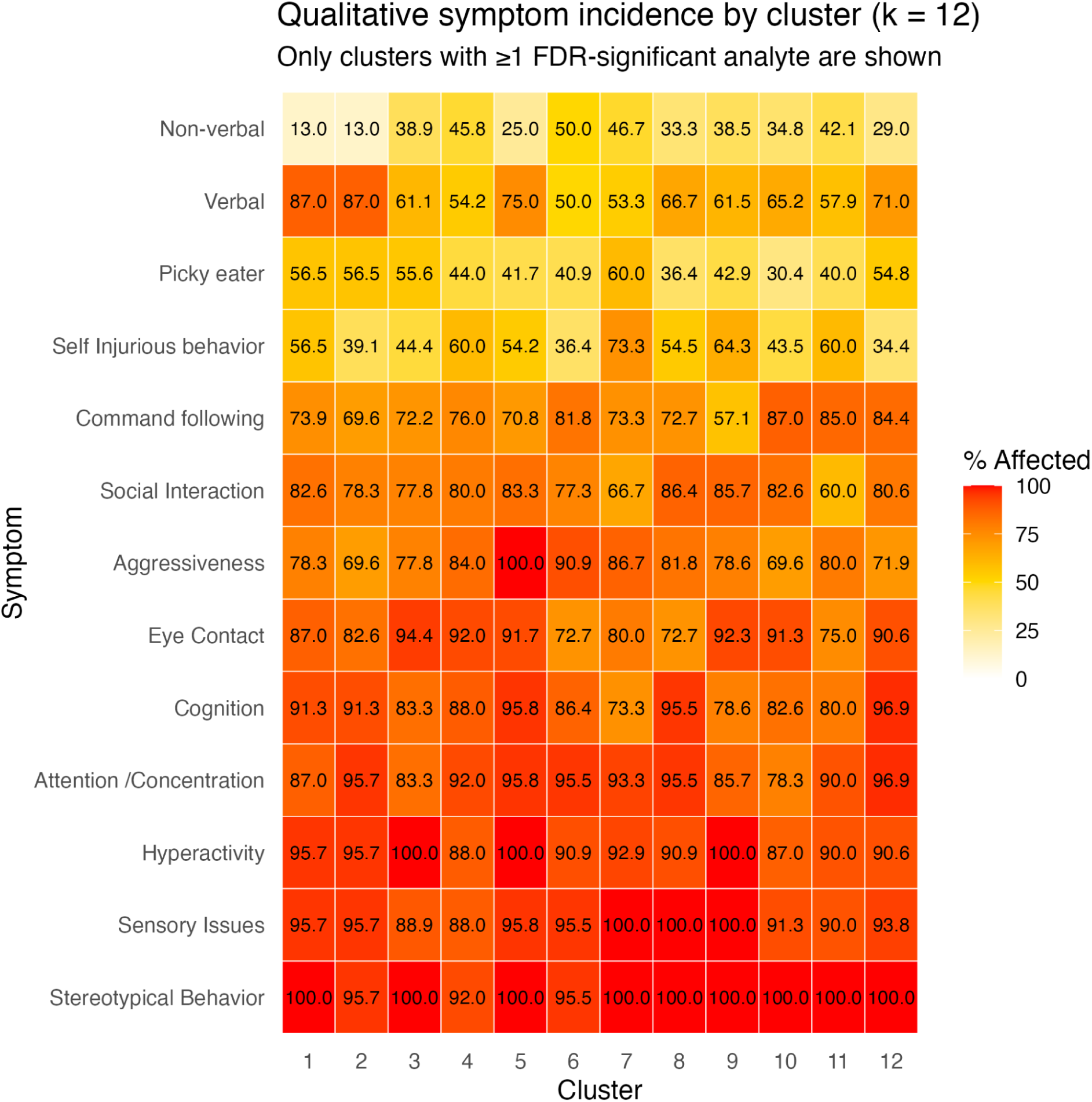
Qualitative symptom incidence across the 12 biochemical clusters. Cluster-wise incidence of qualitative clinical symptoms across the k=12 biochemical partition. Each cell represents the percentage of individuals within a cluster exhibiting the indicated symptom. Warmer colours indicate higher symptom prevalence. The heatmap provides descriptive behavioural context for the biochemical partition.

Within this overall uniformity, several symptoms nevertheless displayed appreciable numerical variation across clusters (Table 6). Aggressiveness ranged from 69% to 100% across clusters, self-injurious behaviour from 34% to 73%, picky eating from 30% to 60%, eye contact impairment from 73% to 94%, social interaction impairment from 60% to 86%, and non-verbal communication from 9% to 50%. No symptom was uniquely confined to any single cluster, and no cluster exhibited a singular behavioural phenotype.

**Table 6.**
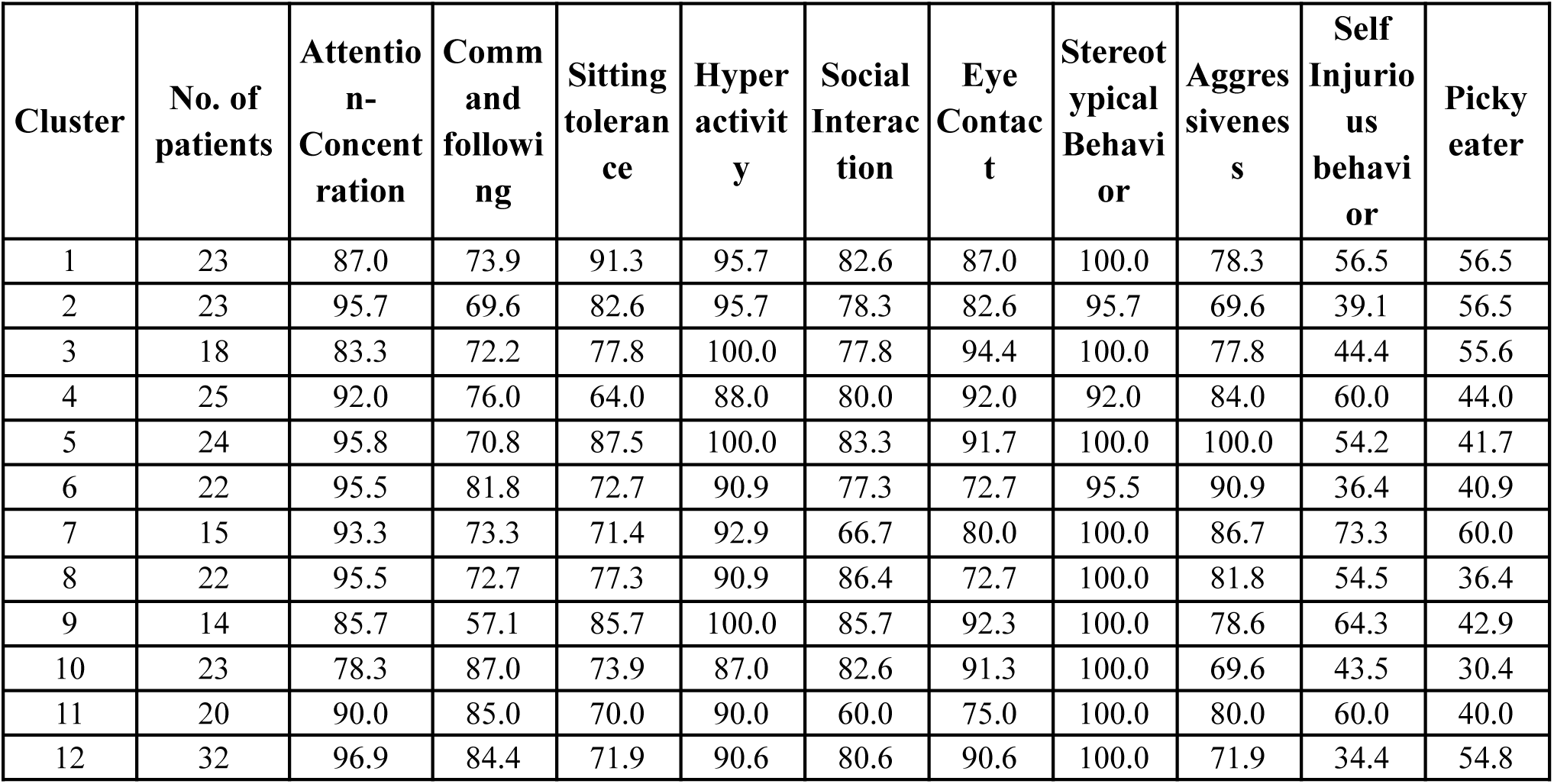
Selected Symptom Incidence Across Clusters. *Cluster-wise incidence of selected qualitative clinical features across the primary k=12 biochemical partition.* A complete symptom-by-cluster incidence table, including verbal status and all assessed symptoms, is provided in Supplementary Table S6.

### Formal testing of symptom-cluster associations

To determine whether the observed differences in symptom prevalence represented statistically distinguishable behavioural profiles, associations between cluster membership and each qualitative symptom were evaluated using Pearson’s chi-square test or Monte Carlo chi-square testing when expected cell counts violated asymptotic assumptions, with Benjamini–Hochberg correction applied across all tested symptoms (Supplementary Table S7).

None of the evaluated symptoms differed significantly across clusters after correction for multiple comparisons (adjusted p = 0.77-0.81). The smallest raw p-value was observed for verbal status (χ² = 16.3, p = 0.131), while aggressiveness exhibited one of the widest prevalence ranges (69–100% across clusters) despite a non-significant raw p-value (χ² = 13.1, p = 0.278). Several core ASD behaviours, including stereotypical behaviour (92-100%), hyperactivity (87–100%), and sensory issues (88-100%), were highly prevalent across all clusters, leaving little between-cluster variation available for statistical discrimination. Conversely, symptoms showing greater numerical variability including verbal status (9-50%), self-injurious behaviour (34-73%), and picky eating (30–60%) were evaluated within relatively small cluster sizes (n = 14-32), limiting statistical power. Overall, no categorical symptom uniquely distinguished the biochemical partition. These findings are consistent with the interpretation that the biochemical organisation identified by NeuroCLAD represents a continuous rather than symptom-defined structure, although larger cohorts and dimensional behavioural measures will be required to determine whether more subtle graded relationships are present.

Taken together, several neurotransmitter axes including catecholamine turnover, trace amines, histamine, and serotonin, appear to align with broad behavioural domains such as aggression, self-injury, feeding difficulties, and communication impairments. However, these associations are non-deterministic and, although formally evaluated in the present cohort, require replication and validation in larger, independent cohorts. The present findings should therefore be interpreted as exploratory descriptive patterns that support a structured continuum of biochemical variation rather than as evidence for symptom-defined biochemical modes within the cohort.

### Cross-Solution Correspondence Between k=12 and k=15 (Alluvial Mapping)

To assess how the primary 12-cluster biochemical structure reorganised under a higher-resolution partition, we examined correspondence between the k=12 and k=15 solutions using two complementary alluvial maps: one based on shared enriched analytes and the other based on shared patients [27, 28].

### Enrichment-based correspondence

The enrichment similarity map (Supplementary Figure S11A) did not reveal a clear or structured correspondence between the k=12 and k=15 solutions. Connections between clusters were diffuse and widely distributed, with no dominant ribbons indicating strong preservation of enrichment signatures. This suggests that the pathway-level biochemical patterns identified at k=12 do not map cleanly onto the higher-resolution k=15 partition.

Rather than reflecting structured subdivision, the enrichment patterns appear to disperse across multiple clusters at k=15, resulting in a loss of coherent biochemical signal. No k=15 cluster exhibited a uniquely strong or interpretable enrichment overlap with any single k=12 cluster, and no new dominant biochemical axes emerged at higher resolution. This indicates that increasing granularity disrupts the pathway-level organisation captured at k=12 rather than refining it.

### Sample-based correspondence

The patient-flow alluvial map (Supplementary Figure S11B) showed a similar pattern. Most ribbons connecting k=12 to k=15 clusters were moderate in width, indicating that individual k=12 clusters were partitioned into several smaller groups at k=15. A small number of k=15 clusters (e.g., k15_8, k15_13) accumulated larger patient flows from multiple k=12 clusters, indicating that some higher-resolution clusters represented mixed reorganisations rather than clean subdivisions. No one-to-one correspondence was observed between any pair of clusters, and no k=15 cluster cleanly absorbed the majority of individuals from a single k=12 group.

Together, the enrichment-based and sample-based alluvial mappings provide complementary perspectives on cluster stability. While the sample-based mapping demonstrates structured subdivision of patient groups at higher resolution, the enrichment-based mapping reveals a loss of coherent biochemical correspondence. This combination suggests that the k=15 solution fragments the biologically meaningful structure identified at k=12 without introducing clearer or more interpretable axes of variation. The k=15 solution thus functions as a sensitivity check, confirming that the dominant biochemical patterns are best captured at the 12-cluster scale [25, 26].

### Supportive Cross-Compartment Urine Analysis

Applying NeuroCLAD to urine neurotransmitter profiles from the same cohort produced stable multivariate structure, with consensus clustering at k=12 yielding a PAC of 0.256, comparable in magnitude to the blood-derived solution (Supplementary Figure S12A). Effect-size patterns for analytes shared between the two fluids showed convergent module-level shifts in catecholaminergic turnover, serotonergic metabolism, and excitatory–inhibitory amino acid balance (Supplementary Figure S12B).

Direct cluster-level concordance between blood and urine was not pursued in the present analysis, given known limitations in the urine sampling protocol, including variable collection times across the day and the use of mixed sample types. These analyses are therefore presented as supportive cross-compartment context rather than as independent validation of the blood-derived clustering structure.

## Discussion

Autism Spectrum Disorder (ASD) is widely appreciated as a heterogeneous neurodevelopmental condition, but the extent and structure of its biochemical heterogeneity remain only partially understood [2, 4]. Prior studies examining neurotransmitters and related metabolites in ASD have often relied on single-analyte comparisons or categorical case–control contrasts, approaches that are not well suited to capturing multidimensional biological organisation [16, 17, 45]. The present study applied an integrated multivariate framework to blood neurotransmitter profiles in a large, clinically characterised ASD cohort, combining age-adjusted dimensionality reduction, consensus clustering, Gaussian mixture modelling, pathway-level enrichment, and complementary null-model validation. Taken together, these analyses reveal a reproducible biochemical landscape characterised by structured modes of variation embedded within a continuously overlapping spectrum. These findings support a view of ASD neurochemistry as a structured continuum rather than a collection of discretely separated biochemical categories, consistent with contemporary dimensional models of ASD [3, 4].

Recent work has begun to explore multivariate biochemical structure within ASD. For example, Ferraro et al. [26], demonstrated that coordinated amino acid patterns can reveal latent grouping within ASD populations using dimensionality reduction and clustering approaches. The present study extends this concept by applying a more comprehensive, layered analytical framework to neurotransmitter systems, incorporating stability assessment, probabilistic membership modelling, and pathway-level interpretation to characterise systems-level biochemical organisation.

A critical prerequisite for multivariate analysis was the removal of demographic structure. Multiple neurotransmitters displayed nonlinear age-related trajectories in the raw measurements. GABA provided one of the clearest examples, with tightly constrained concentrations during early childhood becoming progressively more dispersed with increasing age, producing a curved developmental trajectory [17, 24]. In datasets of this size, such curvature can dominate multivariate decompositions, creating spurious principal components and artificially inflating apparent subgroup structure [29, 30]. The spline-based adjustment effectively removed these systematic demographic gradients, leaving behind the inter-individual biochemical variance more plausibly attributed to neurobiological differences [30]. The age-adjusted matrix therefore formed the basis of all subsequent unsupervised analyses, ensuring that the recovered biochemical structure reflected inter-individual variation rather than systematic differences in age or sex.

The PCA step revealed a distributed multivariate signal rather than one dominated by a small number of analytes [31]. Variance declined gradually across components without a distinct elbow, indicating that neurotransmitter variation in ASD arises from the combined contribution of many biochemical processes rather than a single dominant axis [29]. The first ten principal components captured approximately 70% of the total variance and showed no residual association with age or sex, confirming effective demographic adjustment [30]. These components provided a compact representation of the biochemical landscape while preserving the multivariate structure required for downstream clustering analyses.

K-means clustering across a range of resolutions demonstrated the expected behaviour of a system that contains structure but resists categorisation into a few sharply separated partitions [32]. The elbow curve did not identify a natural breakpoint, and the silhouette curve improved only gradually [39], both hallmarks of an underlying continuum rather than a sharply partitioned latent structure [4, 28]. Nonetheless, centroid heatmaps across candidate k values, particularly k=12 and k=15, revealed coherent motifs: trace-amine shifts, catecholamine turnover differences, serotonergic modulation, and excitatory–inhibitory imbalances [8, 25, 46, 47]. These motifs appeared consistently and were interpretable in terms of known biochemical pathways, suggesting genuine biological signal rather than mathematical artifacts [16, 48]. Among the candidate resolutions, the k=12 solution provided the best balance between granularity, stability, and biochemical interpretability. Although k=15 further subdivided the data, it did not reveal additional coherent pathway-level organisation, instead fragmenting existing biochemical signatures across smaller clusters. Accordingly, k=12 was retained as the primary analytical resolution, while k=15 was carried forward as a higher-resolution sensitivity analysis to evaluate the stability of the identified biochemical organisation.

Consensus clustering provided an independent assessment of partition stability, allowing the reproducibility of the identified biochemical structure to be evaluated across repeated bootstrap resampling [27]. Across bootstrap iterations, the k=12 solution exhibited well-defined diagonal blocks in the consensus matrix, confirming reproducible co-clustering of individuals [27]. Some boundary diffusion was evident in a subset of clusters, reflecting modest internal heterogeneity. Higher-k solutions produced visually sharper matrices, but this sharpness resulted from fragmentation into small, unstable clusters rather than improved biological separation [28]. The combined behaviour of PAC, mean within-cluster consensus, and cluster-size stability all converged on the same conclusion: k=12 captured the most coherent structure before instability dominated. This observation meant the choice of k=12 supported not by a single optimization criterion, but by the convergence of multiple complementary lines of evidence, including consensus stability, biochemical interpretability, probabilistic modelling, and subsequent null-model validation [28].

A reasonable question at this point is whether the observed clustering metrics are sufficient to support the proposed biological interpretation. Although the silhouette values (0.10–0.12) and mean within-cluster consensus (0.42 at k=12) are modest compared with those typically reported for cancer subtyping studies, this difference is biologically informative rather than unexpected. Cancer subtypes arise from clonal expansion of shared driver mutations [51], so sharply separated clusters are exactly what one would expect [50, 51], ASD has no comparable generative mechanism. Its biology is polygenic, multifactorial, and shaped by developmental processes [5, 51], making diffuse rather than discrete organisation biologically plausible. Importantly, the observed k=12 consensus solution exceeded both isotropic Gaussian and covariance-preserving null models, demonstrating that the recovered structure cannot be explained either by random variation or by the global covariance structure of the analytes alone. The distinction between these null models is informative. The isotropic null tests whether clustering exceeds structureless noise, whereas the covariance-preserving null asks the more stringent question of whether the observed organisation exceeds what would be expected from a correlated multivariate cloud with the same covariance structure. The k=12 solution exceeded both null models, indicating that the recovered biochemical organisation cannot be explained either by random variation or by the covariance structure of the analytes alone.

Gaussian mixture modelling provided a complementary probabilistic perspective on the clustering structure. While k-means identifies discrete partitions through hard assignment, GMM enables soft assignment by modelling the data as a mixture of underlying probability distributions and estimating the degree of membership of each individual across clusters [33, 34]. Together, these approaches allow the biochemical landscape to be viewed both as an operational partition and as a probabilistic continuum, providing complementary perspectives on the underlying organisation. In the present data, the distribution of maximum posterior probabilities was strongly skewed toward higher values, indicating that a large proportion of individuals are assigned to clusters with high confidence. This indicates the presence of reproducible biochemical modes that act as centres of organisation within the broader biochemical landscape.

At the same time, a subset of individuals exhibited lower posterior probabilities and higher entropy values, reflecting partial membership across multiple clusters. These individuals occupied transitional regions between neighbouring biochemical modes, indicating that while the modes were reproducible and structurally coherent, their boundaries were gradual rather than sharply discrete [4]. Rather than indicating an absence of organisation, this probabilistic overlap suggests that the identified modes are connected by continuous transitions, consistent with the broader multidimensional organisation of ASD neurochemistry [4, 51].

While k-means and consensus clustering evaluated the stability of an imposed clustering resolution, model-based enumeration addressed a complementary question: how many latent mixture components are supported without fixing the number of clusters in advance. Rather than contradicting the k=12 solution, this analysis indicated that the dominant biochemical variation is captured by a small number of broad mixture components, with finer biochemical modes emerging only when the continuum is partitioned at an operational analytical resolution.

The observed organisation is therefore neither fully discrete nor completely diffuse. An intuitive way to understand it is through the behaviour of a non-Newtonian fluid, whose observed behaviour changes depending on how it is examined, yet the material itself remains unchanged. When a fixed clustering resolution is applied, reproducible biochemical modes emerge that are sufficiently coherent to permit stable assignment and biological interpretation. At the same time, probabilistic modelling reveals gradual transitions between neighbouring modes rather than rigid boundaries. The modes are therefore genuine features of the underlying data, but they are embedded within a continuous biochemical landscape rather than existing as isolated categories. This interpretation is further supported by the low-dimensional visualisation (Supplementary Figure S4), in which the biochemical modes occupy partially overlapping regions rather than forming discrete islands, consistent with a structured continuum of neurochemical variation [4, 33, 34]

Cohen’s d enrichment analysis provided the biochemical grounding for the cluster interpretations [42]. Rather than being driven by single analytes, each cluster exhibited coordinated shifts across multiple neurotransmitter systems, with several effect sizes reaching large or very large magnitudes, particularly among the trace amines [17, 45]. These alterations organised into recurrent pathway-level patterns involving trace amine signalling, catecholamine turnover, serotonergic modulation, histaminergic pathways, and excitatory-inhibitory amino acid balance [8, 17, 19, 24, 25, 46, 47]. The reproducible recurrence of these pathway combinations across multiple clusters indicates that the k=12 solution captures biologically coherent modes of neurochemical variation rather than arbitrary statistical partitions [51].

Several enrichment profiles were particularly noteworthy. Phenethylamine and tyramine primarily act through trace amine-associated receptor 1 (TAAR1), an important regulator of dopaminergic, serotonergic and noradrenergic neurotransmission [52]. Cluster 7 was characterised by elevated phenethylamine together with broadly reduced monoamines, a pattern consistent with altered trace amine regulation, although peripheral blood measurements do not permit direct inference regarding central TAAR1 activity. By contrast, Cluster 9 exhibited elevated phenethylamine and tyrosine without comparable downstream monoamine depletion, potentially representing a more proximal precursor accumulation state and suggesting heterogeneity within trace amine dysregulation itself.

Interestingly, this trace amine signature was also the only biochemical axis independently recovered when the number of mixture components was selected automatically using the Bayesian Information Criterion. Rather than resolving twelve latent biochemical classes, model-based enumeration favoured a two-component solution, in which a smaller subgroup (n = 39) separated from the larger cohort (n = 222) primarily along the phenethylamine–tyrosine axis (Supplementary Table S10) [33, 34]. This convergence between enrichment analysis and model-based enumeration identifies trace amine biology as one of the strongest sources of neurochemical variation within the cohort while further supporting the interpretation that the dominant organisation of the data reflects continuous biochemical variation rather than multiple discrete biochemical classes [4, 33, 34, 52].

Histamine enrichment also emerged as a recurrent feature across several clusters. Beyond its established immunological roles, histamine functions as a central neuromodulator regulating arousal, sensory processing and monoaminergic neurotransmission through H1, H2 and H3 receptor systems [19, 20]. Histamine elevation recurred alongside catecholaminergic and serotonergic alterations in Clusters 1, 3, 8 and 12, suggesting coordinated perturbation of interconnected neurotransmitter pathways rather than isolated analyte abnormalities. Although these clusters also displayed relatively higher incidences of sensory issues and picky eating, the absence of significant symptom-cluster associations precludes direct behavioural inference, and these biochemical-behavioural correspondences should be regarded as hypothesis-generating observations for future studies.

Several biochemical patterns resembled previously reported ASD alterations, including serotonergic imbalance, histaminergic modulation, and catecholaminergic dysregulation, whereas most represented novel integrated neurochemical signatures that emerged only through multivariate analysis [16, 17, 19, 24]. Together, these enrichment profiles indicate reproducible biochemical organisation that is not evident from single-analyte analyses alone.

The k=15 solution served as a higher-resolution sensitivity analysis. Although additional analytes reached statistical significance, the resulting effect-size patterns became increasingly fragmented, and no new biochemical axes emerged [28]. Probabilistic modelling likewise demonstrated greater boundary uncertainty despite retention of several high-confidence modes (Supplementary Figure S10), while pathway-level coherence was reduced compared with k=12. Together, these findings indicate that increased partitioning primarily subdivided existing biochemical organisation rather than revealing additional biologically meaningful structure, supporting k=12 as the most interpretable analytical resolution [27, 28].

Alluvial mapping further illustrated this behaviour. The sample-level mapping demonstrated that increasing resolution primarily subdivided existing patient groups. In contrast, the enrichment-based mapping showed that the biochemical signatures identified at k=12 did not map consistently onto the higher-resolution solution, indicating that additional partitioning reorganised rather than systematically refined the original biochemical modules (Supplementary Figures S11A-B). This observation supports the interpretation that the additional clusters primarily represent alternative subdivisions of the same biochemical landscape rather than the emergence of new, clearly distinguishable biochemical organisation.

Clinical symptom mapping provided an external behavioural context for the biochemical findings [2]. Core ASD features, including stereotypical behaviour, hyperactivity, sensory issues, and attention difficulties, were highly prevalent across all biochemical modes, consistent with the overall clinical profile of the cohort [4, 35]. Although several symptoms displayed appreciable numerical variation across clusters, formal statistical testing found no symptom to differ significantly after correction for multiple comparisons (Supplementary Table S7). These findings suggest that the biochemical organisation identified by NeuroCLAD is not reflected in discrete categorical behavioural profiles. One possible explanation is that current qualitative symptoms and conventional severity scales partition behaviour into broad categories that may not adequately capture continuous neurobiological variation [3, 4]. Future work will therefore investigate clinically derived multidimensional domains designed to better represent the underlying behavioural architecture of ASD.

These findings highlight a broader challenge in ASD research. Biological measurements, such as neurotransmitter profiles, capture continuous, multidimensional variation, whereas commonly used clinical instruments and diagnostic frameworks, including DSM-5, largely summarise ASD in terms of behavioural symptoms and semi-quantitative severity measures rather than underlying biological organisation [1]. This difference in resolution makes robust biomarker-behaviour relationships inherently difficult to detect. The present findings reflect this disconnect: reproducible biochemical organisation was not accompanied by discrete categorical behavioural profiles. Similar challenges have been reported in neuroimaging studies, where biological heterogeneity frequently exceeds the resolution of current clinical phenotyping frameworks [13, 14].

The identification of reproducible biochemical modes has important implications for future precision medicine approaches in ASD. Rather than assigning individuals to rigid biochemical subtypes, NeuroCLAD defines reproducible positions within a multidimensional neurochemical landscape shaped by coordinated variation across trace amines, monoamines, catecholamine turnover, histamine signalling, and excitatory-inhibitory balance [8, 25, 46, 47]. From this perspective, interventions targeting specific neurotransmitter systems may not be expected to produce uniform responses across individuals [16]. Although these implications remain hypothesis-generating and require prospective validation, the present findings provide a biologically grounded rationale for pathway-aware, individualized therapeutic strategies [4].

The implications of this framework extend beyond the present study. Rather than dividing ASD into mutually exclusive biochemical subgroups, future work may be better served by quantifying each individual’s position across multiple interacting neurochemical axes [51]. Such an approach aligns naturally with the spectrum concept of ASD [2]. NeuroCLAD provides a foundation for this direction by defining reproducible biochemical modes within a continuous landscape while offering an analytical framework that is readily extensible to CSF, urine, and other biological compartments.

Applying the same analytical framework to urine neurotransmitter measurements yielded comparable multivariate organisation, with broadly similar catecholaminergic, serotonergic, and excitatory-inhibitory patterns. Interpretation remains cautious because of variability in sampling conditions, including the absence of standardised collection protocols. These findings should therefore be regarded as supportive cross-compartment context rather than independent validation. Nevertheless, they demonstrate that the NeuroCLAD framework can recover biologically interpretable organisation across distinct biological compartments despite differences in their underlying physiology and sampling characteristics.

In summary, the present study demonstrates that ASD blood neurotransmitter profiles are neither homogeneous nor discretely partitioned, but are organised into reproducible biochemical modes embedded within a continuous multidimensional landscape [2, 4]. By integrating demographic adjustment, multivariate dimensionality reduction, consensus clustering, probabilistic modelling, enrichment analysis, and null-model validation, NeuroCLAD provides a reproducible framework for characterising systemic neurochemical heterogeneity in ASD. These findings establish a foundation for future integration of multidimensional biochemical and clinical-domain phenotyping across biological compartments.

## Limitations

### Several limitations of this study should be acknowledged

First, all analytes were measured in peripheral blood, which reflects systemic neurochemical metabolism rather than direct central neurotransmission. Although several analytes, particularly catecholamines and trace amines, have plausible links to central neurobiology, peripheral concentrations are also influenced by physiological processes outside the central nervous system. Accordingly, the present findings should be interpreted as describing systemic neurochemical organisation rather than direct brain neurochemistry.

Second, the NeuroCLAD framework is designed to characterise heterogeneity within the ASD population rather than to perform case–control comparisons. Clustering was performed on within-cohort z-scores, and cluster-level descriptors (e.g., “elevated dopamine”) therefore reflect relative positioning within the ASD distribution rather than deviation from neurotypical norms. A directly matched neurotypical comparison cohort was not available.

Third, the study is cross-sectional, limiting inference regarding the temporal stability and developmental evolution of the identified biochemical modes. Longitudinal studies will be required to determine whether these represent stable phenotypes or dynamic states.

Fourth, associations between biochemical modes and categorical clinical symptoms were formally tested but none survived correction for multiple comparisons. This null finding likely reflects both limited variance in several highly prevalent symptoms and the restricted resolution of categorical behavioural scales, together with relatively small per-cluster sample sizes (n = 14–32). Larger cohorts incorporating multidimensional behavioural domains may provide greater sensitivity for detecting graded neurochemical–clinical relationships.

Fifth, although internal stability analyses support the presence of reproducible structure, external validation in an independent blood cohort was not performed. Cross-fluid comparison using urine measurements provides supportive context but does not constitute formal replication, and is itself subject to variability in sampling conditions.

Sixth, the absolute clustering metrics (silhouette ∼0.10–0.12; mean within-cluster consensus ∼0.42 at k=12) are modest compared with studies of discretely partitioned biological systems. However, these values exceeded both isotropic Gaussian and covariance-preserving null models, indicating that the recovered organisation cannot be explained by random variation or covariance structure alone. The modest metrics therefore reflect the continuous nature of the underlying biological organisation rather than analytical instability.

Finally, the biochemical modes identified here represent structured patterns of variation rather than direct mechanistic pathways. Further experimental and clinical studies will be required to establish causal relationships and therapeutic relevance.

## Conclusion

In conclusion, this study demonstrates that blood neurotransmitter biology in Autism Spectrum Disorder is neither uniform nor discretely partitioned but organised into reproducible biochemical modes embedded within a continuous multivariate landscape. By applying the NeuroCLAD framework to a large, clinically characterised cohort, we identified novel, pathway-level biochemical fingerprints that reflect coordinated variation across trace amines, monoamines, catecholamine turnover, histamine signalling, and excitatory–inhibitory balance. These fingerprints help reconcile the long-standing inconsistency of previous neurotransmitter studies by showing that many apparently conflicting findings arise from structured biological heterogeneity rather than noise or methodological failure. Importantly, the results support a dimensional view of ASD biochemistry, in which stable modes of variation coexist with substantial overlap, rather than a model based on rigid categorical subtypes. Together with complementary null-model validation and model-based enumeration, these findings show that reproducible biochemical modes can emerge within a continuous neurochemical landscape without implying the existence of discrete latent biochemical classes.

Rather than relying solely on conventional descriptive behavioural categories, future stratification strategies may benefit from integrating quantitative biological axes with multidimensional clinical phenotyping, enabling individuals to be characterised by their position within a reproducible neurochemical landscape.

Together, these findings establish NeuroCLAD as a reproducible analytical framework for investigating neurochemical heterogeneity in ASD and provide a foundation for integrating biochemical profiling with multidimensional clinical phenotyping and cross-compartment analyses in future studies. More broadly, the study illustrates how multivariate systems-level approaches can reveal biologically meaningful organisation that remains obscured when complex neurodevelopmental disorders are viewed through single-analyte or categorical frameworks.

## Declarations

### Ethics approval and consent to participate

The study protocol was approved by the Central Drugs Standard Control Organization (CDSCO)-registered Institutional Ethics Committee (IEC) of NeuroGen Brain and Spine Institute, Navi Mumbai, India (Approval No.: NGBSI/IEC/AT-INV-01/2025/ISSUE-01/REVISION-01). The study was conducted in accordance with the ethical principles of the Declaration of Helsinki and applicable national ethical and regulatory guidelines. Written informed consent was obtained from the parents or legal guardians of all participants included in the study. Clinical trial number is not applicable.

### Consent for publication

Not applicable

### Availability of data and materials

The datasets generated and/or analysed during the current study are not publicly available because they contain participant-level clinical information that may remain potentially re-identifiable and are subject to institutional confidentiality requirements. They are available from the corresponding author upon reasonable request and with permission from NeuroGen Brain and Spine Institute.

The code used for data processing, clustering, and analysis is publicly available at: https://github.com/georgevinnit/NeuroCLAD. All scripts are provided to support reproducibility of the analytical pipeline.

### Authors’ contributions

V.G., H.S., and A.S. contributed to the conceptualization of the study. Methodology development, formal analysis, data curation, visualization, and original draft preparation were carried out by V.G. Pipeline conceptualization and coding were performed by V.G. and S.T. Validation was conducted by V.G. and H.S. Resources were provided by A.S., P.K., and H.S. Review and editing of the manuscript were undertaken by V.G., H.S., P.K., N.G., P.B., S.T., and A.S. Supervision was provided by H.S., P.K., and A.S., while project administration was managed by P.K. Overall study oversight was led by A.S. All authors read and approved the final manuscript.

## Acknowledgements

The authors thank Mr. Sudarshan Daga for assistance with data entry, and Ms. Namrata Trivedi and Ms. Tahreem Hamdule for their support as part of the research team. The authors acknowledge the use of AI-assisted tools (ChatGPT, OpenAI, and Claude [Anthropic]) for code development and limited language editing. All study design, analytical decisions, statistical analyses, interpretation of results, and final manuscript content were conceived, reviewed, verified, and approved by the authors.

## Authors’ information

Alok Sharma is Director and Neurosurgeon at NeuroGen Brain and Spine Institute, Navi Mumbai, India. Vinnit George is a Research Associate at the same institute within the Department of Research and Development. Hemangi Sane is Deputy Director, Head of Research and Development, and Consultant Physician. Nandini Gokulchandran is Deputy Director, Head of Medical Services, and Consultant in Regenerative Medicine. Pooja Kulkarni is Chief Research Officer and Head of Regulatory Affairs. Siddhi Talgaonkar is a Research Associate in the Department of Research and Development. Prerna Badhe is Deputy Director, Head of Regenerative Laboratory Services, and Consultant Neuropathologist.

