## Supplementary Appendix for "Reproducible Biochemical Modes Within a Structured Neurochemical Continuum in Autism Spectrum Disorder Revealed by NeuroCLAD"

#### Supplementary Methods

##### 1. Data Quality Control and Pre-processing

All analyses were performed in R (version 4.5.2). Packages included: *tidyverse*, *cluster*, *mclust*, *uwot*, and *pheatmap*.
Analytes with >15% missingness were designated for exclusion, and samples missing >25% of analytes were designated for removal. No analytes or samples exceeded these thresholds, and no residual missing values required imputation. The complete dataset was therefore carried forward for covariate adjustment and dimensionality reduction.

##### 2. Covariate Adjustment

Natural cubic spline regression (3 degrees of freedom) was used to model non-linear age effects for each analyte, with sex included as a fixed-effect factor.

Residuals from each model constituted the covariate-adjusted dataset.

##### 3. Principal Component Analysis

PCA was performed on the residualised, z-scored analyte matrix using *prcomp* with default SVD. No additional scaling was applied. Components explaining ~70% of cumulative variance (~PC1–PC10) were retained for clustering.

##### 4. k-means Clustering

k-Means was applied to the retained PCs using Euclidean distance and 50 random starts. Candidate k values ranged from k=2 to k=20. Model evaluation used within-cluster sum of squares (WCSS), silhouette width, cluster size distributions, and centroid interpretability. A fixed random seed was used to ensure reproducibility.

##### 5. Consensus Clustering

Cluster robustness was evaluated using 200 bootstrap iterations per k. Each iteration sampled 80% of individuals and 80% of PCs without replacement. A consensus matrix was computed as the proportion of times each pair of individuals co-clustered. Stability was summarised using mean within-cluster consensus and the Proportion of Ambiguous Clustering (PAC; 0.1–0.9 thresholds). k-means clustering was applied within each resampling iteration.

**6. Null-model validation**

Two null ensembles (200 datasets each) were generated in the retained principal-component space (261 participants × 10 principal components). One-sided empirical p-values were calculated as the proportion of null datasets producing statistics at least as extreme as the observed value. Because 200 null datasets were generated, the smallest attainable non-zero empirical p-value was 0.005; accordingly, results for which no null dataset was more extreme than the observed value were reported as $p < 0.005$. The isotropic null sampled principal-component scores independently from a standard normal distribution (N(0,1)). The covariance-preserving null sampled observations from a multivariate normal distribution parameterised by the observed principal-component mean vector and covariance matrix. Each null dataset was analysed using the identical consensus clustering workflow as the observed data (k=12, 200 bootstrap resamples, 80% subsampling of participants and principal components, 50 random starts per bootstrap). The Proportion of Ambiguous Clustering (PAC; consensus interval 0.1–0.9) and mean within-cluster consensus were calculated for each null dataset. One-sided empirical p-values were obtained by comparing the observed statistics with the corresponding null distributions.

**7. Bayesian Information Criterion-guided Model Enumeration.**

Gaussian mixture models were fitted to the retained principal-component scores using the mclust package over G = 1–20 mixture components. Candidate covariance parameterisations included EEE (equal volume, equal shape, equal orientation) and VVV (variable volume, variable shape, variable orientation). Model selection was performed using the Bayesian Information Criterion (BIC), with higher (less negative) values indicating stronger support after penalisation for model complexity. The optimal model was subsequently characterised by component membership, demographic composition and correspondence with the consensus k=12 clustering solution.

**8. Gaussian Mixture Modelling (Soft Cluster Membership)**

Gaussian mixture modelling was fitted in the retained principal-component space at the operational clustering resolutions examined in the NeuroCLAD framework. For the primary k=12 analysis, posterior membership probabilities were computed for each individual. Maximum posterior probability was used to quantify assignment confidence, and Shannon entropy was calculated from the posterior probability vector to quantify membership uncertainty. This fixed-resolution analysis was used to characterise overlap within the operational k=12 partition and was distinct from the BIC-guided enumeration analysis used to estimate latent mixture complexity.

**9. UMAP Embedding**

UMAP was applied to the retained principal components (n_neighbors = 15, min_dist = 0.1, metric = "euclidean") using the *uwot* package in R. The embedding was used strictly for visualisation and did not influence clustering or model selection [53].

**10. Enrichment Analysis**

For each cluster, each analyte was compared to all other individuals using two-sample t-tests on z-scores. Cohen's d was used to quantify effect size and direction. FDR correction applied Benjamini–Hochberg ($q < 0.05$). Given the large sample size, t-tests were considered robust to moderate deviations from normality.

**11. Symptom Mapping**

Binary qualitative symptom variables and verbal/non-verbal status were harmonised and merged by patient ID. Cluster-wise incidence proportions were calculated descriptively, followed by symptom-wise contingency-table testing.

**12. Cross-Solution Correspondence**

Alluvial diagrams summarised mapping between k=12 and k=15 clusters using (a) shared analyte enrichments and (b) shared individual memberships.

**13. Supportive Cross-Compartment Urine Analysis**

The NeuroCLAD analytical pipeline was independently applied to the urine neurotransmitter dataset using the same preprocessing, dimensionality reduction, and clustering framework as the blood dataset. The resulting biochemical organization was compared with the blood-derived NeuroCLAD structure to evaluate the consistency of broad biochemical patterns across biological compartments. Owing to the non-standardized urine sampling protocol, including variable collection times and mixed sample types, these analyses were considered exploratory and are presented as supportive cross-compartment context rather than formal validation of the blood-derived clustering framework.

#### Supplementary Tables

**Supplementary Table S1. Analyte metadata and laboratory reference information**

| Analyte Name | Neurochemical Class | Measurement Unit | Laboratory Reference Range (Low-High) | Included in Multivariate Analysis | Notes |
|---|---|---|---|---|---|
| Phenethylamine | Trace amine | pg/ml | 10-60 | Yes | Trace amine |
| Tyrosine | Catecholamine precursor | umol/l | 30-120 | Yes | Neurotransmitter precursor |
| Tyramine | Trace amine | ug/ml | 0.019-0.398 | Yes | Trace amine |
| Dopamine | Catecholaminergic | pmol/l | 10-30 | Yes | Primary neurotransmitter |
| 3,4-Dihydroxyphenylacetic acid (DOPAC) | Catecholaminergic metabolite | ng/ml | 0.1-0.3 | Yes | Catecholamine metabolite |
| Norepinephrine | Catecholaminergic | pg/ml | 70-1700 | Yes | Primary neurotransmitter |
| Normetanephrine | Catecholaminergic metabolite | nmol/l | 0-0.9 | Yes | Methylated catecholamine metabolite |
| Epinephrine | Catecholaminergic | pg/ml | 0-140 | Yes | Primary neurotransmitter |
| Metanephrine | Catecholaminergic metabolite | nmol/l | 0-0.5 | Yes | Methylated catecholamine metabolite |
| Serotonin | Serotonergic | ng/ml | 50-200 | Yes | Primary neurotransmitter |
| Glutamate | Excitatory amino acid | umol/l | 10-50 | Yes | Amino acid neurotransmitter |
| Gamma-aminobutyric acid (GABA) | Inhibitory amino acid | pmoles/ml | 500-1200 | Yes | Amino acid neurotransmitter |

| | | | | | |
|---|---|---|---|---|---|
| Glycine | Inhibitory amino acid | umol/L | 20-60 | Yes | Amino acid neurotransmitter |
| HIstamine | Histaminergic | ng/ml | 50-60 | Yes | Primary neurotransmitter |
| Taurine | Amino acid modulator | umol/L | 164-318 | Yes | Neuromodulatory amino acid |
| 3-Methoxytyramine (3-MT) | Catecholamin ergic metabolite | pmol/L | 0-180 | Yes | Catecholamine metabolite |

**Supplementary Table S1.** *Summary of the blood neurotransmitter and related analytes included in the NeuroCLAD multivariate analysis.* For each analyte, the table reports its neurochemical class, laboratory measurement unit, and the corresponding unmodified laboratory reference interval used for univariate classification. The "Included in multivariate analysis" column indicates analytes retained after quality-control filtering and used in all downstream preprocessing, dimensionality-reduction, clustering, and enrichment analyses. The "Notes" column specifies the biochemical role of each analyte (e.g., primary neurotransmitter, metabolite, or precursor) to distinguish transmitter species from downstream metabolic products and modulatory compounds. Reference ranges were used only for descriptive univariate summaries and were not rescaled or incorporated into multivariate modelling.

#### Supplementary Table S2. PCA loadings for retained principal components

| Analyte | PC1 | PC2 | PC3 | PC4 | PC5 | PC6 | PC7 | PC8 | PC9 | PC10 |
|---|---|---|---|---|---|---|---|---|---|---|
| Phenethylamine (PEA) | 0.403 | 0.072 | 0.174 | -0.108 | -0.058 | -0.006 | 0.171 | -0.064 | -0.629 | 0.176 |
| Tyrosine | 0.420 | -0.325 | -0.198 | -0.130 | -0.009 | -0.055 | 0.058 | -0.148 | -0.058 | 0.342 |
| Tyramine | -0.098 | 0.362 | -0.300 | 0.190 | 0.246 | -0.263 | -0.150 | -0.358 | -0.105 | 0.336 |
| Dopamine | 0.229 | 0.053 | -0.271 | 0.044 | 0.012 | -0.484 | -0.162 | 0.449 | 0.346 | 0.301 |
| 3,4-Dihydroxyphenylacetic acid (DOPAC) | -0.075 | -0.110 | -0.479 | 0.371 | 0.140 | 0.222 | -0.061 | 0.146 | -0.425 | -0.021 |
| 3-methoxytyramine (3-MT) | 0.109 | -0.072 | 0.441 | 0.452 | 0.167 | 0.103 | -0.061 | -0.256 | 0.131 | 0.199 |
| Norepinephrine | 0.122 | -0.283 | -0.176 | 0.367 | -0.403 | 0.132 | -0.197 | -0.336 | 0.253 | -0.180 |
| Normetanephrine | -0.093 | 0.327 | -0.209 | -0.063 | -0.014 | 0.549 | -0.396 | 0.023 | 0.001 | 0.217 |
| Epinephrine | -0.455 | -0.188 | 0.126 | 0.104 | 0.044 | -0.178 | -0.190 | 0.219 | -0.121 | 0.008 |
| Metanephrine | -0.046 | -0.196 | -0.370 | 0.047 | 0.360 | 0.113 | 0.587 | -0.160 | 0.239 | -0.047 |
| Serotonin (5-HT) | -0.313 | -0.040 | 0.125 | 0.061 | -0.208 | 0.300 | 0.409 | 0.249 | 0.106 | 0.596 |
| Glutamate | -0.296 | -0.373 | 0.023 | -0.001 | -0.246 | -0.268 | -0.137 | -0.300 | -0.150 | 0.339 |
| Gamma-aminobutyric acid (GABA) | 0.295 | -0.229 | 0.086 | 0.431 | 0.026 | 0.106 | -0.076 | 0.456 | -0.092 | -0.007 |
| Glycine | -0.068 | 0.101 | -0.285 | 0.003 | -0.641 | -0.086 | 0.208 | 0.068 | -0.147 | -0.135 |
| Histamine | -0.219 | -0.440 | -0.080 | -0.212 | 0.283 | -0.026 | -0.129 | 0.080 | -0.213 | -0.115 |
| Taurine | -0.156 | 0.284 | 0.078 | 0.453 | 0.014 | -0.299 | 0.289 | -0.006 | -0.167 | -0.158 |

**Supplementary Table S2** reports the loading coefficients for the first ten principal components derived from principal component analysis of age and sex-adjusted, z-scored blood neurotransmitter residuals. Rows correspond to individual analytes and columns correspond to principal components (PC1–PC10). Loadings indicate the relative contribution and direction of each analyte to the corresponding component and are unitless. Principal components were used solely for dimensionality reduction and construction of the clustering input space; loadings are provided for transparency and reproducibility and were not interpreted as independent biological axes.

**Supplementary Table S3: Full centroid matrix of blood neurotransmitters clusters (k=12)**

| Analyte | Cluster 1 | Cluster 2 | Cluster 3 | Cluster 4 | Cluster 5 | Cluster 6 | Cluster 7 | Cluster 8 | Cluster 9 | Cluster 10 | Cluster 11 | Cluster 12 |
|---|---|---|---|---|---|---|---|---|---|---|---|---|
| Dopamine | -0.531 | -0.007 | -0.267 | 1.701 | 0.170 | 0.490 | -0.529 | -0.337 | -0.300 | 0.113 | 0.154 | -0.823 |
| Epinephrine | 0.403 | -0.144 | -0.519 | 0.478 | -0.451 | -0.512 | -0.687 | -0.018 | -0.607 | 0.220 | -0.208 | 0.995 |
| Gamma-aminobutyric acid (GABA) | -0.489 | -0.400 | 0.479 | 0.061 | -0.281 | -0.224 | -0.142 | -0.264 | 0.253 | 1.889 | -0.124 | -0.457 |
| Glutamate | -0.079 | -0.193 | -0.056 | 0.743 | -0.475 | -1.018 | -0.864 | 0.459 | 0.367 | -0.486 | 0.138 | 0.895 |
| Glycine | -0.454 | 1.226 | -0.406 | -0.312 | -0.142 | -0.397 | -0.258 | 0.177 | 0.377 | -0.501 | 0.675 | 0.068 |
| Histamine. | 0.668 | -0.808 | 0.812 | -0.151 | -0.878 | 0.090 | -0.803 | 0.400 | -0.156 | -0.498 | 0.212 | 0.753 |
| Metanephrine. | -0.173 | -0.626 | 0.447 | -0.413 | 0.097 | 1.062 | -0.841 | -0.747 | -0.063 | -0.250 | 0.679 | 0.533 |
| Norepinephrine. | -0.649 | 0.714 | 1.174 | 0.028 | 0.049 | -1.022 | -0.563 | -0.502 | 0.239 | 0.127 | 0.389 | 0.107 |
| Normetanephrine. | 0.772 | 0.087 | 0.132 | -0.531 | 1.119 | -0.487 | 0.278 | -0.268 | -0.321 | -0.392 | -0.646 | 0.099 |
| Phenethylamine (PEA) | -0.324 | -0.534 | -0.283 | -0.097 | -0.377 | 0.003 | 1.841 | 0.058 | 2.229 | -0.108 | -0.121 | -0.594 |
| Serotonin (5-HT) | 0.611 | 0.364 | -0.843 | -0.216 | -0.135 | 0.016 | -0.783 | -0.764 | 0.013 | 0.442 | -0.460 | 0.888 |
| Taurine | -0.862 | -0.124 | -1.228 | -0.152 | 0.290 | 0.171 | 0.231 | 0.631 | -0.336 | 0.349 | -0.006 | 0.541 |
| Tyramine. | -0.577 | -0.784 | -0.515 | 0.219 | 1.299 | -0.196 | -0.143 | 0.589 | -0.321 | -0.321 | 0.283 | 0.114 |
| Tyrosine | -0.386 | -0.388 | 1.018 | 0.384 | -0.180 | -0.026 | -0.525 | -0.452 | 1.625 | 0.024 | 0.482 | -0.635 |
| 3,4-Dihydroxyphenylacetic acid (DOPAC) | -0.423 | -0.375 | -0.291 | -0.423 | 0.756 | -0.439 | -0.851 | -0.153 | -0.060 | 0.634 | 0.748 | 0.409 |
| 3-methoxytyramine (3-MT) | -0.225 | -0.331 | 0.366 | 0.203 | 0.179 | -0.581 | 0.786 | -0.404 | -0.367 | 0.673 | -0.339 | 0.098 |

**Supplementary Table S3** reports the mean z-scored values of age- and sex-adjusted blood neurotransmitter residuals for each of the twelve clusters identified by k-means clustering (k=12). Rows correspond to individual analytes and columns correspond to cluster centroids

(Cluster 1-Cluster 12). Values represent within-cluster means computed in the standardized residual space used for clustering and visualised in the centroid heatmap (Fig. 5). Positive and negative values indicate relative increase or decrease with respect to the cohort mean and were used to derive cluster-specific biochemical fingerprints (Table 5). Centroids for alternative clustering resolutions are not shown and were examined only in sensitivity analyses.

**Supplementary Table S4: Expanded consensus clustering metrics across tested resolutions**

| k | Proportion of Ambiguous Clustering (PAC) | Mean-within cluster consensus | Minimum cluster size (K-means) | Maximum cluster size (K-means) |
|---|---|---|---|---|
| 8 | 0.3553 | 0.4904 | 24 | 50 |
| 10 | 0.2875 | 0.4314 | 19 | 33 |
| 12 | 0.2436 | 0.4179 | 14 | 32 |
| 15 | 0.1923 | 0.3965 | 12 | 24 |
| 20 | 0.1424 | 0.3944 | 1 | 19 |

**Supplementary Table S4**: Consensus clustering metrics. For each tested number of clusters (*k*), the table reports the Proportion of Ambiguous Clustering (PAC) and the mean within-cluster consensus, derived from the consensus clustering procedure, together with the minimum and maximum cluster sizes of the corresponding k-means candidate solutions used in downstream analyses. These complementary metrics summarize clustering performance across candidate resolutions, complement the consensus matrices and stability trends shown in Fig. 6 and Supplementary Figs. S6–S7, and provide the quantitative basis for selecting $k = 12$ as a balanced resolution that minimizes clustering ambiguity while preserving biologically interpretable cluster structure without excessive fragmentation..

**Supplementary Table S5:** Complete cluster-wise enrichment statistics for all FDR-significant analytes, including effect sizes and group means are provided in the Supplementary Table S5. *Supplementary Table S5 is provided as a machine readable CSV file to enable full transparency and reproducibility of the enrichment analysis*.
Cluster-wise biochemical enrichment results for the blood neurotransmitter cohort (k=12). For each cluster, analytes showing significant differences relative to the remainder of the cohort are reported (Benjamini–Hochberg FDR $< 0.05$). The table includes cluster-specific mean values, mean values in the complementary group, Cohen's *d* effect sizes, nominal *p*-values, and FDR-adjusted *q*-values. Rows are ordered by increasing FDR within each cluster.

**Supplementary Table S6:** The number of individuals contributing to each cluster-wise symptom percentage, along with absolute counts is provided in Supplementary Table S6.
Cluster-wise incidence of qualitative clinical symptoms for the blood cohort (k =12), showing the number of affected individuals, the number assessed for each symptom, and the corresponding percentage. Percentages reported in the main text and figures were calculated using these counts. This table is provided as a machine-readable CSV file. Severity ratings were excluded.

**Supplementary Table S7:** Association between qualitative clinical symptoms and the k=12 biochemical clusters.

| Symptom | Statistical test | $\chi^2$ | Raw p-value | BH-adjusted p-value |
|---|---|---|---|---|
| Eye Contact | Chi-square (Monte Carlo) | 12.582 | 0.320 | 0.775 |
| Stereotypical Behavior | Chi-square (Monte Carlo) | 12.433 | 0.322 | 0.775 |
| Aggressiveness | Chi-square (Monte Carlo) | 13.109 | 0.278 | 0.775 |
| Self Injurious Behavior | Pearson $\chi^2$ | 13.408 | 0.268 | 0.775 |
| Cognition | Chi-square (Monte Carlo) | 12.024 | 0.357 | 0.775 |
| Verbal | Pearson $\chi^2$ | 16.281 | 0.131 | 0.775 |
| Attention / Concentration | Chi-square (Monte Carlo) | 10.841 | 0.459 | 0.807 |
| Command Following | Pearson $\chi^2$ | 8.075 | 0.707 | 0.807 |
| Sitting Tolerance | Pearson $\chi^2$ | 9.155 | 0.608 | 0.807 |
| Hyperactivity | Chi-square (Monte Carlo) | 7.890 | 0.745 | 0.807 |
| Social Interaction | Chi-square (Monte Carlo) | 7.544 | 0.757 | 0.807 |
| Picky Eater | Pearson $\chi^2$ | 8.688 | 0.651 | 0.807 |
| Sensory Issues | Chi-square (Monte Carlo) | 7.258 | 0.807 | 0.807 |

**Supplementary Table S7:** Associations between cluster membership and qualitative clinical symptoms were evaluated using Pearson's chi-square test or Monte Carlo chi-square testing where expected cell counts violated asymptotic assumptions. Benjamini–Hochberg (BH) correction was applied across all tested symptoms. *Pearson's chi-square test was used when expected cell counts satisfied asymptotic assumptions; otherwise, Monte Carlo estimation was used. Benjamini–Hochberg correction was applied across all 13 tested symptoms. No symptom remained statistically significant after correction.*

**Supplementary Table S8:** Null-model validation of the k=12 consensus partition.

| Metric | Observed | Null model | Null median | Null 5th–95th percentile | Empirical p-value (one-sided) |
|---|---|---|---|---|---|
| PAC | 0.244 | Isotropic Gaussian | 0.261 | 0.253–0.267 | <0.005 |
| PAC | 0.244 | Covariance-preserving Gaussian | 0.259 | 0.252–0.267 | <0.005 |
| Mean within-cluster consensus | 0.418 | Isotropic Gaussian | 0.367 | 0.341–0.396 | 0.005 |
| Mean within-cluster consensus | 0.418 | Covariance-preserving Gaussian | 0.377 | 0.352–0.407 | 0.015 |

**Supplementary Table S8:** Observed clustering metrics were compared with two null models: an isotropic Gaussian null and a covariance-preserving multivariate Gaussian null, each generated from 200 simulated datasets. Lower PAC values and higher mean within-cluster consensus indicate stronger clustering structure. Empirical p-values were computed using one-sided tests. *Null distributions were generated from 200 simulated datasets for each null model. The isotropic Gaussian null assumes independent standard normal variables, whereas the covariance-preserving Gaussian null retains the covariance structure of the observed principal-component scores.*

**Supplementary Table S9:** Bayesian Information Criterion (BIC) scan for Gaussian mixture model enumeration

| Number of Components (G) | Covariance Model | BIC | Interpretation |
|---|---|---|---|
| 2 | EEE | −8086.5 | Global optimum (highest BIC) |
| 3 | EEE | −8118.9 | Second-best solution |
| 1 | EEE | −8142.1 | Single-component model |
| 1 | VVV | −8142.1 | Equivalent to EEE for a single-component model |
| 4 | EEE | −8153.6 | Lower support than the optimal solution |
| 5 | EEE | −8184.2 | — |
| 6 | EEE | −8214.9 | — |
| 7 | EEE | −8232.9 | — |
| 8 | EEE | −8271.8 | — |
| 9 | EEE | −8291.4 | — |
| 2 | VVV | −8330.2 | Flexible covariance model; substantially lower support than G = 2 EEE |
| 10 | EEE | −8331.2 | — |
| 11 | EEE | −8401.6 | — |

| | | | |
|---|---|---|---|
| 12 | EEE | −8417.1 | Corresponds numerically to the consensus clustering resolution (k=12) |
| 13 | EEE | −8461.1 | — |
| 14 | EEE | −8496.0 | — |
| 15 | EEE | −8557.4 | Corresponds numerically to the higher-resolution sensitivity analysis (k=15) |
| 16 | EEE | −8566.4 | — |
| 17 | EEE | −8573.7 | — |
| 18 | EEE | −8651.3 | — |
| 19 | EEE | −8668.6 | — |
| 20 | EEE | −8706.6 | Highest number of mixture components evaluated |

**Supplementary Table S9:** Gaussian mixture models were evaluated using Bayesian Information Criterion (BIC) over candidate solutions ranging from G = 1–20. Among the fitted models retained in the BIC scan, the highest BIC was obtained for the G = 2 EEE solution (BIC = −8086.5). The G = 2 VVV model yielded substantially lower support (BIC = −8330.2), indicating that additional covariance flexibility did not improve model fit for the two-component solution. *EEE denotes equal volume, equal shape and equal orientation, whereas VVV denotes variable volume, variable shape and variable orientation.*

**Supplementary Table S10:** Cross-tabulation between the optimal Gaussian mixture model and the consensus k=12 partition

| Consensus Cluster | Component 1 (n = 222) | Component 2 (n = 39) |
|---|---|---|
| C1 | 22 | 1 |
| C2 | 23 | 0 |
| C3 | 17 | 1 |
| C4 | 23 | 2 |
| C5 | 24 | 0 |
| C6 | 21 | 1 |
| C7 | 3 | 12 |
| C8 | 15 | 7 |
| C9 | 0 | 14 |
| C10 | 22 | 1 |
| C11 | 20 | 0 |
| C12 | 32 | 0 |

**Supplementary Table S10:** Cross-tabulation of the optimal Gaussian mixture model (G = 2, EEE covariance) against the primary k=12 partition selected following consensus-stability assessment. Values represent the number of individuals assigned to each Gaussian mixture component within each consensus cluster. The smaller Gaussian mixture component (Component 2; n = 39) was concentrated predominantly within consensus Clusters 7–9, comprising all individuals assigned to Cluster 9 (14/14), 12 of 15 individuals in Cluster 7 (80.0%), and 7 of 22 individuals in Cluster 8 (31.8%). The remaining clusters contributed only sparse membership to Component 2.

### SUPPLEMENTARY FIGURES

**Figure S1. Analyte- and sample-level missingness.**

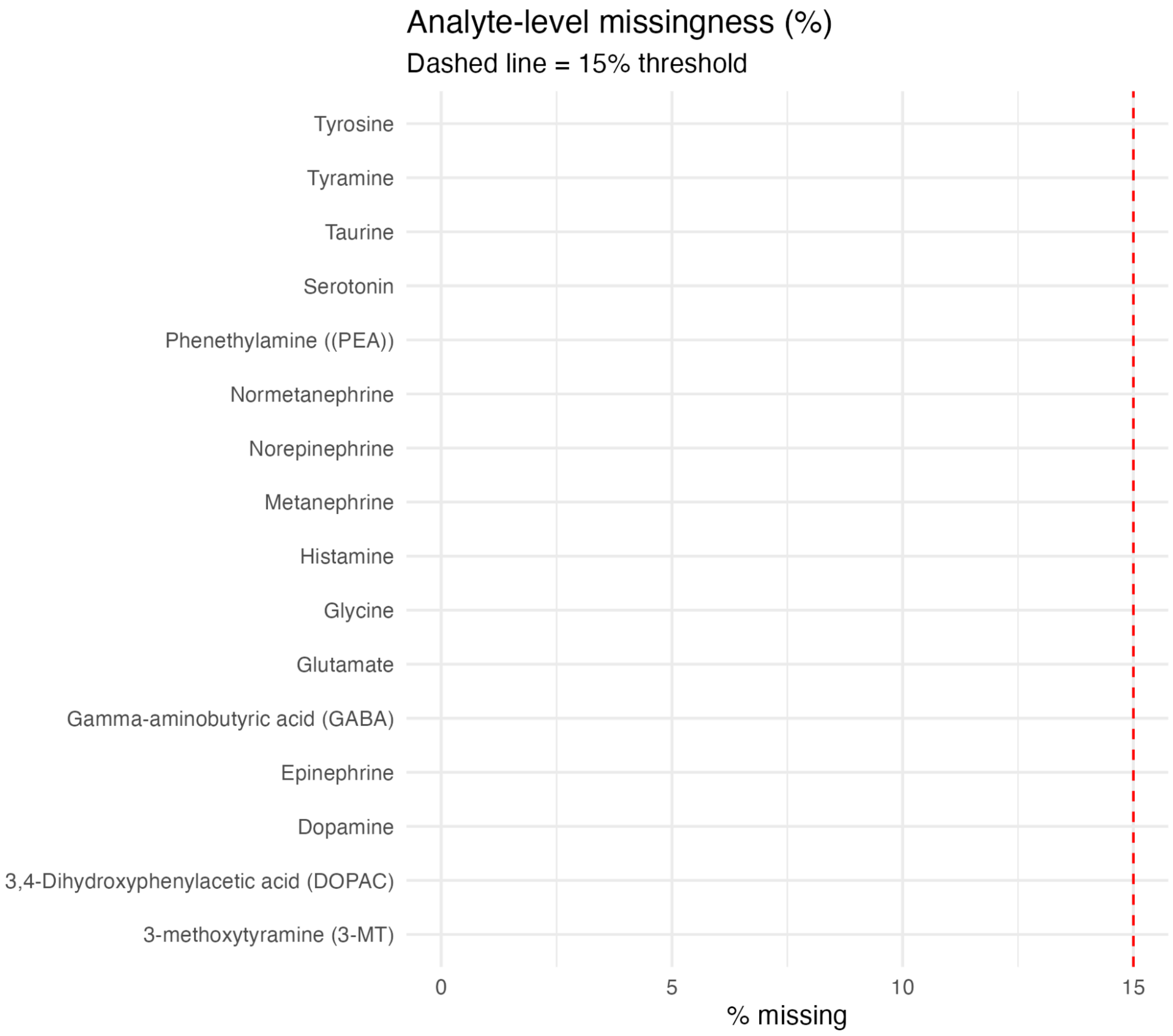

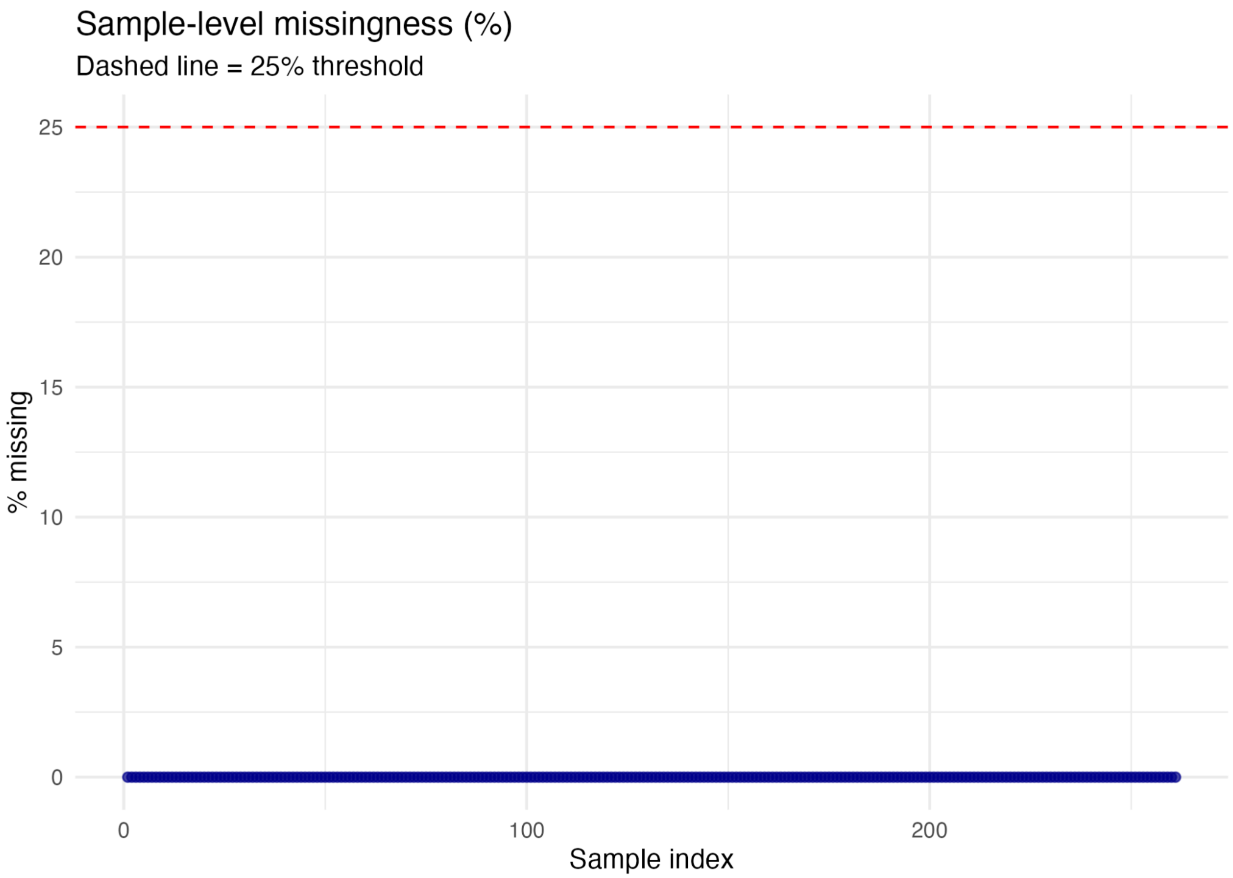


**Figure S1.** *Analyte- and sample-level missingness.* (A) Bar plot summarising the percentage of missing values per analyte; the vertical dashed line indicates the analyte exclusion threshold (>15% missing). (B) Scatter plot showing the percentage of missing values per sample; the horizontal dashed line indicates the sample exclusion threshold (>25% missing). These quality-control metrics guided pre-processing prior to multivariate analysis.

**Figure S2. Residualisation of neurotransmitter analytes after age–sex adjustment.**

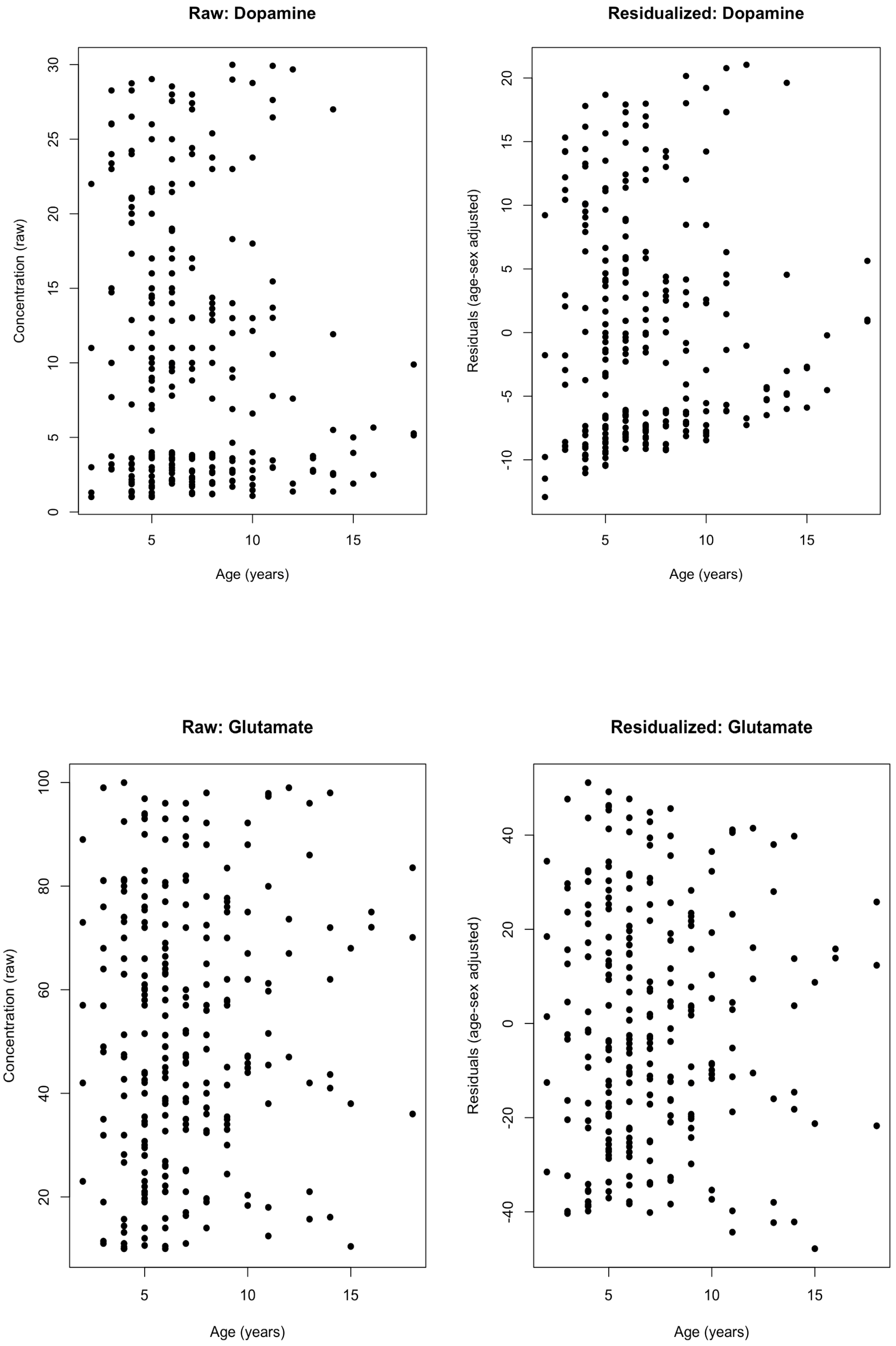

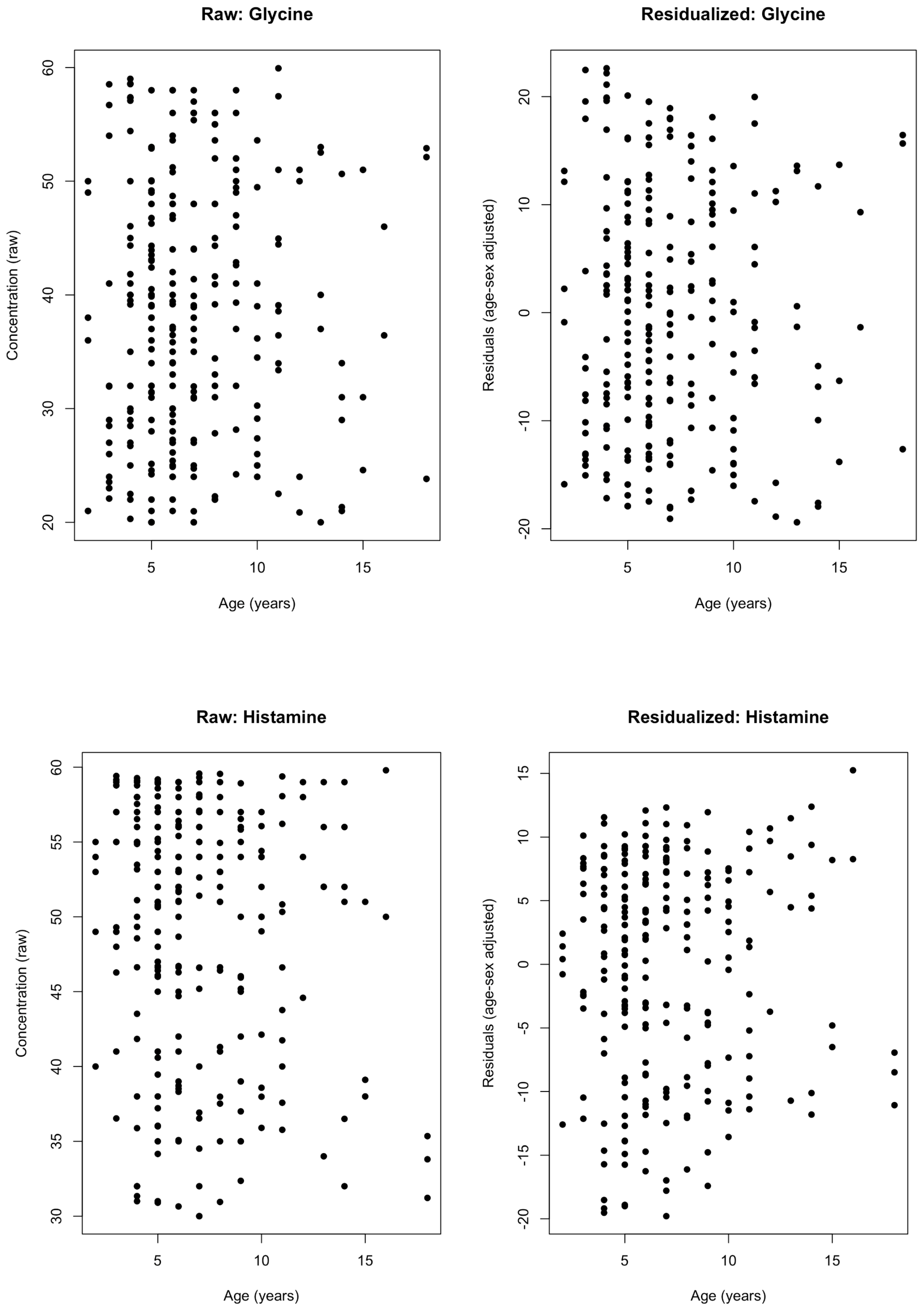

Raw: Glycine
Concentration (raw)
20
30
40
50
60
5
10
15
Age (years)
Residualized: Glycine
Residuals (age-sex adjusted)
-20
-10
0
10
20
5
10
15
Age (years)
Raw: Histamine
Concentration (raw)
30
35
40
45
50
55
60
5
10
15
Age (years)
Residualized: Histamine
Residuals (age-sex adjusted)
-20
-15
-10
-5
0
5
10
15
5
10
15
Age (years)

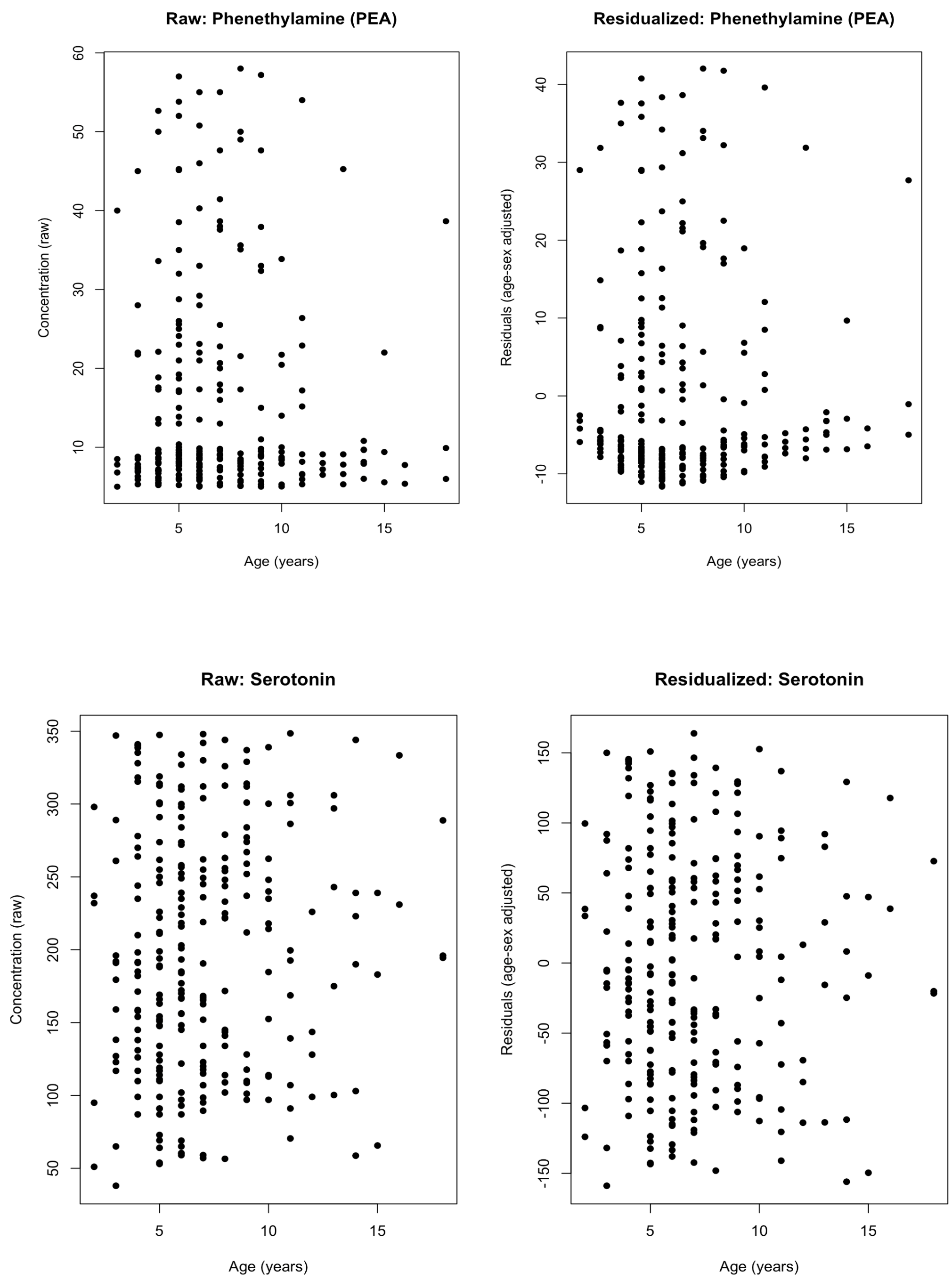


**Figure S2.** *Residualisation of neurotransmitter analytes after age–sex adjustment.* Representative examples showing raw concentrations (left) and covariate-adjusted residuals (right) plotted against age for selected neurotransmitter and related metabolites. Residuals were obtained using natural cubic spline regression with age and sex as covariates, removing non-linear age-associated structure while retaining inter-individual variability. These examples extend the GABA illustration shown in the main text and demonstrate the general applicability of the residualisation procedure across analytes.

**Figure S3. PCA loadings and extended PCA scatterplots.**

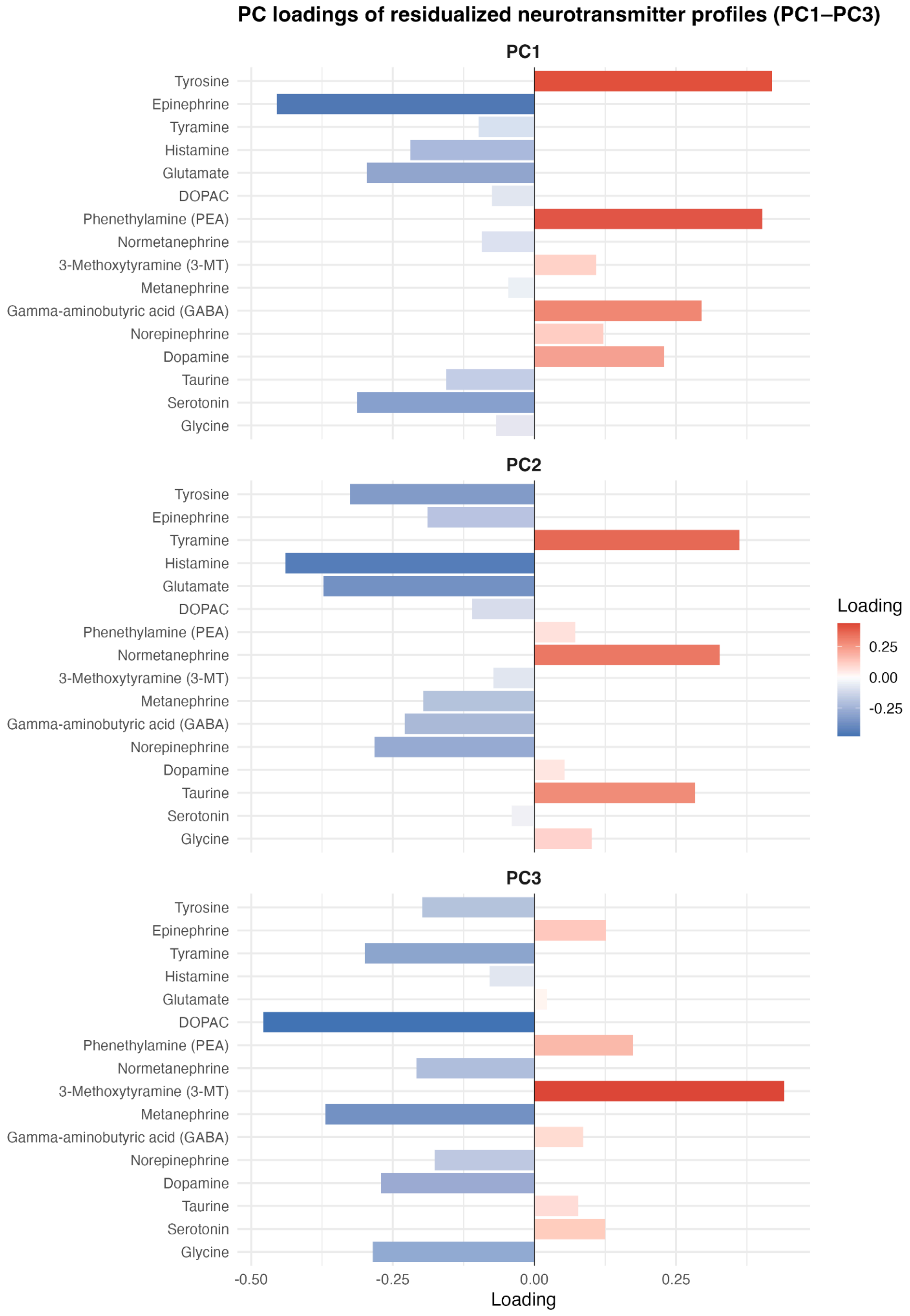

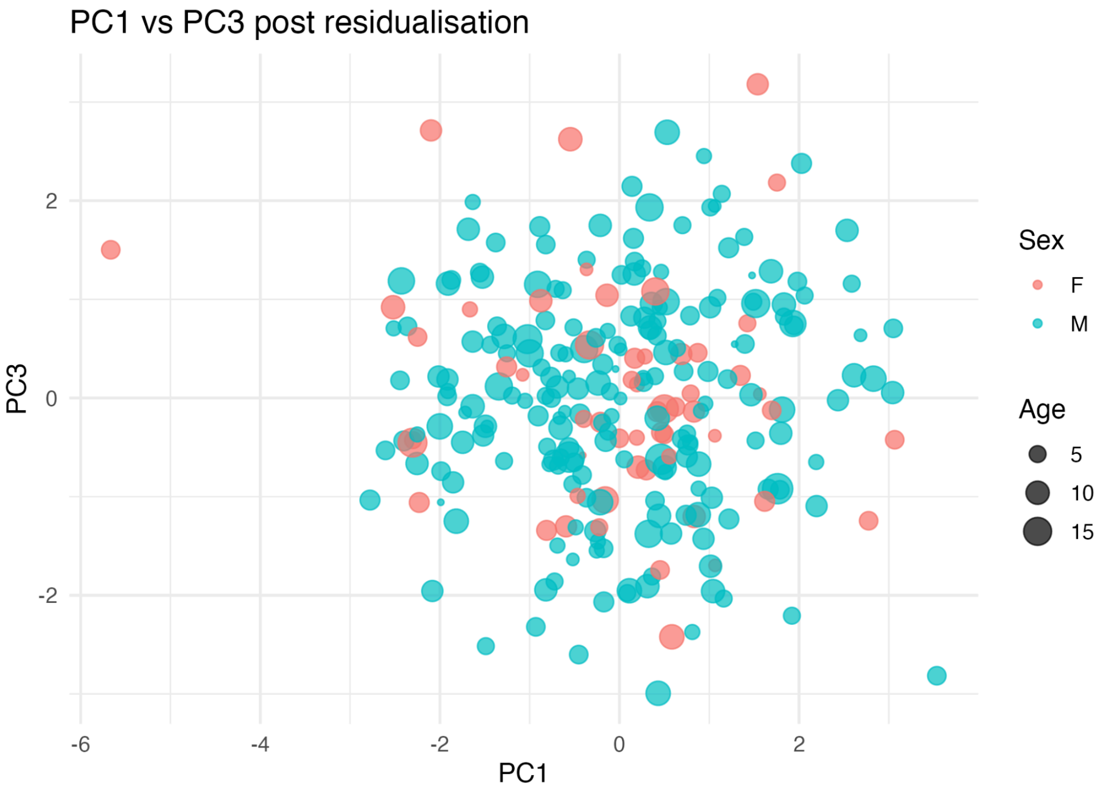
PC1 vs PC3 post residualisation
PC3
PC1
2
0
-2
-6
-4
-2
0
2
Sex
F
M
Age
5
10
15

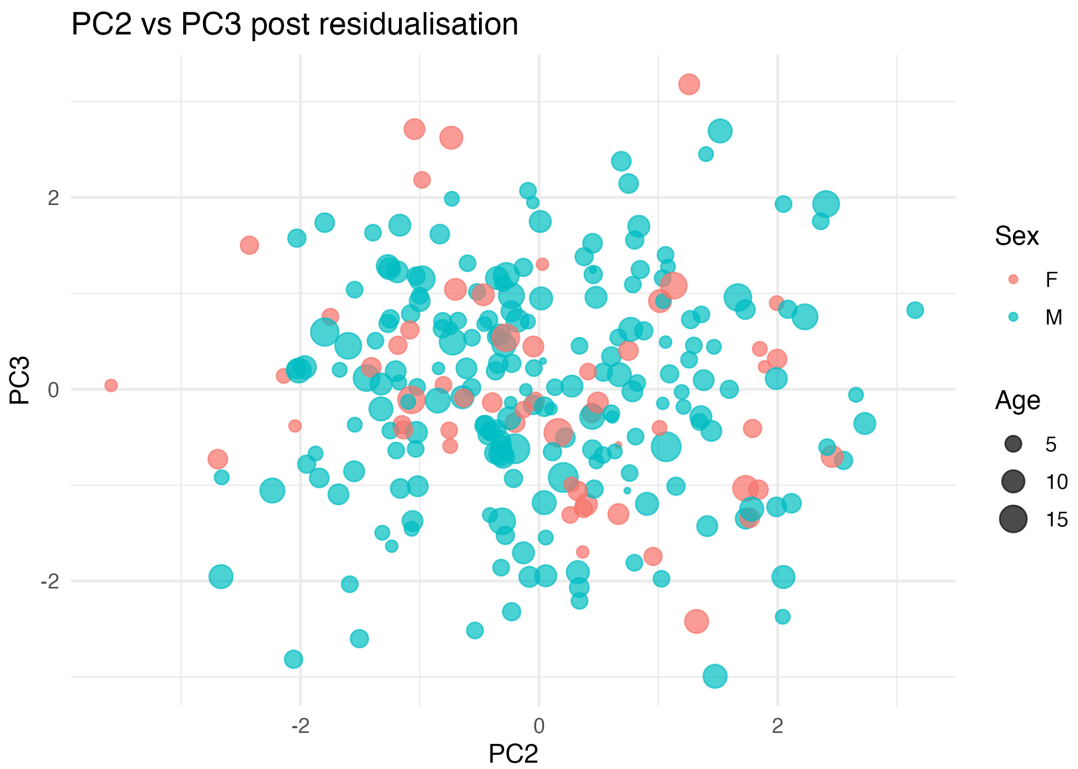
PC2 vs PC3 post residualisation
PC3
PC2
2
0
-2
-2
0
2
Sex
F
M
Age
5
10
15

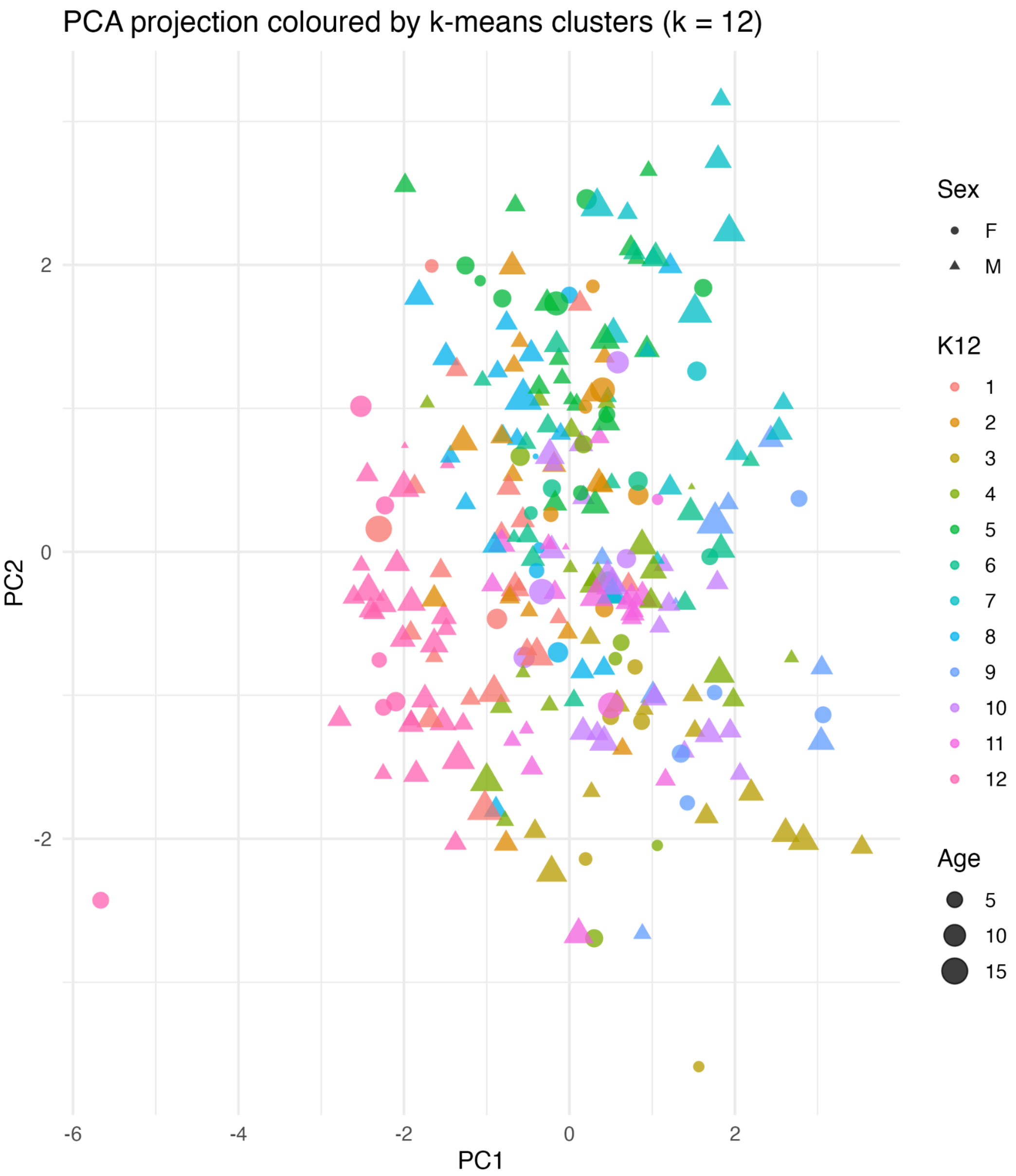


**Figure S3.** *Principal component loadings and extended PCA scatterplots.* (A) Loading coefficients for the first three principal components (PC1–PC3) derived from the residualised, z-scored analyte matrix.
(B–C) Scatterplots of PC1 vs PC3 and PC2 vs PC3, with points annotated by sex and age, illustrate the distribution of individuals in reduced-dimensional space following spline-based age–sex adjustment. These visualisations demonstrate that retained principal components primarily capture biochemical variation rather than residual demographic structure. (D) Scatterplot of PC1 vs PC2 coloured by k-means cluster assignments (k = 12), with symbol shape indicating sex and point size reflecting age. The projection shows partial overlap between clusters in the reduced-dimensional space, consistent with stable cluster cores embedded within a continuous multivariate biochemical landscape.

**Figure S4. UMAP embedding of residualised analytes.**

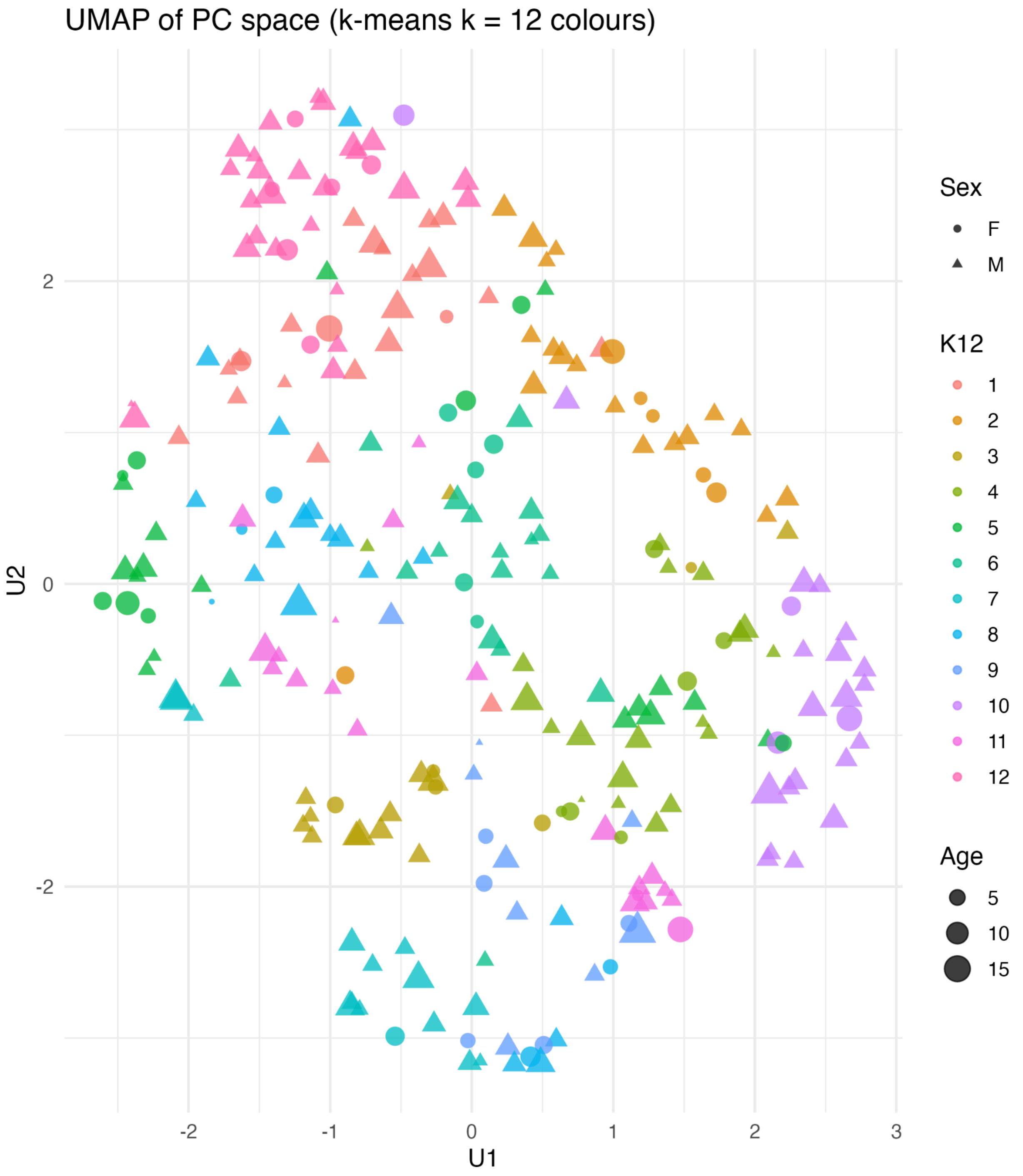

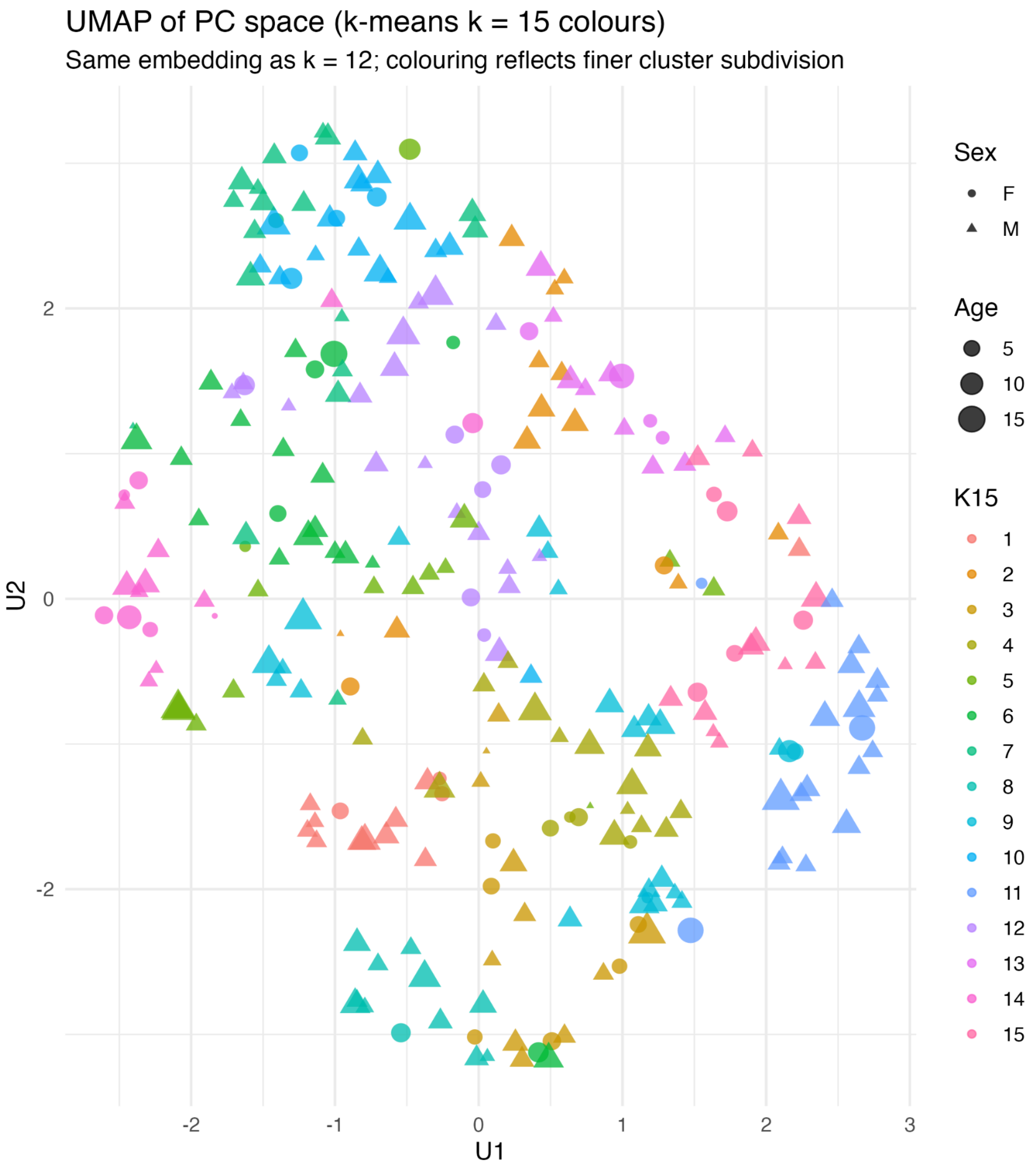


**Figure S4.** *UMAP embedding of residualised analytes across clustering resolutions.* (A) Two-dimensional UMAP projection of the retained principal component space, with points coloured by k-means cluster assignments at *k=12*. The embedding provides a low-dimensional visualisation of the multivariate biochemical structure after age–sex adjustment, illustrating partial overlap and gradual transitions among clusters rather than sharp separations. (B) The same UMAP embedding coloured by k-means assignments at *k=15*, demonstrating finer subdivision of the k=12 structure without introduction of additional, biochemically independent regions. The additional clusters primarily partition existing neighbourhoods in PC space, indicating increased resolution rather than fundamentally new biochemical organisation.

**Figure S5. Centroid heatmaps for alternative cluster resolutions (k = 3, 15).**

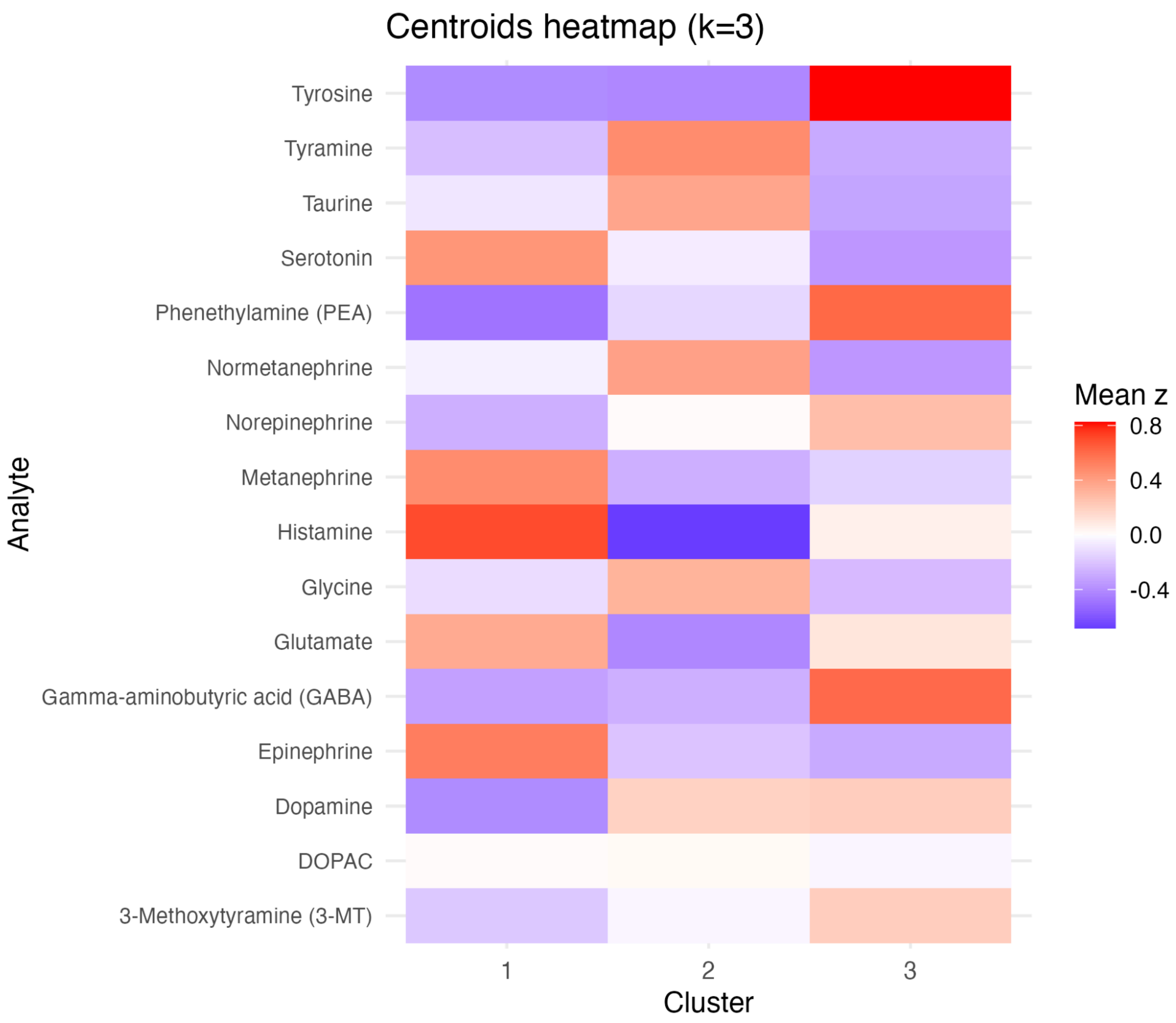

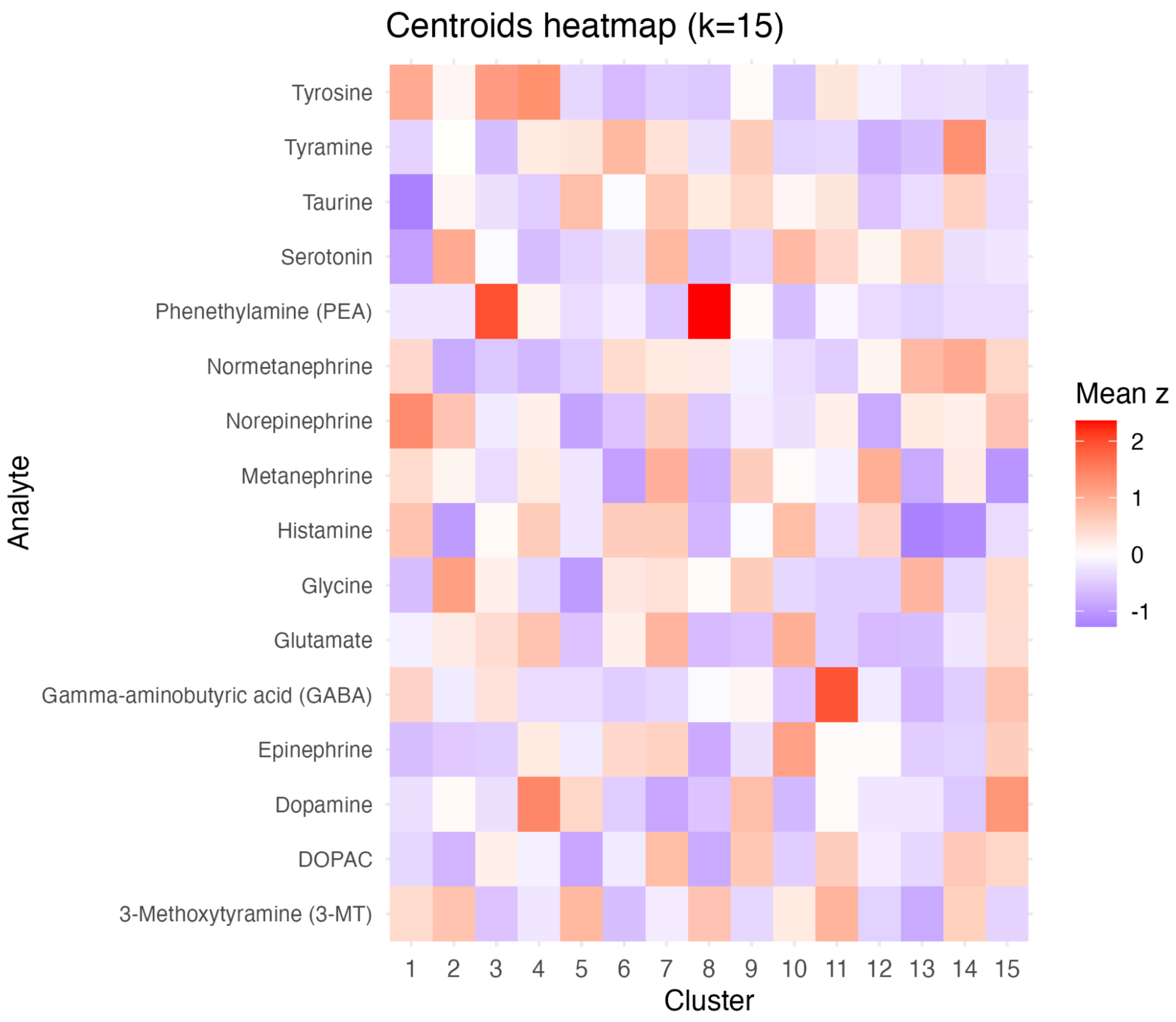


**Figure S5. Centroid heatmaps for alternative clustering resolutions (k = 3 and k = 15).** Heatmaps showing mean z-scored analyte levels for lower- and higher-resolution clustering solutions. A) The k = 3 solution summarises broad biochemical patterns across analytes, whereas B) the k = 15 solution resolves finer-scale variation in centroid profiles. These panels illustrate the effect of clustering resolution on the granularity of biochemical patterns.

**Figure S6. Consensus matrices across k = 8, 15, and 20.**

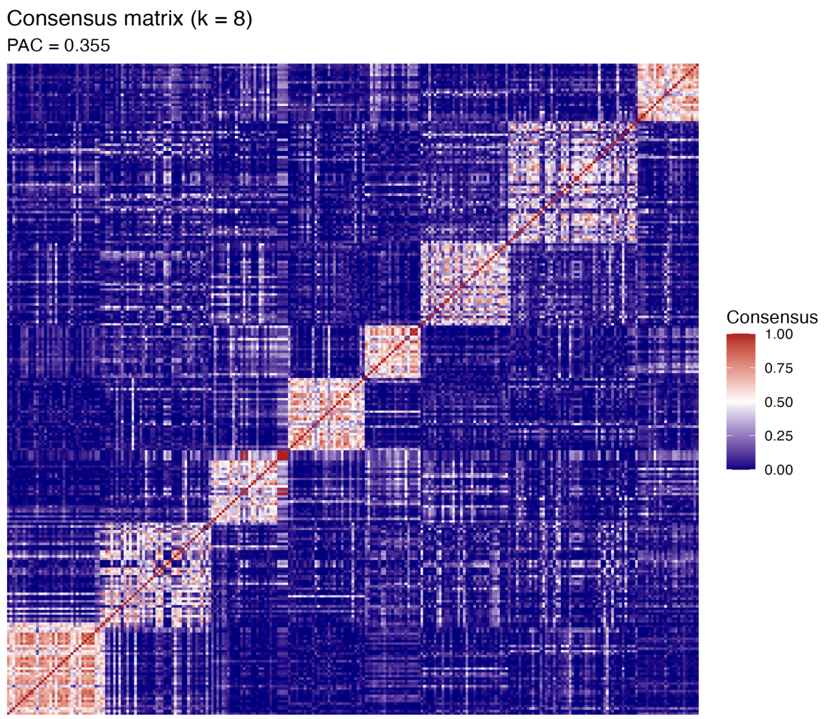


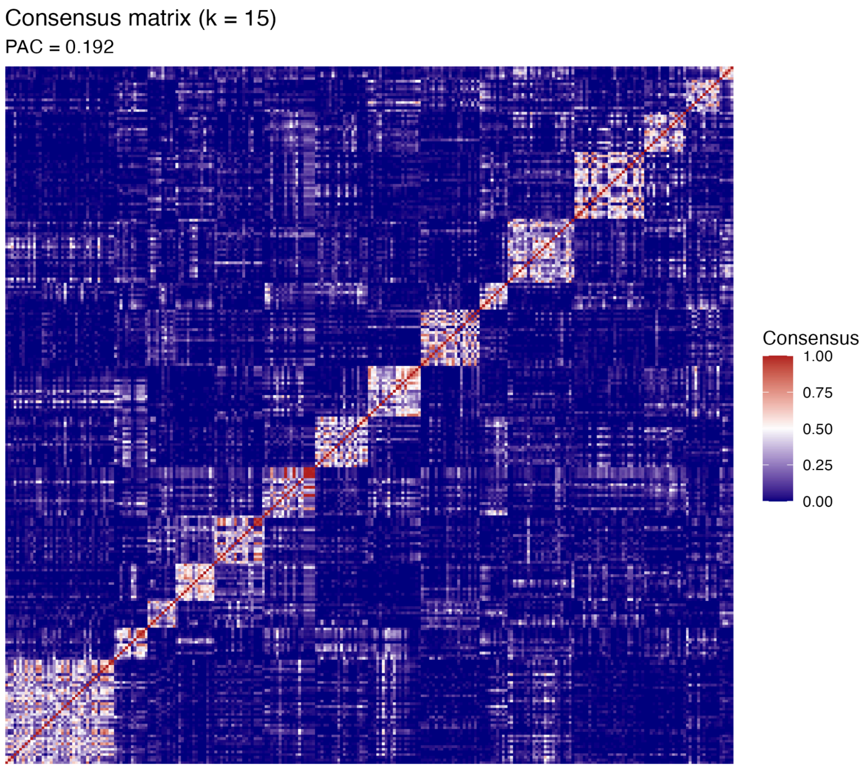

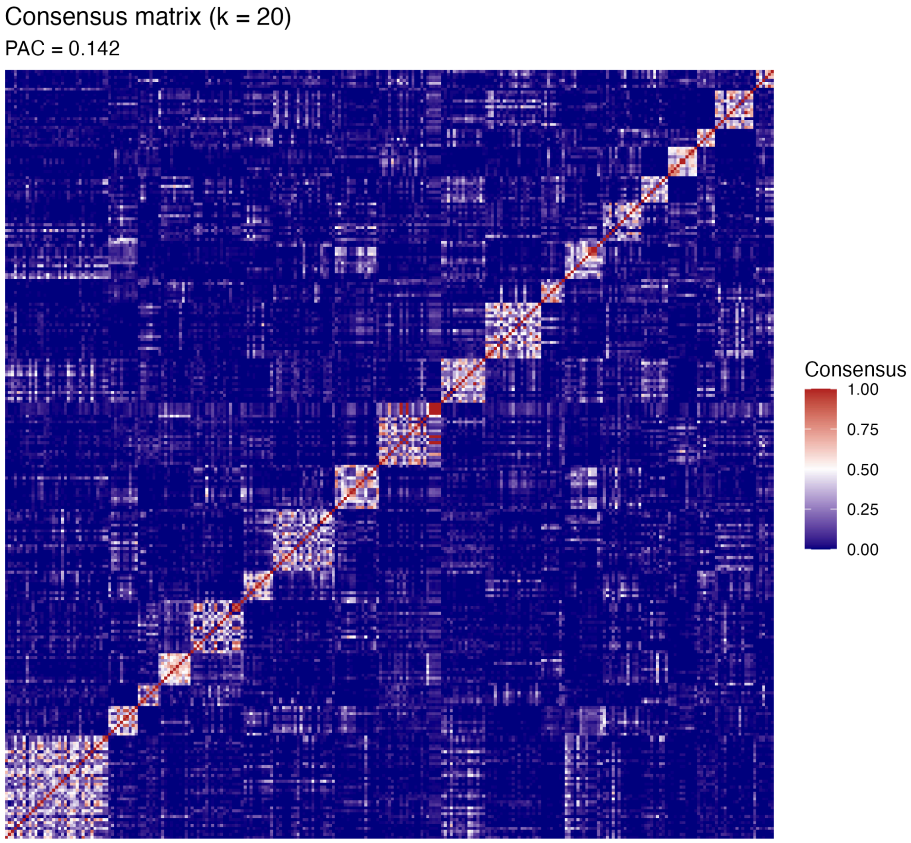


**Figure S6.** *Consensus clustering stability across alternative resolutions.* Consensus matrices showing pairwise co-clustering probabilities across 200 bootstrap resamples for alternative cluster resolutions. At k = 8, broad diagonal blocks indicate coarse, heterogeneous groupings with substantial within-cluster variability. At k = 15, clusters appear visually sharper but are subdivided into smaller units with reduced overall stability. At k = 20, further fragmentation produces narrow high-consensus blocks alongside widespread low-consensus regions, consistent with over-partitioning of the data. Together, these patterns illustrate the trade-off between apparent sharpness and stability, motivating selection of an intermediate resolution in the main analysis.

**Figure S7. PAC and stability curves across k = 8–20.**

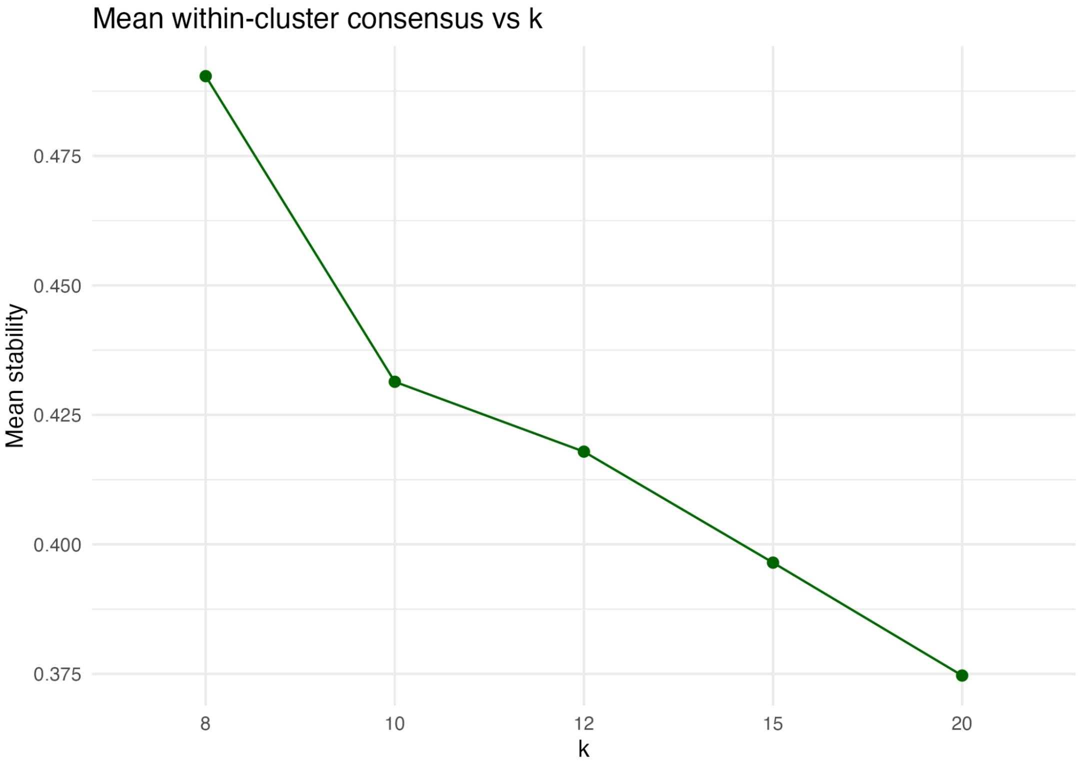

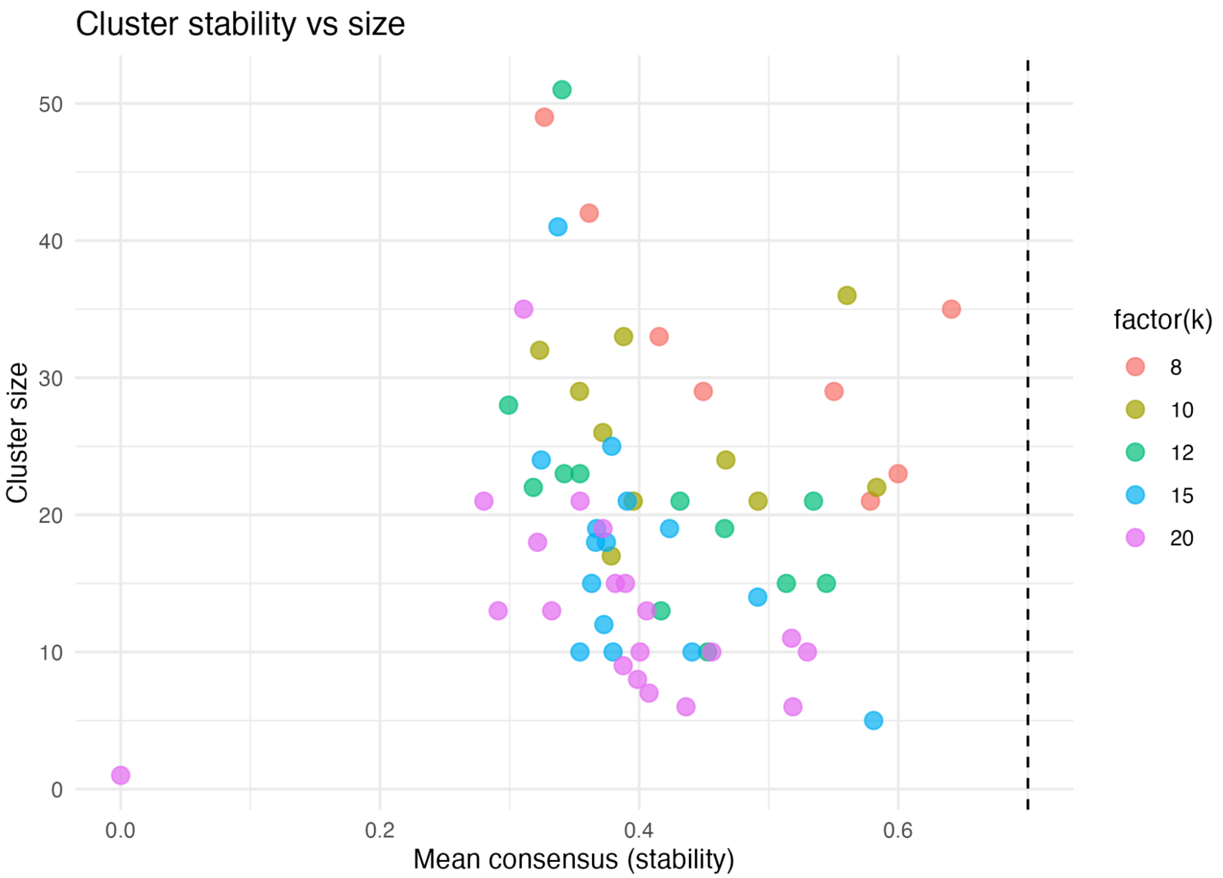


**Figure S7.** *Stability diagnostics across candidate clustering resolutions.* (A) Mean within-cluster consensus across k=8,10,12,15, and 20, demonstrating a gradual decline as clustering resolution increases. (B) Relationship between cluster size and within-cluster consensus across the evaluated resolutions. Higher-k solutions contain a greater number of small clusters with more variable stability, whereas the k=12 solution occupies an intermediate regime between coarse grouping and excessive fragmentation.

**Figure S8. Null-model validation of the k = 12 consensus partition.**

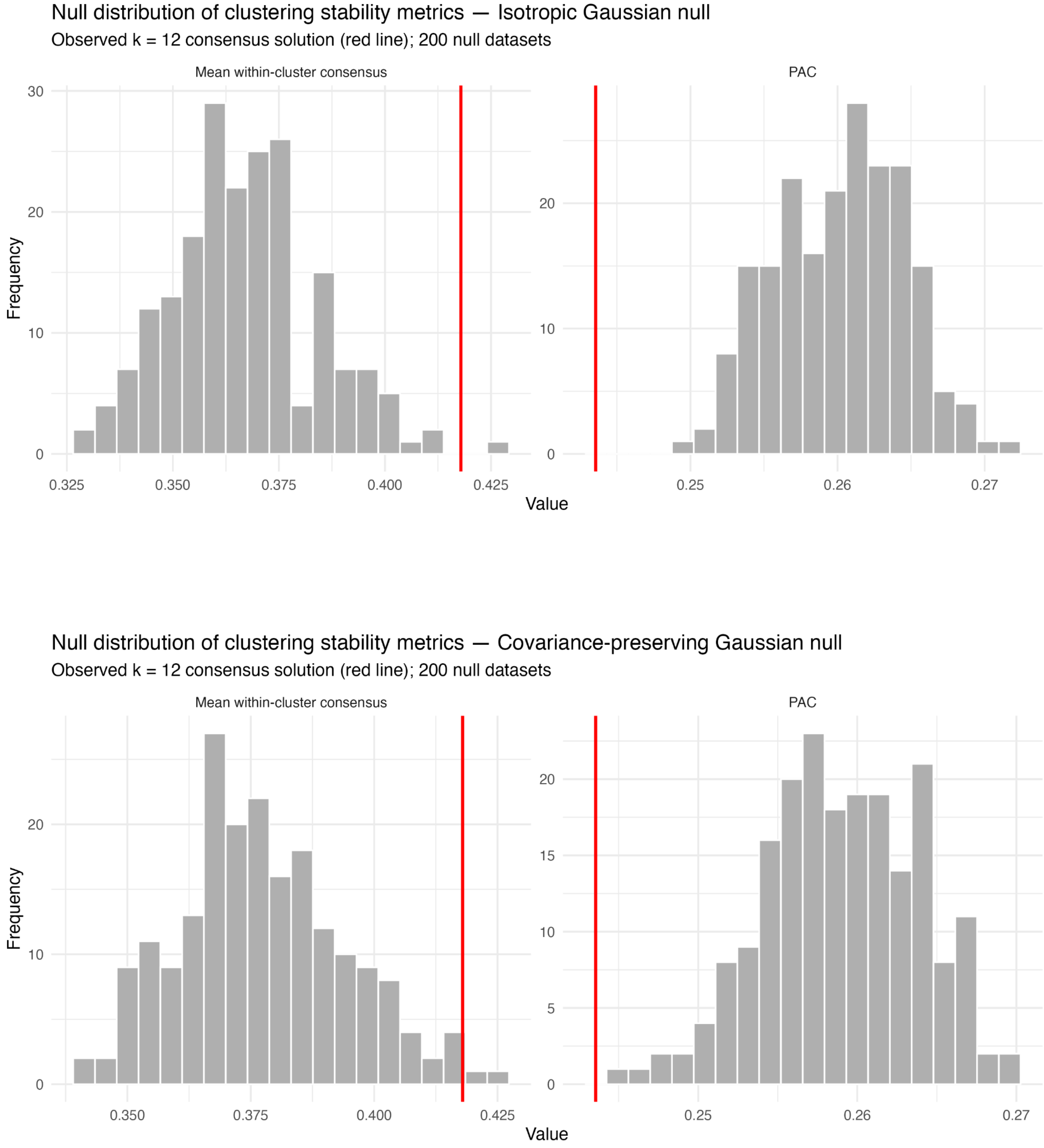


**Figure S8.** Null-model validation of the k = 12 consensus partition. (A) Distributions of mean within-cluster consensus and the proportion of ambiguous clustering (PAC) obtained from 200 isotropic Gaussian null datasets. (B) Corresponding distributions obtained from 200 covariance-preserving Gaussian null datasets that retain the observed covariance structure of the principal component space. In each panel, the red vertical line denotes the observed value from the k = 12 consensus solution. Lower PAC and higher mean within-cluster consensus indicate stronger clustering structure. The observed partition exceeds both null models, with a larger margin over the isotropic null than over the covariance-preserving null, consistent with reproducible organisation beyond that expected from random variation or covariance structure alone.

**Figure S9. Bayesian Information Criterion (BIC) across candidate Gaussian mixture models.**

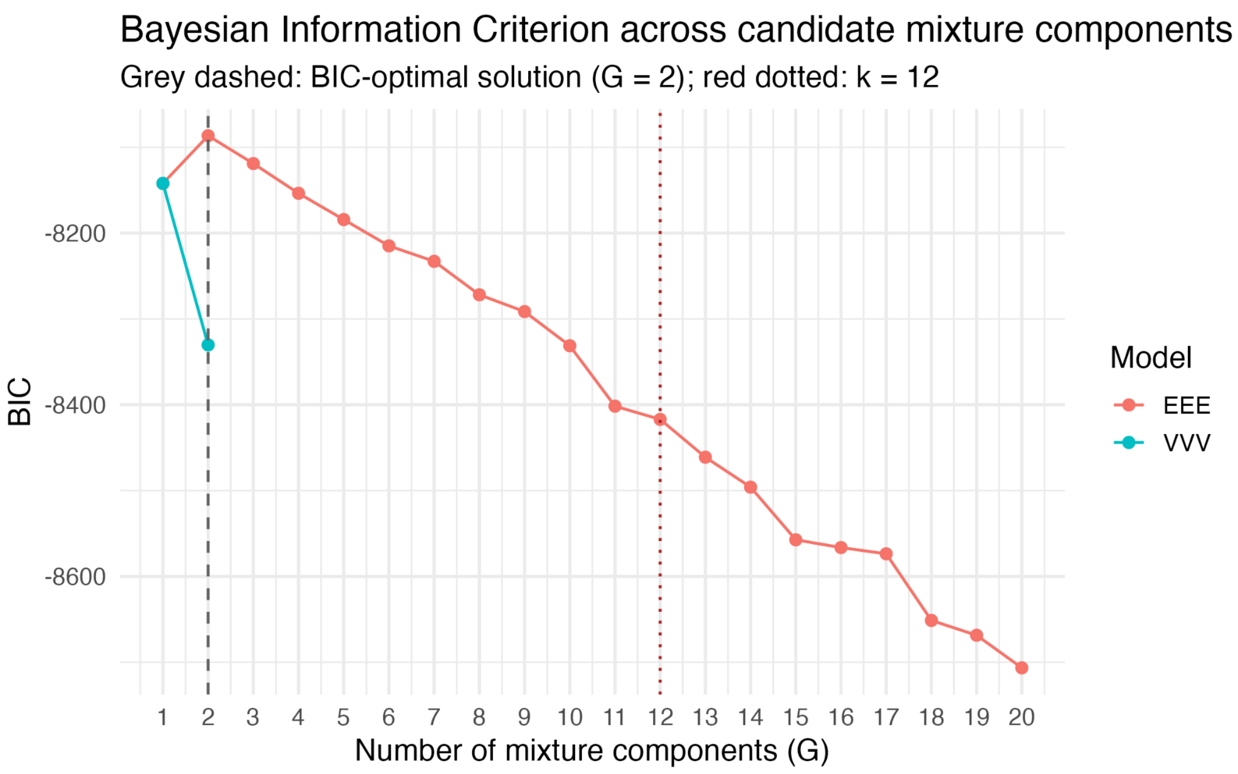


**Figure S9.** Bayesian Information Criterion (BIC) across candidate Gaussian mixture models. Bayesian Information Criterion (BIC) values for the fitted Gaussian mixture models used for model enumeration. The highest BIC was obtained for the G = 2 EEE solution, while the G = 2 VVV solution showed substantially lower support. The grey dashed line denotes the BIC-optimal solution (G = 2), and the red dotted line indicates the operational k = 12 consensus clustering resolution selected for downstream analyses.

**Figure S10. Gaussian Mixture Modelling (GMM) for k = 15 clustering solution.**

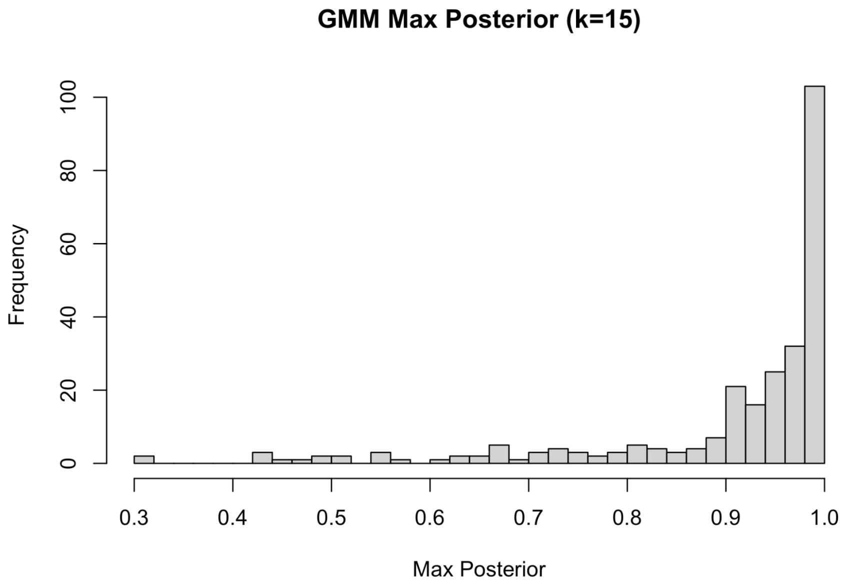


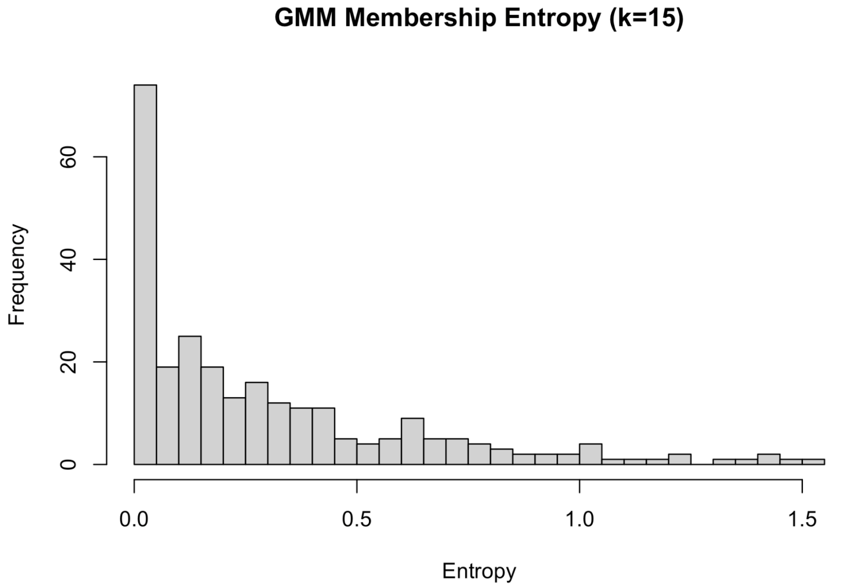


**Figure S10. Probabilistic cluster assignment characteristics for the k = 15 Gaussian mixture model solution.**

(A) Distribution of maximum posterior probabilities across individuals for k = 15. The majority of individuals retain high posterior probabilities, indicating the presence of well-defined cluster cores even at higher resolution. However, a broader spread toward lower values compared to k = 12 suggests reduced assignment confidence for a subset of individuals.

(B) Distribution of entropy values derived from posterior membership probabilities. Entropy values show increased dispersion relative to k = 12, reflecting greater uncertainty in cluster membership for a subset of individuals. This pattern indicates that increasing k introduces finer partitions without strengthening probabilistic separation.

Together, these results suggest that while the k = 15 solution preserves identifiable cluster cores, it increases boundary-level ambiguity, supporting its interpretation as a more fragmented representation of the same underlying biochemical structure rather than the emergence of new, well-separated subtypes.

**Figure S11. Sample-based and analyte-based alluvial diagrams (k = 12 to k = 15).**

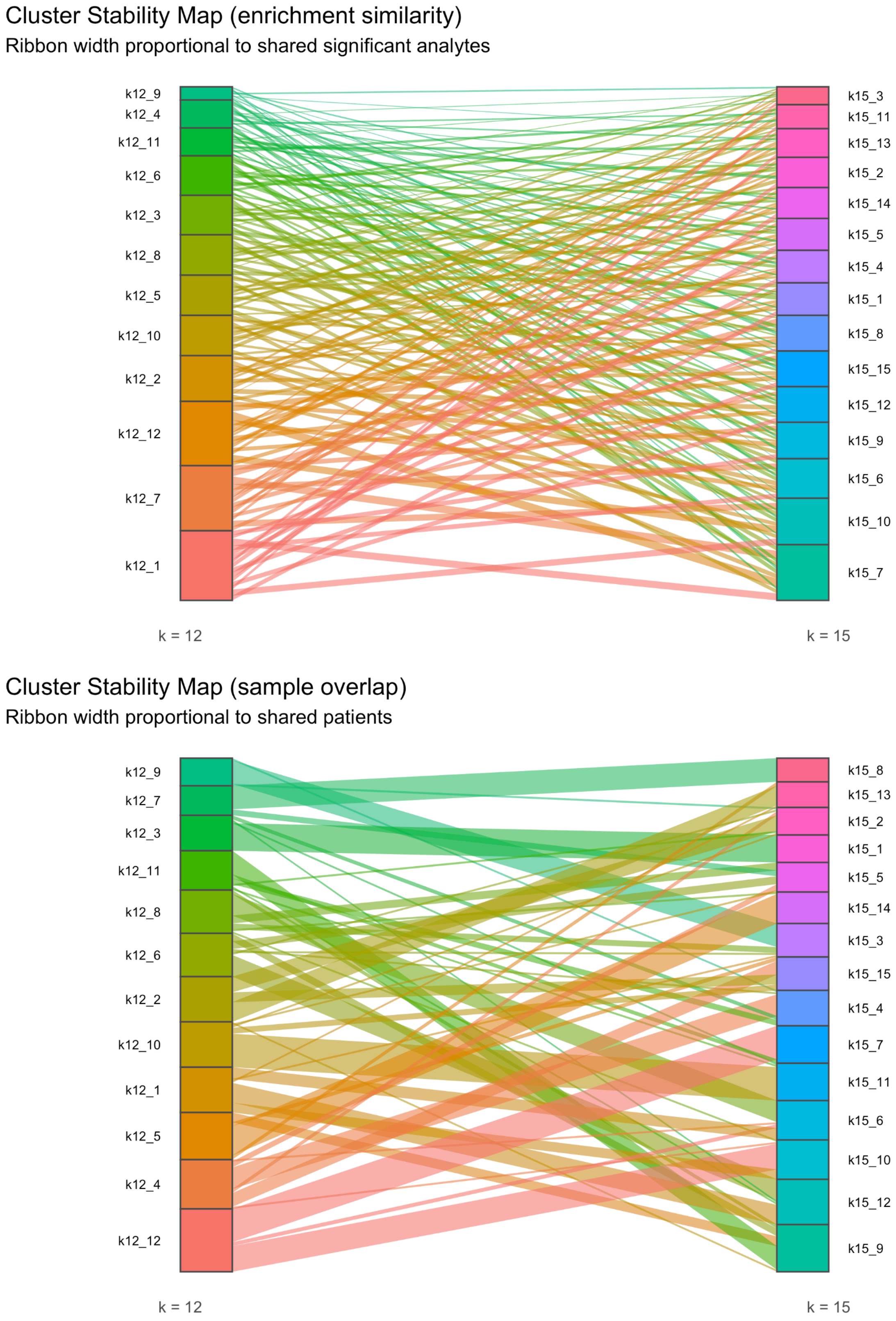

**Figure S11.** *Correspondence between k = 12 and k = 15 cluster solutions.*Alluvial diagrams illustrating how the k = 12 biochemical clusters relate to higher-resolution partitions at k = 15. (A) **Enrichment-based correspondence**, where ribbon width reflects the number of shared FDR-significant analytes between clusters. The absence of dominant or one-to-one mappings indicates dispersion of pathway-level signatures across multiple k = 15 clusters rather than preservation of discrete biochemical patterns.(B) **Patient-flow correspondence**, where ribbon width reflects the number of shared individuals. Most k = 12 clusters partition into multiple k = 15 subclusters, demonstrating increased granularity at the sample level.Together, these diagrams indicate that increasing k from 12 to 15 leads to subdivision of patient groups without corresponding preservation of pathway-level structure, consistent with over-fragmentation rather than the emergence of new biochemical organisation.

**Figure S12. Supportive Cross-Compartment Urine Analysis**

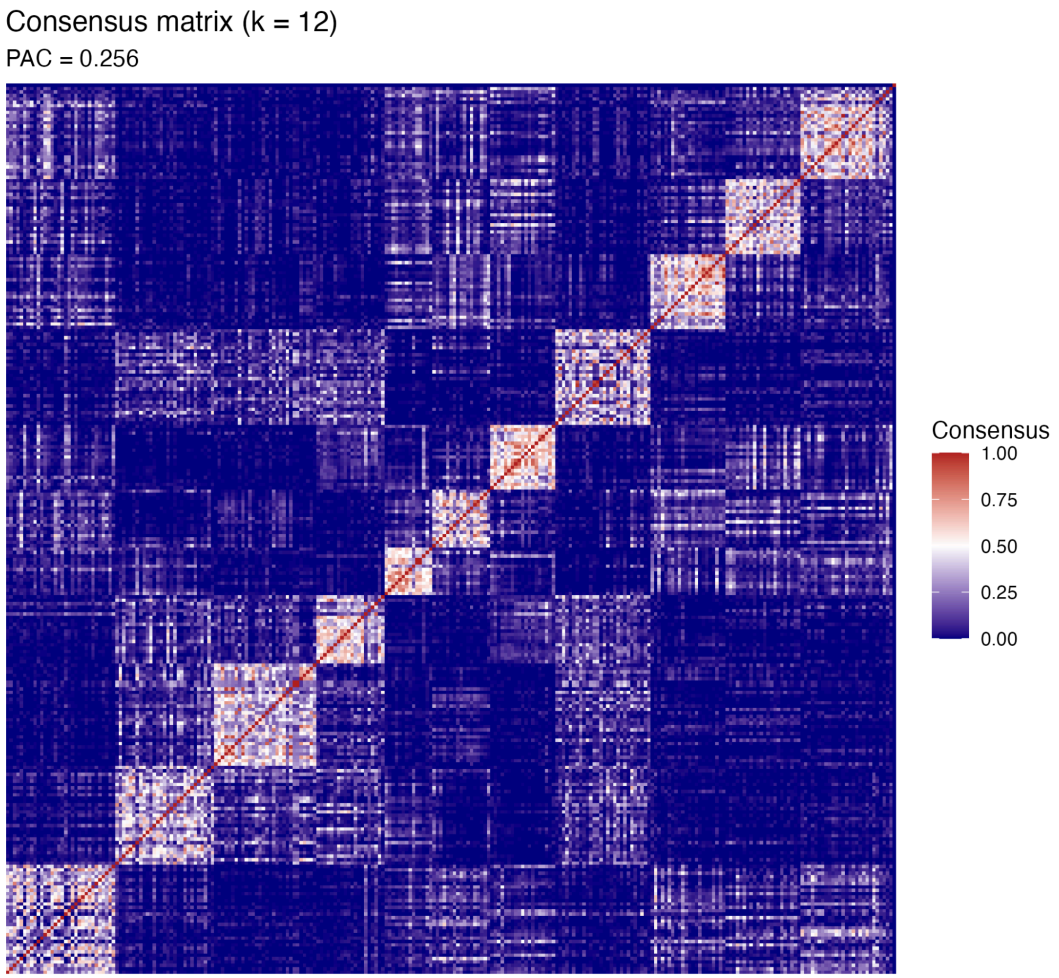

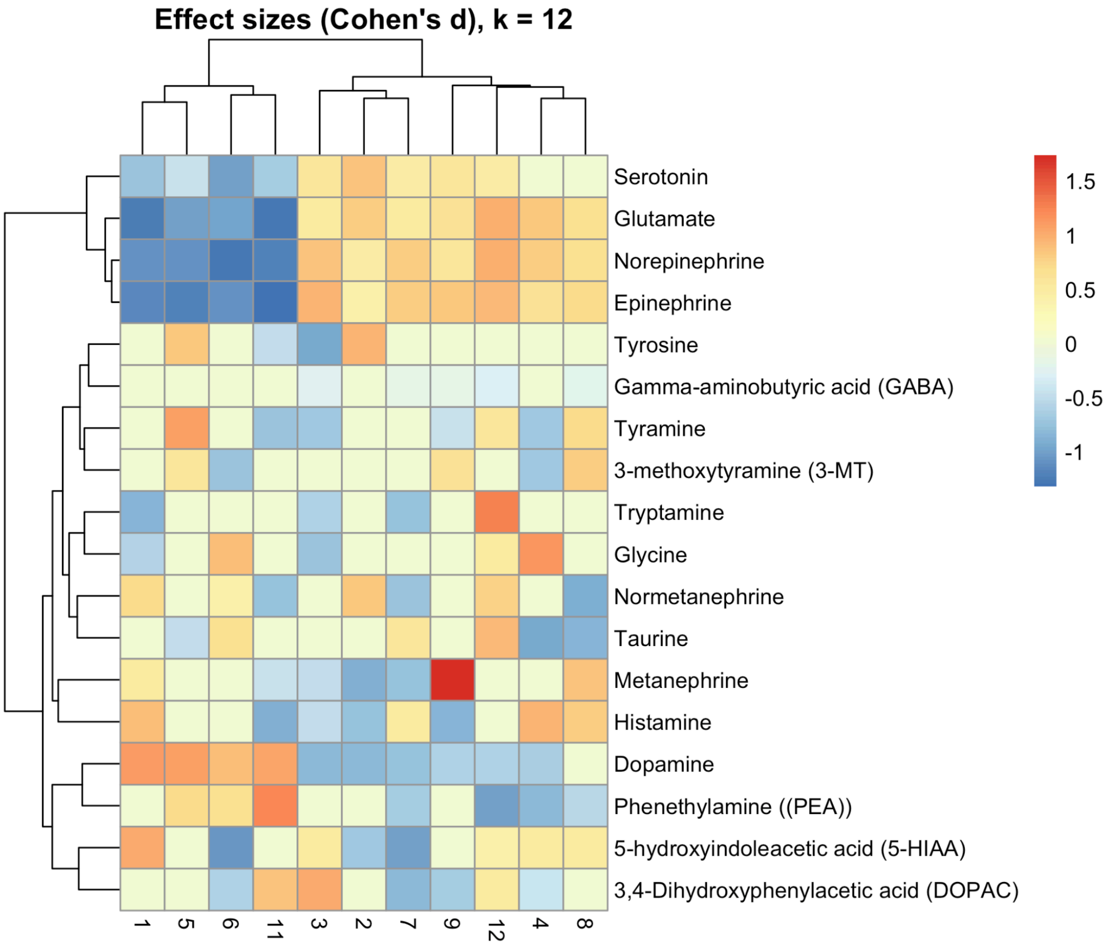


**Figure S12.** *Supportive cross-compartment comparison of urine neurochemical structure.* (A) Consensus clustering matrix for urine neurotransmitter profiles at k = 12, demonstrating stable cluster boundaries under bootstrap resampling (PAC = 0.256). (B) Cohen's d heatmap summarizing cluster-wise effect sizes for analytes overlapping between blood and urine panels. The heatmap summarises cluster-wise Cohen's d effect sizes for analytes shared between the blood and urine panels. Only analytes remaining statistically significant after Benjamini–Hochberg FDR correction are displayed, highlighting conserved module-level patterns involving catecholaminergic turnover, serotonergic metabolism, and excitatory–inhibitory amino acid balance. Together, these analyses demonstrate that urine profiles summarise key biochemical axes identified in blood, providing cross-compartment support for the inferred neurochemical landscape.
